# Early-Phase Fluid Diagnostic Biomarkers in Acute Ischemic Stroke: An Umbrella Review

**DOI:** 10.1101/2025.07.09.25331017

**Authors:** Alessandro Bombaci, Federico Emanuele Pozzi, Salvatore Mazzeo, Elisa Bortolin, Giulia Bruschi, Maria Vittoria Corbari, Alberto Astengo, Gianluca Stufano, Sergio Soeren Rossi, Maria Paola Perini, Giuseppe Rotondo, Fabiana Novellino, Massimo Filippi, Maria Salsone

## Abstract

**Background and Purpose:** Acute ischemic stroke (AIS) is a time-critical emergency in which rapid diagnosis within established therapeutic windows (≤ 4.5 h for intravenous thrombolysis; ≤ 6 h for mechanical thrombectomy) is essential to optimize outcomes. Fluid biomarkers offer a promising adjunct to clinical and neuroimaging assessment but their temporal dynamics in the acute phases remain incompletely characterized.

**Methods:** We performed an umbrella review of systematic reviews and meta-analyses evaluating fluid biomarkers in AIS versus controls or stroke mimics. Quantitative synthesis of primary studies (random-effects meta-analysis of standardised mean differences [eG]) was stratified by clinically relevant time windows. Heterogeneity (I²), small-study effects (Egger’s test) and excess significance bias were assessed.

**Results:** We included 27 publications (18 biochemistry, 1 metabolomic, 10 transcriptomic, 5 cell-free-DNA). Across all time-points the largest effect sizes were observed for neuron-specific enolase (NSE), ischemia-modified albumin (IMA), d-Dimer, S100B, GFAP, and IL-6. Looking at metabolites, studies revealed early accumulation of lactate, succinate, glutamate and lysophosphatidylcholines, alongside depletion of arginine, citrulline and citrate, indicating anaerobic glycolysis, excitotoxicity and nitric-oxide impairment. A catalogue of 220 micro-RNAs (132 upregulated; 108 downregulated) identified robust markers (miR-16-5p, let-7e-5p, miR-107, miR-451a and miR-126-3p) validated in ≥ 3 cohorts. 46 circulating-RNAs, and 55 long-non-coding-RNAs were consistently dysregulated. Five studies reported elevated nuclear (B-globin, TERT) and mitochondrial (MT-ND2; 2.5–3.5×) cfDNA within 6 h.

**Conclusions:** Fluid biomarkers exhibit a temporally evolving signature: early coagulopathy (D-dimer), glial activation (GFAP, S100B) and inflammation (IL-6), followed by neuronal necrosis (NSE) and oxidative stress (IMA) within 24 h. Multi-omic integration, including metabolomics, transcriptomics and cfDNA, highlights convergent pathways (PI3K/Akt, NF-κB, immunometabolism) and supports the development of rapid, point-of-care panels. Standardized sampling windows and harmonized assay protocols are essential for clinical translation and prospective validation in prehospital settings.

## 1. Introduction

Acute ischemic Stroke (AIS) is the second leading cause of death globally, with 15 million annual cases^1^. It is a time-sensitive neurological emergency, for which diagnostic delay translates into lost neurons and worse long-term outcomes. Prompt and accurate diagnosis, ideally within the first 4.5 hours from the onset of symptoms, is essential to guide treatments that can significantly improve outcomes

Randomized controlled trials (RCT) have demonstrated a favorable risk–benefit profile for reperfusion, balancing successful recanalization against hemorrhagic complications only when therapies are delivered within established time windows. Specifically, the NINDS rt-PA Stroke Study^2^ and the ECASS III trial^3^ demonstrated that intravenous alteplase provides a favorable net benefit within 4.5 hours of symptom onset (extended to 9 hours in select cases). Similarly, multiple RCT of mechanical thrombectomy, including MR CLEAN^4^, ESCAPE^5^, EXTEND-IA^6^, and SWIFT PRIME^7^, established that endovascular clot retrieval within six hours (extended to 24 hours in carefully selected patients) significantly improves functional outcomes without increasing hemorrhagic complications.

A major challenge is distinguishing true AIS from so-called “stroke mimics” (for example, migraine aura, seizure, functional neurological disorders or hypoglycemia). An early identification of AIS avoids loss of time in starting treatments. Rapid and accurate diagnosis of AIS not only ensures timely initiation of reperfusion therapies but also prevents the inappropriate treatment of mimics, which offers no benefit and may expose patients to serious hemorrhagic complications.

Nowadays, the diagnosis of AIS and differentiation from stroke-mimics remains clinical. Current diagnostic methods for AIS face significant limitations. Computed tomography (CT) is used to rule out contraindications to reperfusion therapies and has a sensitivity of only 57-71% in the hyperacute phase, while magnetic resonance imaging, although more sensitive, is impractical in emergencies or resource-limited settings due to logistical and time-related challenges.

In this context, fluid-based biomarkers detectable in blood are emerging as promising complement for rapid AIS diagnosis and differentiation from stroke mimics. Over the past decade, numerous primary studies have investigated a wide array of candidate markers in early (≤ 6 h) phases of cerebral ischemia, including proteins and metabolites (e.g., Iinterleukin-6 [IL-6]^8^, d-Dimer^9^, glial fibrillary acidic protein [GFAP]^10^, neuron-specific enolase [NSE]^11^, S100 calcium-binding protein B [S100B]^8^, lactate^12^, succinate^13^). In recent years, an ever-expanding body of literature has highlighted the remarkable potential of circulating nucleic/ribonucleic acids in the hyperacute phase of AIS^14^. Specifically, cell-free DNA (cfDNA) and circulating RNAs, including circular RNA (circRNA) gene/transcripts, long non-coding RNA (lncRNA), micro-RNA (miRNA), and small nuclear RNA (snRNA).

However, conflicting methodologies, heterogeneous findings, and differences in control definition (healthy subjects, mimics) have prevented a clear consensus. To address this, systematic reviews and meta-analyses have been conducted in an effort to pool quantitative effect sizes and resolve discrepancies; yet, despite these efforts, the evidence remains dispersed. Biochemistry-based meta-analyses yield quantitative effect sizes, whereas nucleic acid studies often report simple directional changes without concentration data, precluding pooling.

Therefore, we performed an umbrella review, collating and critically appraising all existing evidence to generate a coherent, overarching picture. We aimed to synthesize these disparate findings into a unified evidence map, stratified by clinically relevant time windows, and to identify the most promising fluid protein, metabolite and nucleic/ribonucleic acid biomarkers for rapid AIS diagnosis and stroke-mimic differentiation.

## 2. Methods

### 2.1 Search strategy and gatering of data

This umbrella review has been conducted in accordance with the principles of the Preferred Reporting Items for Systematic Reviews and Meta-Analyses (PRISMA) 2020 statement^15^. The full checklist is available in the supplementary material.

We conducted electronic searches for eligible studies within each of the following databases, from inception up to May 10th, 2025: MEDLINE (accessed via PubMed), Epistemonikos, and the Cochrane Database of Systematic Reviews. Records were identified in each database with tailored search strings and filters, which can be found in the supplementary material.

We considered systematic review and meta-analysis that included studies:

- On fluid biomarkers used to differentiate ischemic stroke from stroke mimics or controls
- With fluids sampling < 72h
- Including a confirmatory MRI imaging with diffusion sequences, as standard of truth
- On human subjects
- In English, German, Dutch, Italian, French, Spanish, Portuguese

Exclusion criteria comprised:

- Studies comparing ischemic stroke vs hemorrhagic stroke
- Systematic reviews not reporting differences in biomarkers levels between AIS and stroke mimics
- Systematic reviews or meta-analysis which did not report data from primary studies
- Primary studies
- Narrative reviews
- Conference abstracts

Citations identified from the literature searches were imported to Rayyan (https://rayyan.ai/) and duplicates were automatically identified by the software and manually checked before removal. Two reviewers (AB and FEP) then independently screened the titles and abstracts of all records. In case of disagreement about eligibility, a consensus was reached through discussion with all the authors. Full texts of all potentially eligible studies, systematic reviews and meta-analyses were retrieved, and reference lists were checked to identify additional eligible studies and to elucidate theoretical aspects in the discussion.

Data extraction from eligible studies was performed by all authors independently. We extracted data for quantitative analysis extending the dataset required for the R package *metaumbrella* (REF GOSLING), including: meta-analysis, and for each primary study satisfying inclusion and exclusion criteria, biomarker, author and year, presence of multiple effect sizes, type of effect size (either mean difference (MD) or standardized mean difference (SMD), when applicable), estimate and 95% CI of the effect size, number of cases, number of controls, risk of bias (low, unclear, high, recoding what was done in the original systemic review if needed), time from onset, type of biomarker, medium, definition of controls, direction of the difference (significantly higher in stroke, no difference, significantly lower in stroke, mixed results – the latter meaning that directions varied according to subgroups or timepoints). In case of absence of MD or SMD, data from correspondent studies were summarized narratively. For this reason, transcriptomic data were not meta-analyzed.

### 2.2 Transcriptomic analysis

We identified 391 miRNAs, alongside 46 circRNAs, and 55 lncRNAs. We have harmonised nomenclature and rigorously eliminated duplicates using miRbase (https://www.mirbase.org/), circBase (https://www.circbase.org/), and GENCODE (https://www.gencodegenes.org/) identifiers. After this process, the catalogue comprises: 220 distinct miRNAs, 45 unique circRNAs, and 44 nonredundant lncRNAs.

### 2.3 Statistical analysis

We conducted a meta-analysis of pooled SMD of primary studies, after exclusion of duplicates. A random effect model was used. In cases in which only median-based measures were provided, we converted those to means and standard devation with the method described in Shi et al 202116. The package *metaumbrella* in R 4.3.0 was used to run the analyses and generate forest plots17. The excess of significance bias test and the Egger’s tests were performed to evaluate publication bias. Heterogeneity was estimated with I2. The class of the evidence was computed with the method proposed by Ioannidis18. We ran separate analyses restricting the meta-analysis to studies performed within therapeutic time-windows, i.e. 4.5 hours (thrombolysis), 6 hours (mechanical thrombectomy), 9 hours (extended thrombolysis), 24 hours (extended thrombectomy).

To visualize the qualitative data, we devised a new type of plot called an arrow plot, which graphically represents both the main direction(s) of the biomarker effects and the quality of the evidence. Each line in the plot summarizes the systematic review results for one biomarker. Specifically:

- Blue arrows pointing left represent biomarkers that were significantly lower in ischemic stroke patients compared to controls.
- Gray lines represent biomarkers with no significant difference.
- Red arrows pointing right represent biomarkers that were significantly higher in controls.

The length of each arrow corresponds to the total number of patients and controls for which the outcome (lower, no difference, or higher) was observed in the primary studies. The opacity of each line reflects the average risk of bias across these studies: the more transparent the line, the higher the risk of bias.

Duplicates of primary studies were removed for the analysis by biomarker. If duplicated primary studies had conflicting risk of bias assessments, we used the highest risk of bias value to adopt a conservative approach.

## 3. Results

We identified 910 studies across three databases; after duplicate removal, and title and abstract screening 49 studies were retrieved and full-text checked for inclusion and exclusion criteria. Finally, 27 systematic reviews were included in the current umbrella review. The flow-chart of the umbrella review is presented in **Figure 1**.

**Figure 1.**
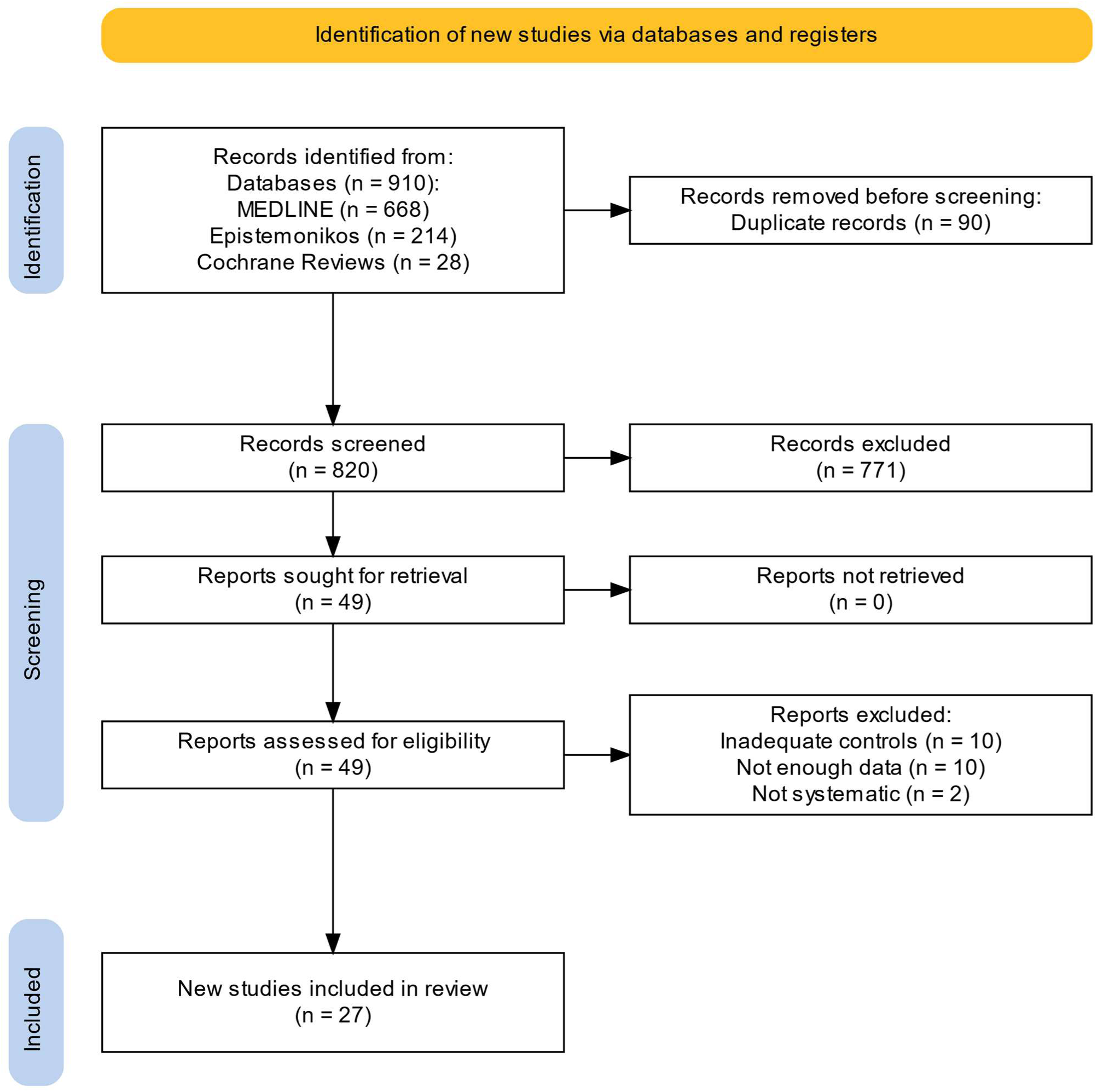
Prisma flowchart.

Biomarkers were categorized as follow: biochemistry, metabolomics, transcriptomic (included microRNAs [miRNAs], circular RNAs [circRNAs], and long noncoding RNAs [lncRNAs]), and cell-free DNA (cfDNA). Summary characteristics of the included meta-analyses for each category are presented in **Tables S1-S8** in the supplementary material.

### 3.1 Biochemistry biomarkers

Fourteen studies reported data on biochemistry biomarkers in blood or CSF^19–32^. The results of the meta-analysis of the primary studies showed significant differences between patients with ischemic stroke and controls in several molecules. NSE showed the greatest effect size (estimated SMD (eG) 2.61), followed by ischemia-modified albumin (IMA, 2.54), d-Dimer (2.01), S100B (1.63), copeptin (1.36), BDNF (1.28), IL-6 (1.20), NfL (0.77), neutrophiles (0.77), C-reactive protein (CRP, 0.70), GFAP (0.48), matrix-metalloproteinase 9 (MMP-9, 0.46), HbA1c (0.45), BNP (0.41), creatinine (0.40), glucose (0.40), albumin (-0.35), leukocytes (0.34), blood urea nitrogen (BUN, 0.25). However, for several of these biomarkers the meta-analysis revealed high heterogeneity across studies (with I2 > 80% for BDNF, Copeptin, CRP, d-Dimer, GFAP, IL-6, IMA, Leukocytes, S100B) and positive excess significance bias (BNP, CRP, d-Dimer, IL-6). The Egger’s test suggested publication bias for CRP, d-Dimer, HbA1c, IMA, IL-6, S100B. The forest plot of the meta-analyses is shown in **Figure 2**.

**Figure 2.**
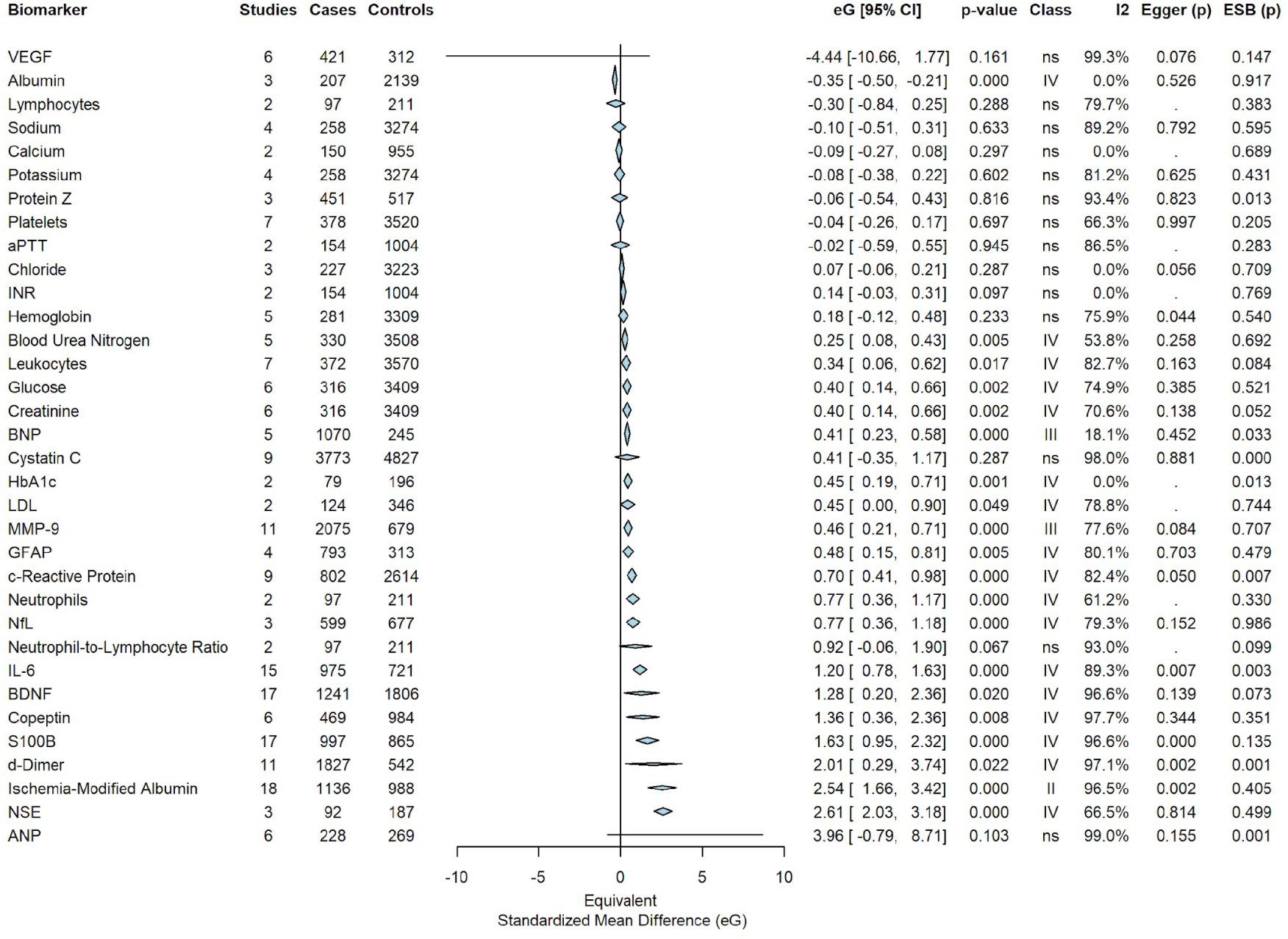
Meta-analyses for each factor, all time-points.

#### 3.1.1 Meta-analysis considering usual window for acute therapeutic procedures

We restricted these analyses to studies occurring in the time window for acute therapeutic procedures. Quantitative data were available for only three biomarkers within the 4.5 hours for intravenous thrombolysis, coming from one study each. IL-6 and GFAP showed significantly positive effect sizes, while a nearly significant difference was noted for d-Dimer. Within 6 hours, the usual time window for mechanical thrombectomy, d-Dimer was the only significant biomarker with at least two studies (eG 0.45); quantitative data from single studies showed that lymphocytes were significantly lower in stroke patients, while creatinine, CRP, IL-6, leucocytes, NSE neutrophiles, neutrophiles-to-lymphocytes ratio and S100B were significantly elevated in stroke.

#### 3.1.2. Meta-analysis considering extended window for acute therapeutic procedures

We then considered the extended time window for such procedures. For extended thrombolysis (within 9 h), the highest eG was noted for NSE (2.89), followed by S100B (2.82) and d-Dimer 0.45 across biomarkers with at least two studies each; S100B data showed high heterogeneity (I2 > 80%). Within the time window for extended thrombectomy (24 h), several biomarkers were evaluated across multiple studies. The highest significant eG was reached by NSE (2.89), followed by IMA (2.54), S100B (2.11), d-Dimer (1.89), IL-6 (1.28), GFAP (0.48), MMP9 (0.46), BNP (0.41). Platelets were significantly elevated in stroke patients compared to controls in one study. The relative forest plots are shown in **Figures S1-S4** in the supplementary material.

#### 3.1.3 Summary of the systematic review

Looking at the overall data derived by the summary of the systematic review, the number of non-duplicated primary studies for each biomarker for each prevalent direction and timing are presented in **Table 1**; **Table S1** in the supplementary material adds the risk of bias and the medium in which the biomarkers have been analyzed. In **Figure 3** we summarized using an arrow plot all the qualitative results of the systematic review. Among biomarkers with at least two studies, a few showed quite consistent prevalent direction (> 50% of involved patients across primary studies): aPTT, BUN, CRP, copeptin, creatinine, d-Dimer, e-Selectine, glucose, HbA1c, ICAM-1, IL-6, IMA, l-Selectin, LDL, neutrophiles-to-lymphocytes ratio, neutrophiles, NfL, NSE, p-Selectin, S100B, VCAM-1 and TNF-α were elevated in stroke compared to controls.

**Figure 3.**
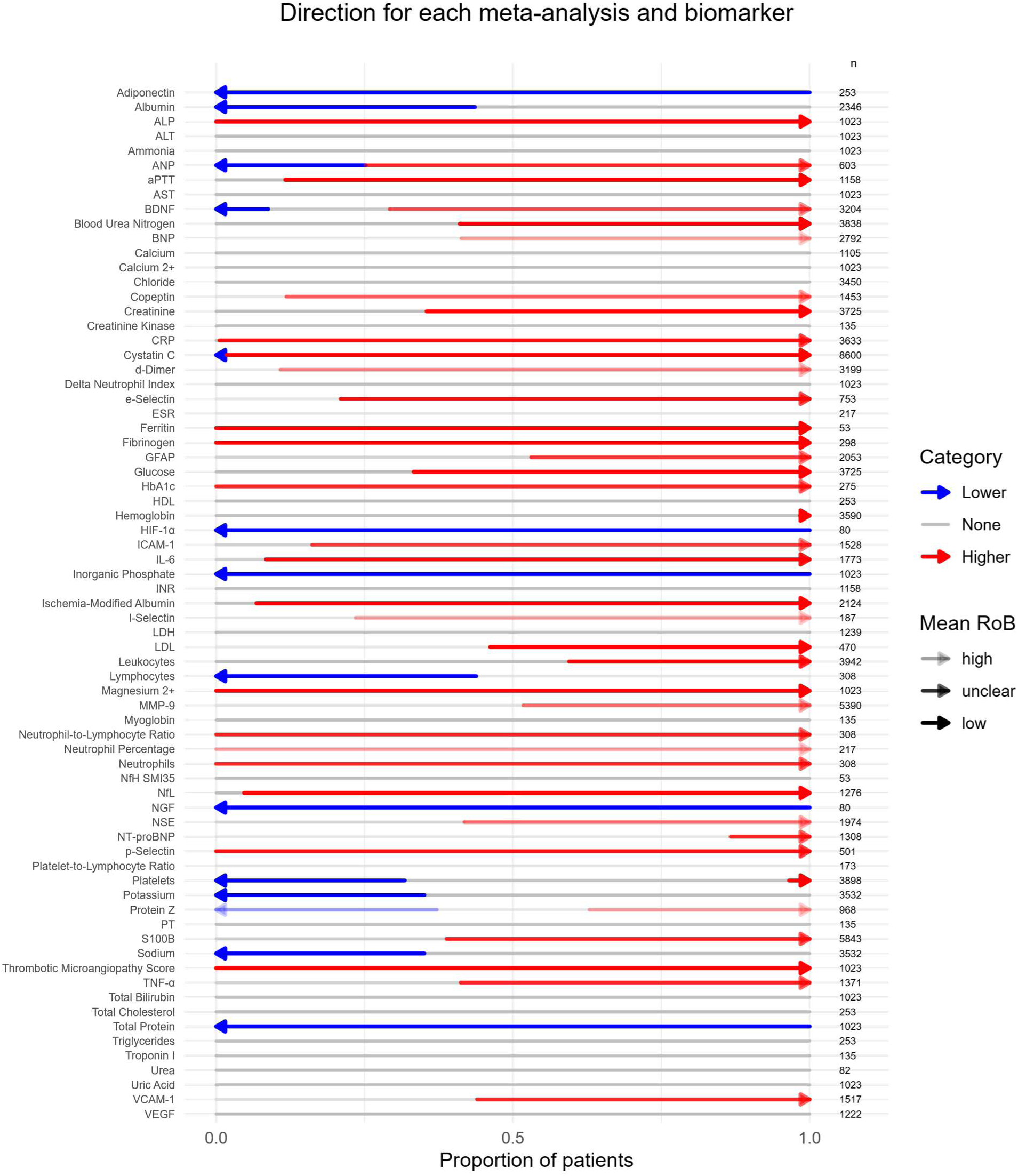
Summary of the systematic review: direction of biochemistry biomarkers (all primary studies combined)

**Table 1.**
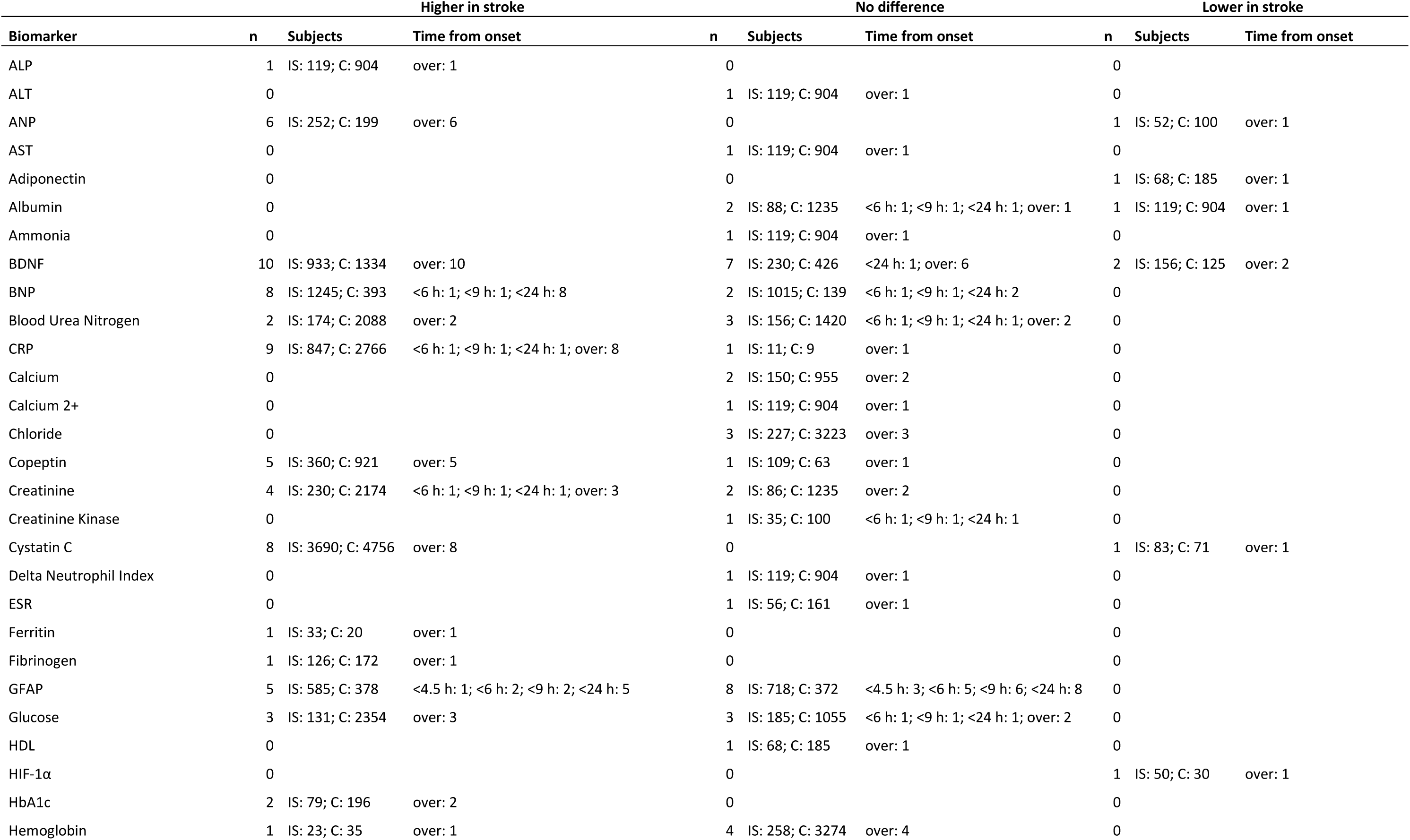
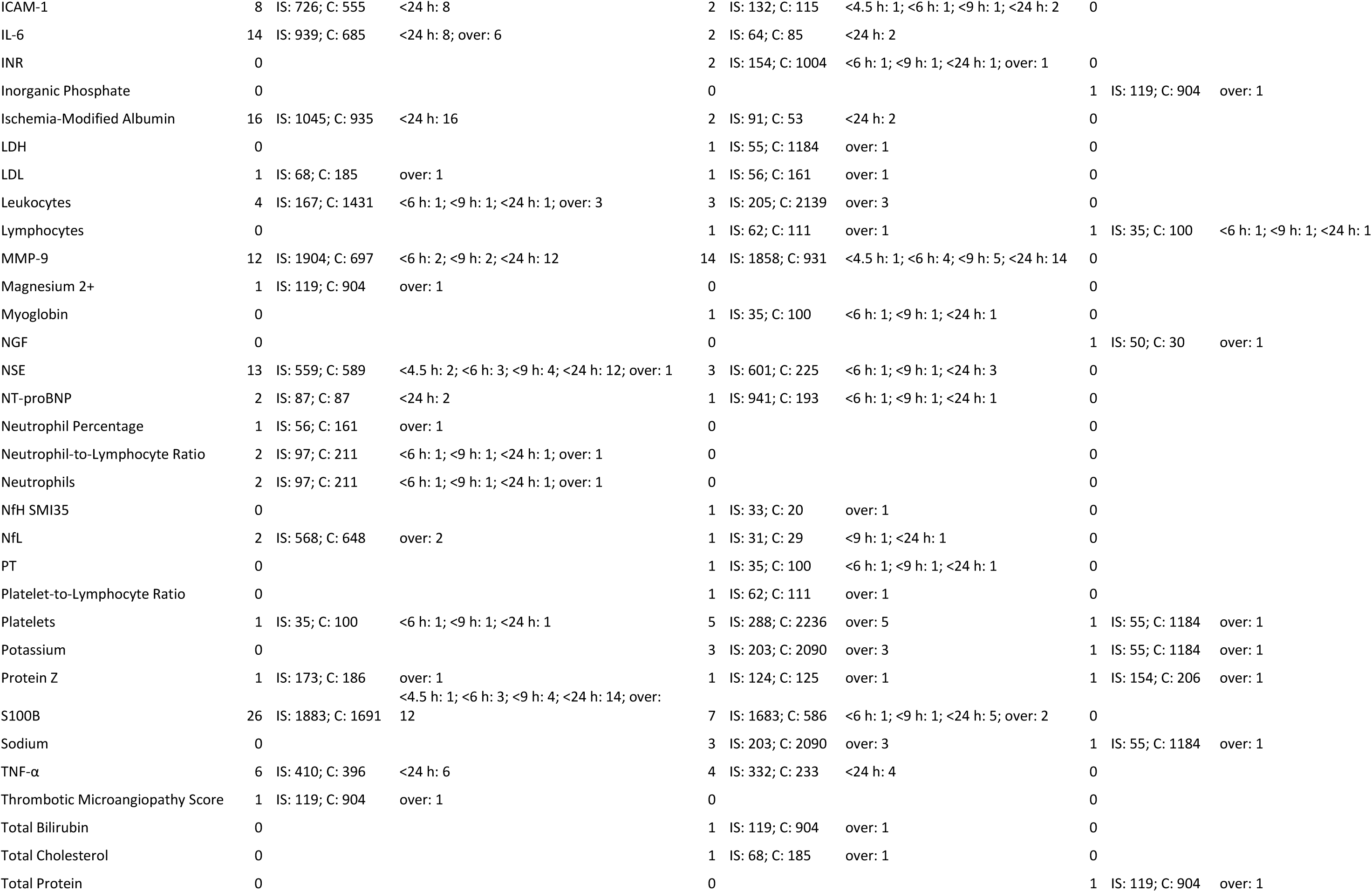
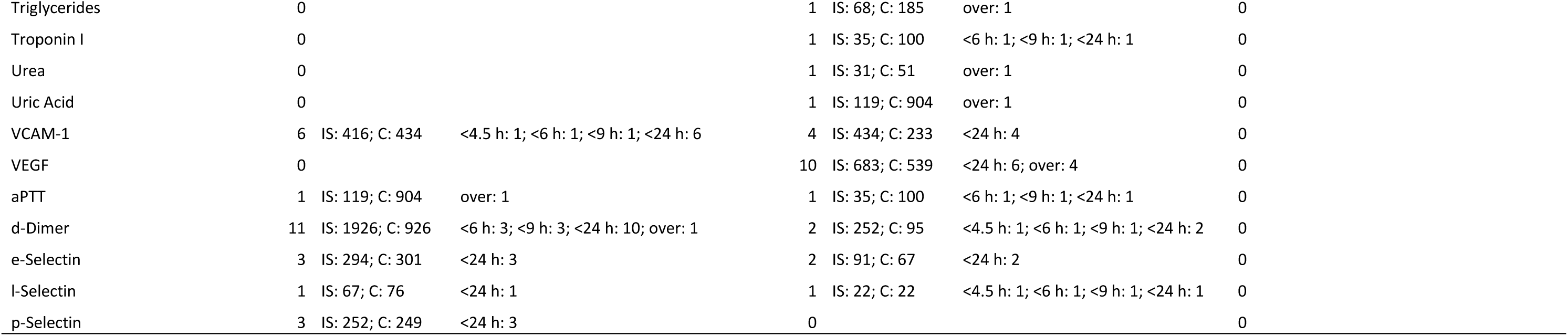
Directions of biomarkers (pooled data from all primary studies with exclusion of duplicates): biochemistry.

The arrow plots for the usual therapeutic time windows are presented in **Figure 4**. Within 4.5 hours, a number of biomarkers were quite consistently elevated in stroke patients vs controls (> 50 of involved patients across primary studies): d-Dimer, ICAM-1, IL-6, l-Selectin, NSE, S100B, VCAM1. Within 6 hours, besides these biomarkers, also aPTT, BUN, BNP, creatinine, d-Dimer, glucose and NSE were elevated in stroke compared to controls (CRP was higher in the only included study).

**Figure 4.**
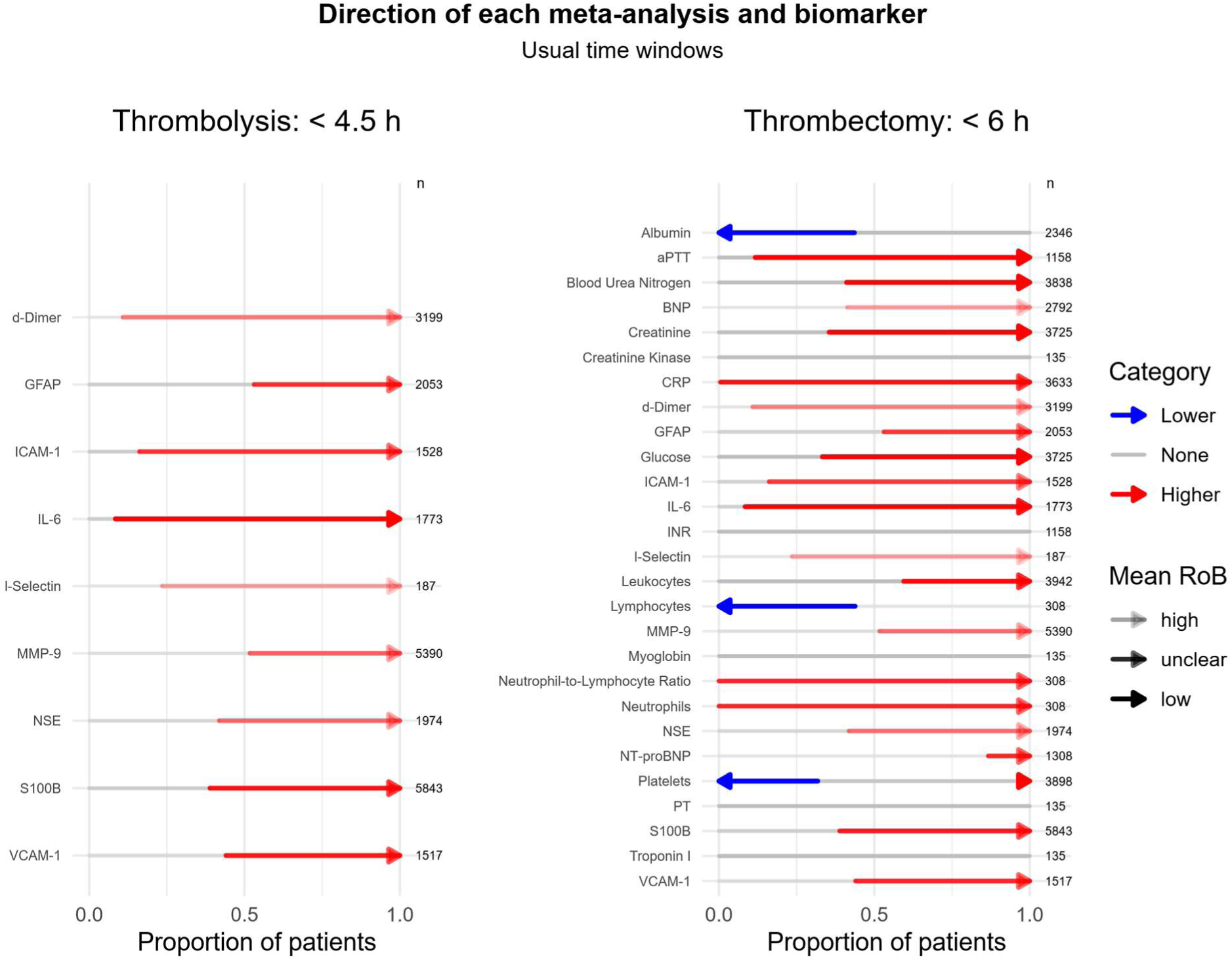
Summary of the systematic review: direction of biochemistry biomarkers (all primary studies combined) in the usual time-window.

The arrow plots for the extended therapeutic time windows are presented in **Figure 5**. Within 9 hours, in addition to the elevated biomarkers at 6 hours, CRP and NfL were elevated in stroke compared to controls. Within 24 hours, also e-Selectin, IMA, p-Selectin, TNF-α were more elevate in stroke.

**Figure 5.**
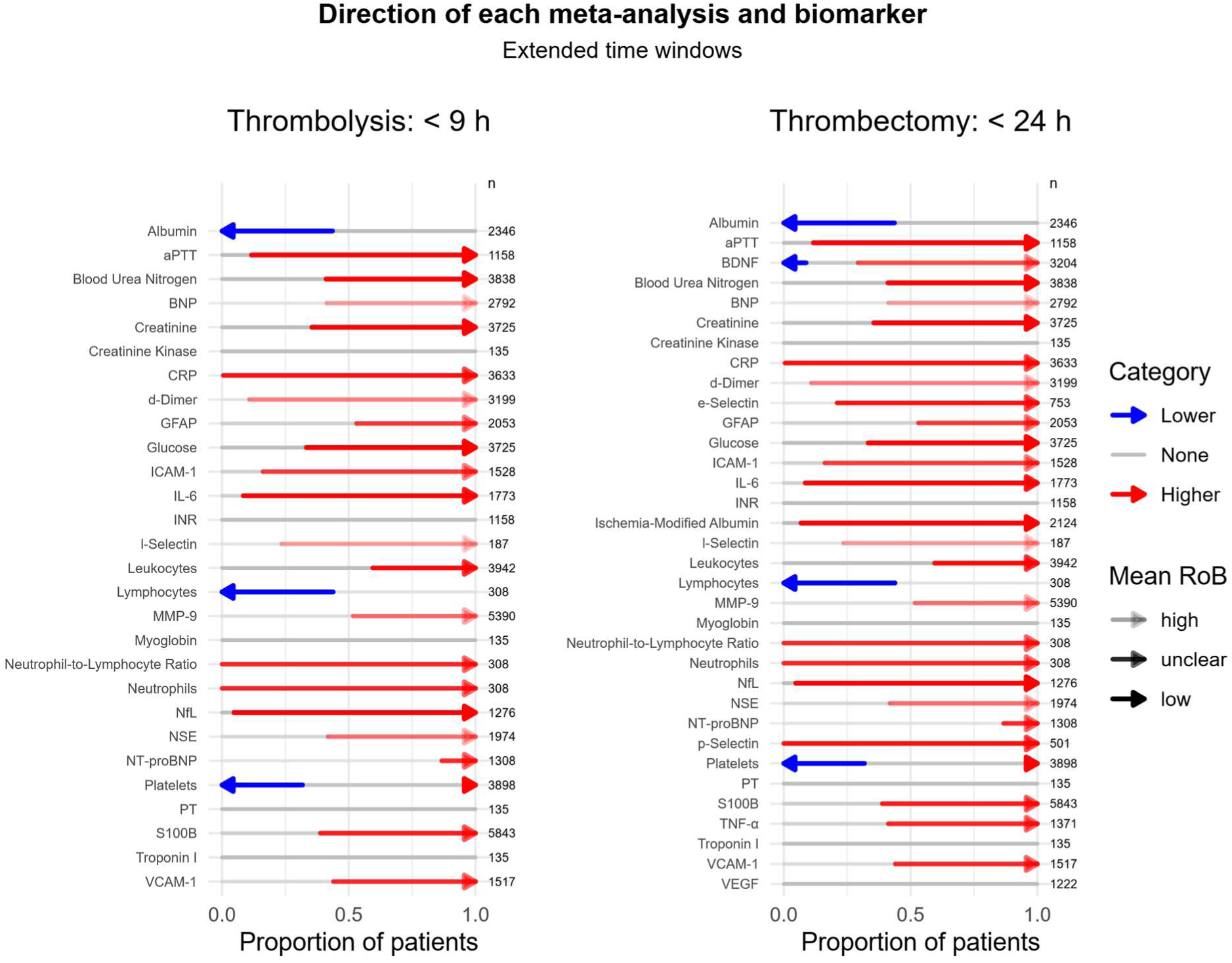
Summary of the systematic review: direction of biochemistry biomarkers (all primary studies combined) in the extended time-window.

### 3.2 Metabolomics biomarkers

Ke and colleagues conducted the sole systematic review of metabolomic signatures in AIS, examining 28 studies published between 2000 and 2019^33^. Their qualitative synthesis highlighted a consistent pattern of metabolic perturbation within hours of cerebral ischemia. Key elevations were observed in lactate and succinate, markers of anaerobic glycolysis, and glutamate, reflecting excitotoxic stress. Lipid-derived mediators, most prominently LysoPC(16:0) and LysoPC(18:2), were also increased. Among amino acids, branched-chain species (leucine, isoleucine, valine) and free fatty acids rose alongside uric acid, signalling oxidative stress and cell membrane turnover. In contrast, arginine and citrulline levels declined, implicating disruption of the nitric oxide synthesis pathway, while citrate depletion underscored tricarboxylic-acid-cycle failure. The authors advocated targeted prospective validation of these candidates, ideally integrated with proteomic and transcriptomic datasets, to develop a truly multimodal diagnostic panel.

### 3.3 Transcriptomics biomarkers

The data from 126 primary studies included in 10 systematic reviews are presented in **Tables S2-S4** in the Supplementary material.

#### 3.3.1 MiRNAs

Across eight high-quality systematic reviews^14,20,34–39^, 220 records described changes in circulating miRNAs levels within the first 24 hours of AIS, most often quantified by RT-qPCR in plasma or serum (**Table S2**).

Of the 220 miRNAs, **132** were found to be up-regulated and **108** down-regulated. The most promising and robust miRNAs increased in at least two separate investigations and evaluated in large cohorts included:

- **miR-16**: up-regulated in 384 AIS vs 2531 HCs across 6 primary studies (Tian 2016, Tan 2009, Wu 2015, Salinas 2019, Rainer 2016, Leung 2014). In these studies, its dysregulation has been linked to BCL2-mediated apoptosis.
- **let-7e-5p:** up-regulated in 4 primary studies (Gui 2019, Huang 2016, Yoo 2019, Peng 2015) including 516 AIS and 420 HCs. Its increase may represent a compensatory attenuation of sterile inflammation by silencing HMGA2 and repressing TLR4–MyD88–NF-κB signalling.
- **miR-107**: consistently up-regulated in a total of 161 AIS and 102 HCs in 3 primary studies (Tian 2016, Yang 2016, Sørensen 2017), which is linked to oxidative-stress and endothelial dysfunction pathways.
- **miR-451a**: up-regulated in 182 AIS vs 198 HCs by Li P. (2015), Giordano (2019) and Aldous (2022), each emphasizing its role in dampening neuroinflammation.

Conversely, the most down-regulated species of miRNAs in at least two primary studies and evaluated in more than 50 AIS and 50 HCs are:

- **miR-146a-5p**: lower levels in AIS reported by Li S. (2015) and Kotb (2019), in a total of 104 AIS and 52 HC. It is implicated in failure of its anti-inflammatory feedback on toll-like receptor (TLR) signaling.
- **let-7d-3p:** lower levels in AIS reported by Salinas 2019 and Sepramaniam 2014, in a total of 249 AIS and 2384 HC. Its suppression lifts repression of RAS/MAPK and IL-6 pathways, amplifying ERK-mediated cytokine release and neuronal injury.
- **miR-142-3p**: down-regulated in 57 AIS and 107 HCs reported in two studies (Yoo 2019 and Aldous 2022). Its down-regulation release inhibition of SMAD3, enhancing TGF-β-driven BBB disruption and leukocyte infiltration.

Moreover, a handful of miRNAs show a mixed-effect, being reduced in most of the studies, and increased in other studies. The most relevant included in this group are: **miR-185-5p**, implicated it in apoptotic and inflammatory cascades, **miR-130a-3p**, consistent links to VEGF signaling and blood– brain-barrier disruption, **miR-124-3p**, whose decline was associated with unchecked excitotoxicity and NF-κB activation, **miR-126-3p**, playing a protective role in PI3K–Akt-mediated endothelial repair.

Three comprehensive systematic reviews^14,20,37^ have also documented the dynamic temporal regulation of specific miRNAs over the first three days post-onset. In particular, miR-30a was shown to be upregulated at 6 hours but inversely downregulated by 72 h, suggesting a shift from early stress response to recovery phase signaling. Meanwhile, miR-124 maintained persistent downregulation at 24, 48 and 72 hours, correlating with ongoing neuronal loss. Interestingly, fragments derived from transfer RNAs rose steadily from 24 hours through day 7, pointing to a sustained stress response, while miR-503 peaked transiently at 6–24 h before returning towards baseline^38,39^.

#### 3.3.2 Circular RNAs

Three systematic reviews^14,37,40^ collated 46 circRNA entries derived from case–control and cohort studies within 24 hours of AIS (**Table S3**).

After nomenclature unification, 21 circRNAs were consistently reported as upregulated, most notably circHECTD1 and circCDC14A, each appearing in all three reviews. Additional upregulated circRNAs included circFUNDC1, circTLK1, circRBM33 and circZCCHC11, with fold of changes ranging from 1.8 to 3.2 times those of healthy controls.

Conversely, 20 circRNAs were downregulated, among which circDLGAP4 and hsa_circ_0001599 were most commonly cited.

Functional enrichment analyses across these reviews repeatedly implicated PI3K–Akt, MAPK and NF-κB signalling pathways, as well as processes governing neuroinflammation and blood–brain barrier integrity.

#### 3.3.3 Long Noncoding RNAs

Two systematic reviews^14,41^ summarised 55 lncRNA records, all assayed within the first 24 hours of symptom onset (**Table S4**). From these, approximately 40 were reported as upregulated, among them MALAT1 and HOTAIR, both of which exhibited fold-changes of 1.7–2.2× in small patient cohorts. Conversely, around 15 lncRNAs were found to be downregulated, with LINK-A and MEG3 emerging as the most promising in differentiating AIS from non-stroke controls. Although these findings are compelling, the generally modest sample sizes (typically fewer than 60 subjects per study) and paucity of stroke mimics comparisons temper conclusions about specificity and underline the need to perform larger multicenter studies.

### 3.4 Cell-Free DNA

Five primary studies (Tsai 2011, Bustamante 2016, O’Connell 2017, Valles 2017, and Vasilyeva 2020) relating to cfDNA were synthesized in the Zhang et al. review^14^, all focusing on blood collections within six hours of AIS (**Table S5**). Five unique cfDNA species were identified, including total nuclear cfDNA markers such as B-globin and TERT, which were elevated in stroke patients versus controls. Mitochondrial cfDNA species (for example MT-ND2) exhibited even greater relative increases (between 2.5- and 3.5-fold) and in several studies directly correlated strongly with admission NIHSS scores. Smaller low-molecular-weight fragments also rose markedly. However, varied DNA extraction protocols (phenol–chloroform versus silica-column kits) and differences in qPCR calibration kits introduced considerable methodological heterogeneity, precluding formal meta-analytic pooling of effect sizes.

## 4. Discussion

This umbrella review represents the most comprehensive synthesis to date of early fluid biomarkers in AIS, spanning biochemistry, metabolomics and multiple layers of nucleic acid regulation. They chart the molecular evolution of AIS and show a clinical potentiality in rapid diagnosis, in prognostication and as therapeutic guidance.

### 4.1 Biochemistry Biomarkers and Barrier Disruption Biomarkers

The temporal evolution of proteomic and cellular biomarkers in AIS reveals a striking choreography of endothelial, inflammatory and necrotic processes unfolding within the first 24 hours of symptom onset.

#### Hyperacute Phase (≤ 4.5 h)

The hyperacute phase of AIS (≤ 4.5 h), coinciding with current windows for intravenous thrombolysis, is dominated by markers of coagulopathy and gliovascular injury:

- d-Dimer reflects acute fibrin turnover and microthrombus resolution. Mechanistically, fibrin degradation products engage endothelial PAR-1, amplifying NF-κB-dependent IL-6 release and transiently increasing vascular permeability through VE-cadherin phosphorylation.
- GFAP signals immediate astrocytic cytoskeletal disruption. Extravasated GFAP binds TLR4 on microglia, driving NLRP3 inflammasome assembly and IL-1β secretion^42,43^. Although it’s relative rapid increase in AIS, it is also increased in hemorrhagic stroke, reducing the utility of this biomarker^44^.
- IL-6 is secreted by activated microglia and peripheral monocytes, triggering JAK/STAT3-mediated acute-phase responses and upregulating ICAM-1/VCAM-1 to recruit neutrophils across the blood–brain barrier.

#### Acute Phase (≤ 24 h)

Within the first 24 hours, a broader array of proteins and cells resulted to be increased in AIS compared to mimics. They mainly reflect the consolidation of necrosis, inflammation and extracellular-matrix remodeling:

- **NSE** exhibits the largest effect, a direct marker of neuronal necrosis.
- **IMA** rises markedly, reflecting oxidative N-terminal alteration that promotes peroxynitrite formation and endothelial nitrosative injury.
- **S100B** is secreted by reactive astrocytes and engages RAGE on glia and endothelium, perpetuating NF-κB-driven inflammation.
- **MMP-9** degrades basal lamina collagen IV, exacerbating vasogenic oedema and risk of haemorrhagic transformation.
- **BNP** signifies neuro-cardiac cross-talk; acute sympathetic surge releases BNP which, while vasodilatory, correlates with poorer outcome via maladaptive fluid shifts.
- **CRP** completes the acute-phase profile, binding FcγR on microglia to amplify phagocytic activation.

Moreover, also expanding the sampling window to 9- and 24-hours post-onset and including additional studies, GFAP and d-Dimer were confirmed to be significantly increased in AIS compared to mimics and HC.

Another notable data concerns white blood cells changes: lymphocyte counts fall by six hours, reflecting immediate stroke-induced immunosuppression, while neutrophils rise, contributing to early oxidative and proteolytic damage via myeloperoxidase and elastase release.

Very few data exist on the dynamic changes of these biomarkers during the first hours of AIS, limiting our understanding of their true hyperacute dynamics and the optimal time for sampling in pre-hospital settings.

Considering these proteomic and cellular changes, most important alterations involve: BBB function, Inflammation pathways, Coagulation–Fibrinolysis Balance and Neuronal survival pathways.

By mapping these alterations onto canonical pathways (TLR4/NF-κB, JAK/STAT3, PAR-1, MMP-mediated matrix remodeling and RAGE signaling) we delineate the multi-layered pathobiology of hyperacute AIS.

### 4.2 Metabolomic Shifts and Energetic Collapse

Metabolomic profiling highlights the accumulation of lactate and succinate, hallmarks of anaerobic glycolysis, alongside elevated glutamate, which underpins excitotoxic neuronal death (Ke et al. 2019). Succinate, in particular, functions as a danger-associated molecular pattern (DAMP), stabilising hypoxia-inducible factor-1α (HIF-1α) and driving IL-1β production via HIF-1α and TLR4 signalling in immune cells^45^. This succinate-HIF-1α-IL-1β axis not only links metabolic collapse to sterile inflammation but also suggests that modulating succinate levels could temper cytokine surges in AIS. Moreover, depletion of arginine and citrulline impairs nitric oxide (NO) synthesis, exacerbating vascular dysfunction.

### 4.3 Transcriptomic Regulators of Survival and Inflammation

Our analysis confirmed that certain miRNAs are robustly and reproducibly dysregulated within the first 24 hours of AIS. miR-16-5p, let-7e-5p, miR-107 and miR-451a consistently rose across three or more independent cohorts (e.g. Tian 2016; Huang 2016; Li 2015; Giordano 2019; Aldous 2022), underlining their putative roles in apoptosis regulation (via BCL2), oxidative-stress responses and modulation of neuroinflammation. By contrast, miR-126-3p and miR-124-3p demonstrated uniform suppression in multiple studies (Tan 2009; Long 2013; Jin 2017; Liu 2015; Zeng 2015; Sun 2019), consistent with loss of endothelial-protective and neuronal-survival signaling (PI3K/Akt/eNOS and NF-κB pathways, respectively).

However, a subset of miRNAs, including miR-130a-3p, miR-185-5p, miR-30a-5p and miR-9-5p, exhibited mixed directionality (up in some cohorts, down in others), often in time-dependent fashion (e.g. miR-30a-5p initial rise at 6 h then fall by 72 h). Such discrepancies likely reflect dynamic temporal kinetics, cohort heterogeneity (stroke severity, comorbidity), and methodological variables (RNA extraction, platform sensitivity). Future studies must standardize sampling intervals (e.g. hyperacute ≤ 6 h vs early acute 6–24 h) and assay protocols to reconcile these conflicting findings.

CircRNAs (circHECTD1, circCDC14A) and lncRNAs (lncRNA H19, ANRIL, LINK-A, MEG3) further diversify the landscape of AIS biomarkers. circHECTD1’s elevation, observed in all three meta-analyses, supports its role in mTOR-mediated autophagy and reactive astrocytosis, while the consistent down-regulation of circDLGAP4 implicates loss of MAPK/ERK-mediated endothelial barrier protection. Among lncRNAs, LINK-A and MEG3 achieved high diagnostic accuracy (AUCs > 0.85) despite small sample sizes, suggesting that these transcripts act as master regulators of chromatin state and cell survival. Nonetheless, modest cohort sizes and limited mimic comparisons highlight the need for larger, multicenter validation.

### 4.4 Cell-Free DNA as a DAMP mediator

Elevated cfDNA, particularly mitochondrial MT-ND2 fragments, was uniformly reported across five primary studies within six hours of AIS. Beyond its value as a passive marker of necrosis, mt-cfDNA engages endosomal TLR9 on microglia to trigger type I interferon and NLRP3 inflammasome activation, thus serving as an active danger-associated molecular pattern (DAMP)^42,46^. The amplitude of cfDNA release correlates with NIHSS scores, suggesting that cfDNA quantification could both diagnose AIS and gauge stroke severity. Standardizing pre-analytical protocols (extraction method, fragment sizing) will be crucial for cfDNA’s adoption in clinical settings.

### 4.5 Integrative Pathway Network

Collectively, these biochemistry, metabolomic and transcriptomic layers converge on a few nodal axes:

1. PI3K/Akt/eNOS: regulated by miR-126, miR-124 and succinate-HIF-1α signaling, this axis governs endothelial survival and barrier function.
2. NF-κB/JAK/STAT: amplified by GFAP-driven DAMP signaling, S100B via TLR4/RAGE interaction and cfDNA-TLR9, orchestrating cytokine storms.
3. MAPK/ERK and mTOR/autophagy: modulated by circDLGAP4 and circHECTD1, balancing cell survival and glial scarring.
4. Metabolic reprogramming: succinate accumulation under hypoxia stabilizes HIF-1α, shifting immune cells towards glycolytic, pro-inflammatory phenotypes while depleting arginine disrupts NO-mediated vasodilation.

### 4.6 Clinical and Translational Implications

This integrated mechanistic framework suggests that a multi-analyte panel, combining GFAP, S100B, d-Dimer, key metabolites (succinate, lactate), cfDNA markers and a tailored miRNA/lncRNA/circRNA signature, could be applied in a clinical setting in a near future. This could be fundamental in: A) differentiating AIS from mimics in pre-hospital or emergency settings with high sensitivity and specificity; B) stratifying patients by predominant pathophysiological endotype (coagulopathic vs inflammatory vs metabolic); C) informing personalized therapeutic decisions (e.g. tailored anti-inflammatory or anti-fibrinolytic strategies), and monitoring treatment response and predicting secondary complications (e.g. hemorrhagic transformation via MMP-9 dynamics).

### 4.7 Strengths, Limitations and Future Directions

Our work has some limitations. Drawing conclusions from different meta-analysis and several primary studies necessarily entails dealing with heterogeneity in assay platforms, modest sample sizes (for instance for non-coding RNA studies) and variability in control definitions. Nevertheless, our umbrella review has several strengths. It includes a comprehensive multi-omic coverage, with rigorous harmonization of nomenclature and temporal stratification across clinically meaningful windows. Moreover, our wide approach allows us to define the biochemical landscape of AIS. This broader time frame underscores an important gap: very few data exist on the hour-to-hour kinetics of these biomarkers in the first hours of AIS, limiting our understanding of their true hyperacute dynamics and the optimal time for sampling in pre-hospital settings.

Future research must prioritize standardized protocols, absolute quantification of transcriptomic markers (e.g. digital PCR), refine temporal profiling to capture minute-to-minute biomarker kinetics, develop point-of-care devices (microfluidic immunoassays, isothermal amplification) capable of multiplexed, rapid (≤ 20 min) readouts, and large-scale, multi-center validation against stroke mimics. Additionally, therapeutic modulation of key nodes warrants exploration in preclinical and early-phase clinical trials. Crucially, standardizing sampling windows and analytical platforms will be essential to mitigate temporal and methodological heterogeneity.

By bridging mechanistic insight with clinical pragmatism, this multi-omic framework lays the groundwork for next-generation, fluid-based diagnostics and prognostic that can narrow treatment delays and improve outcomes in AIS.

## 5 Conclusions

In conclusion, our umbrella review demonstrates that several fluid biomarkers, including proteins, RNAs (circRNAs, miRNA, lncRNA), cfDNA, and metabolic indicators, can discriminate AIS from mimics and HCs within the hyperacute window. Together, this omic approach constitutes a compelling framework for point-of-care diagnostics that may transform hyperacute stroke triage and management. Prospective standardization of sampling protocols and validation in pre-hospital cohorts will be essential to translate these findings into rapid diagnostic assays.

## Supporting information

Supplementary Material_Strings

## Data Availability

All data produced in the present work are contained in the manuscript and in the supplementary data.

## Declarations

### Ethical Statement

No informed consent nor ethical committee agreement was required since it is a metanalysis.

## References

1. Hilkens NA, Casolla B, Leung TW, de Leeuw FE. Stroke. The Lancet. 2024;403(10446):2820–2836. doi:10.1016/S0140-6736(24)00642-1

2. Tissue plasminogen activator for acute ischemic stroke. The New England journal of medicine. 1995;333(24):1581–1588. doi:10.1056/NEJM199512143332401

3. Hacke W, Kaste M, Bluhmki E, et al. Thrombolysis with Alteplase 3 to 4.5 Hours after Acute Ischemic Stroke. New England Journal of Medicine. 2008;359(13):1317–1329. doi:10.1056/NEJMOA0804656,

4. Berkhemer OA, Fransen PSS, Beumer D, et al. A Randomized Trial of Intraarterial Treatment for Acute Ischemic Stroke. New England Journal of Medicine. 2015;372(1):11–20. doi:10.1056/NEJMOA1411587,

5. Goyal M, Demchuk AM, Menon BK, et al. Randomized Assessment of Rapid Endovascular Treatment of Ischemic Stroke. New England Journal of Medicine. 2015;372(11):1019–1030. doi:10.1056/NEJMOA1414905,

6. Campbell BCV, Mitchell PJ, Kleinig TJ, et al. Endovascular Therapy for Ischemic Stroke with Perfusion-Imaging Selection. New England Journal of Medicine. 2015;372(11):1009–1018. doi:10.1056/NEJMOA1414792,

7. Saver JL, Goyal M, Bonafe A, et al. Stent-Retriever Thrombectomy after Intravenous t-PA vs. t-PA Alone in Stroke. New England Journal of Medicine. 2015;372(24):2285–2295. doi:10.1056/NEJMOA1415061,

8. Purroy F, Farré-Rodriguez J, Mauri-Capdevila G, Vicente-Pascual M, Farré J. Basal IL-6 and S100b levels are associated with infarct volume. Acta Neurologica Scandinavica. 2021;144(5):517–523. doi:10.1111/ANE.13487,

9. Chen X, Li S, Chen W, et al. The Potential Value of D-Dimer to Fibrinogen Ratio in Diagnosis of Acute Ischemic Stroke. Journal of Stroke and Cerebrovascular Diseases. 2020;29(8). doi:10.1016/j.jstrokecerebrovasdis.2020.104918

10. Cabezas JA, Bustamante A, Giannini N, et al. Discriminative value of glial fibrillar acidic protein (GFAP) as a diagnostic tool in acute stroke. Individual patient data meta-Analysis. Journal of Investigative Medicine. 2020;68(8):1379–1385. doi:10.1136/JIM-2020-001432,

11. Onatsu J, Vanninen R, JÄaKÄaLÄa P, et al. Tau, S100B and NSE as blood biomarkers in acute cerebrovascular events. In Vivo. 2020;34(5):2577–2586. doi:10.21873/INVIVO.12075,

12. Camara-Lemarroy CR, Escobedo-Zúñiga N, Guzmán-de la Garza FJ, Castro-Garza J, Vargas-Villarreal J, Góngora-Rivera F. d-Lactate and intestinal fatty acid-binding protein are elevated in serum in patients with acute ischemic stroke. Acta Neurologica Belgica. 2021;121(1):87–93. doi:10.1007/S13760-018-0940-X,

13. Qi B, Zhang Y, Xu B, et al. Metabolomic Characterization of Acute Ischemic Stroke Facilitates Metabolomic Biomarker Discovery. Applied Biochemistry and Biotechnology. 2022;194(11):5443–5455. doi:10.1007/S12010-022-04024-1,

14. Zhang X, Cai Y, Sit BHM, et al. Cell-Free Nucleic Acids for Early Diagnosis of Acute Ischemic Stroke: A Systematic Review and Meta-Analysis. International Journal of Molecular Sciences. 2025;26(4). doi:10.3390/IJMS26041530,

15. Page MJ, McKenzie JE, Bossuyt PM, et al. The PRISMA 2020 statement: an updated guideline for reporting systematic reviews. The BMJ. 2021;372:n71. doi:10.1136/BMJ.N71

16. Shi J, Luo D, Weng H, et al. Optimally estimating the sample standard deviation from the five-number summary. Research Synthesis Methods. 2020;11(5):641–654. doi:10.1002/JRSM.1429,

17. Gosling CJ, Solanes A, Fusar-Poli P, Radua J. metaumbrella: the first comprehensive suite to perform data analysis in umbrella reviews with stratification of the evidence. BMJ Mental Health. 2023;26(1). doi:10.1136/BMJMENT-2022-300534,

18. Fusar-Poli P, Radua J. Ten simple rules for conducting umbrella reviews. Evidence Based Mental Health. 2018;21(3):95–100. doi:10.1136/EBMENTAL-2018-300014

19. Misra S, Montaner J, Ramiro L, et al. Blood biomarkers for the diagnosis and differentiation of stroke: A systematic review and meta-analysis. International Journal of Stroke. 2020;15(7):704–721. doi:10.1177/1747493020946157,

20. Klokman VW, Koningstein FN, Dors JWW, et al. Blood biomarkers for the differentiation between central and peripheral vertigo in the emergency department: a systematic review and meta-analysis. Academic Emergency Medicine. 2024;31(4):371–385. doi:10.1111/ACEM.14864,

21. Wang Y, Li W, Yang J, et al. Association Between Cystatin C and the Risk of Ischemic Stroke: a Systematic Review and Meta-analysis. Journal of Molecular Neuroscience. 2019;69(3):444–449. doi:10.1007/S12031-019-01373-1,

22. Xing DG, Zhang DY, Wang ZF, Ding DL, Wang J, Wang YJ. Correlations of ANP genetic polymorphisms and serum levels with ischemic stroke risk: A meta-analysis. Genetic Testing and Molecular Biomarkers. 2014;18(5):349–356. doi:10.1089/GTMB.2013.0498,

23. Ye H, Wang L, Yang XK, Fan LP, Wang YG, Guo L. Serum S100B levels may be associated with cerebral infarction: A meta-analysis. Journal of the Neurological Sciences. 2015;348(1-2):81–88. doi:10.1016/j.jns.2014.11.010

24. Mangoni AA, Zinellu A. A Systematic Review and Meta-Analysis of Serum Concentrations of Ischaemia-Modified Albumin in Acute Ischaemic Stroke, Intracerebral Haemorrhage, and Subarachnoid Haemorrhage. Biomolecules. 2022;12(5). doi:10.3390/BIOM12050653,

25. Ahmad A, Islam Z, Manzoor Ahmad S, et al. The correlation of D-dimer to stroke diagnosis within 24 hours: A meta-analysis. Journal of Clinical Laboratory Analysis. 2022;36(3). doi:10.1002/JCLA.24271,

26. Blek N, Szwed P, Putowska P, et al. The diagnostic and prognostic value of copeptin in patients with acute ischemic stroke and transient ischemic attack: A systematic review and meta-analysis. Cardiology Journal. 2022;29(4):610–618. doi:10.5603/CJ.A2022.0045,

27. Hasan N, McColgan P, Bentley P, Edwards RJ, Sharma P. Towards the identification of blood biomarkers for acute stroke in humans: A comprehensive systematic review. British Journal of Clinical Pharmacology. 2012;74(2):230–240. doi:10.1111/J.1365-2125.2012.04212.X,

28. Huang X, Zhang M, Wang J, Hu F. Association between interleukin-6 levels and stroke: a systematic review and meta-analysis. Journal of International Medical Research. 2024;52(9). doi:10.1177/03000605241274626,

29. Mojtabavi H, Shaka Z, Momtazmanesh S, Ajdari A, Rezaei N. Circulating brain-derived neurotrophic factor as a potential biomarker in stroke: a systematic review and meta-analysis. Journal of Translational Medicine. 2022;20(1). doi:10.1186/S12967-022-03312-Y,

30. Sanchez JD, Martirosian RA, Mun KT, et al. Temporal Patterning of Neurofilament Light as a Blood-Based Biomarker for Stroke: A Systematic Review and Meta-Analysis. Frontiers in Neurology. 2022;13. doi:10.3389/FNEUR.2022.841898,

31. Seidkhani-Nahal A, Khosravi A, Mirzaei A, Basati G, Abbasi M, Noori-Zadeh A. Serum vascular endothelial growth factor (VEGF) levels in ischemic stroke patients: a systematic review and meta-analysis of case–control studies. Neurological Sciences. 2021;42(5):1811–1820. doi:10.1007/S10072-020-04698-7,

32. Słomka A, Kowalewski M, Zekanowska E, Suwalski P, Lorusso R, Eikelboom JW. Plasma Levels of Protein Z in Ischemic Stroke: A Systematic Review and Meta-Analysis. Thrombosis and Haemostasis. 2020;120(5):815–822. doi:10.1055/S-0040-1708878,

33. Ke C, Pan CW, Zhang Y, Zhu X, Zhang Y. Metabolomics facilitates the discovery of metabolic biomarkers and pathways for ischemic stroke: a systematic review. Metabolomics. 2019;15(12). doi:10.1007/S11306-019-1615-1,

34. Barrera-Vázquez OS, Gomez-Verjan JC, Ramírez-Aldana R, Torre PG Dela, Rivero-Segura NA. Structural and Pharmacological Network Analysis of miRNAs Involved in Acute Ischemic Stroke: A Systematic Review. International Journal of Molecular Sciences. 2022;23(9). doi:10.3390/IJMS23094663,

35. Bejleri J, Jirström E, Donovan P, Williams DJ, Pfeiffer S. Diagnostic and prognostic circulating microrna in acute stroke: A systematic and bioinformatic analysis of current evidence. Journal of Stroke. 2021;23(2):162–182. doi:10.5853/JOS.2020.05085,

36. Dewdney B, Trollope A, Moxon J, Thomas Manapurathe D, Biros E, Golledge J. Circulating MicroRNAs as Biomarkers for Acute Ischemic Stroke: A Systematic Review. Journal of Stroke and Cerebrovascular Diseases. 2018;27(3):522–530. doi:10.1016/j.jstrokecerebrovasdis.2017.09.058

37. Loggini A, Hornik J, Hornik A. The role of microRNAs as super-early biomarkers in acute ischemic stroke: A systematic review. Clinical Neurology and Neurosurgery. 2024;244. doi:10.1016/j.clineuro.2024.108416

38. Naik A, Adeleye O, Koester SW, et al. Cerebrospinal Fluid Biomarkers for Diagnosis and the Prognostication of Acute Ischemic Stroke: A Systematic Review. International Journal of Molecular Sciences. 2023;24(13). doi:10.3390/IJMS241310902,

39. Wang Y, Su X, Leung GHD, et al. Circulating microRNAs as diagnostic biomarkers for ischemic stroke: evidence from comprehensive analysis and real-world validation. International Journal of Medical Sciences. 2023;20(8):1009–1023. doi:10.7150/IJMS.83963,

40. Zhao J, Wang Q, Zhu R, Yang J. Circulating Non-coding RNAs as Potential Biomarkers for Ischemic Stroke: A Systematic Review. Journal of Molecular Neuroscience. 2022;72(8):1572–1585. doi:10.1007/S12031-022-01991-2,

41. Pan J, Fan W, Gu C, Xi Y, Wang Y, Wang P. Long Non-Coding RNAs as Diagnostic Biomarkers for Ischemic Stroke: A Systematic Review and Meta-Analysis. Genes. 2024;15(12). doi:10.3390/GENES15121620,

42. Tong Y, Ding ZH, Zhan FX, et al. The NLRP3 inflammasome and stroke. International Journal of Clinical and Experimental Medicine. 2015;8(4):4787. https://pmc.ncbi.nlm.nih.gov/articles/PMC4483817/. Accessed July 7, 2025.

43. Amalia L. Glial Fibrillary Acidic Protein (GFAP): Neuroinflammation Biomarker in Acute Ischemic Stroke. Journal of Inflammation Research. 2021;14:7501–7506. doi:10.2147/JIR.S342097,

44. Kalra L-P, Zylyftari S, Blums K, et al. Rapid Diagnosis of Intracerebral Hemorrhage in Patients With Acute Stroke by Measuring Prehospital GFAP Levels on a Point-of-Care Device (DETECT). Neurology. 2025;105(2). doi:10.1212/WNL.0000000000213823

45. Tannahill GM, Curtis AM, Adamik J, et al. Succinate is an inflammatory signal that induces IL-1β through HIF-1α. Nature. 2013;496(7444):238–242. doi:10.1038/NATURE11986,

46. Roth S, Wernsdorf SR, Liesz A. The role of circulating cell-free DNA as an inflammatory mediator after stroke. Seminars in Immunopathology. 2023;45(3):411–425. doi:10.1007/S00281-023-00993-5,

