## Supplementary Material for "Early-Phase Fluid Diagnostic Biomarkers in Acute Ischemic Stroke: An Umbrella Review"

### Supplementary figures

**Figure S1. Summary of the systematic review: direction of the meta-analyses for biochemistry biomarkers**

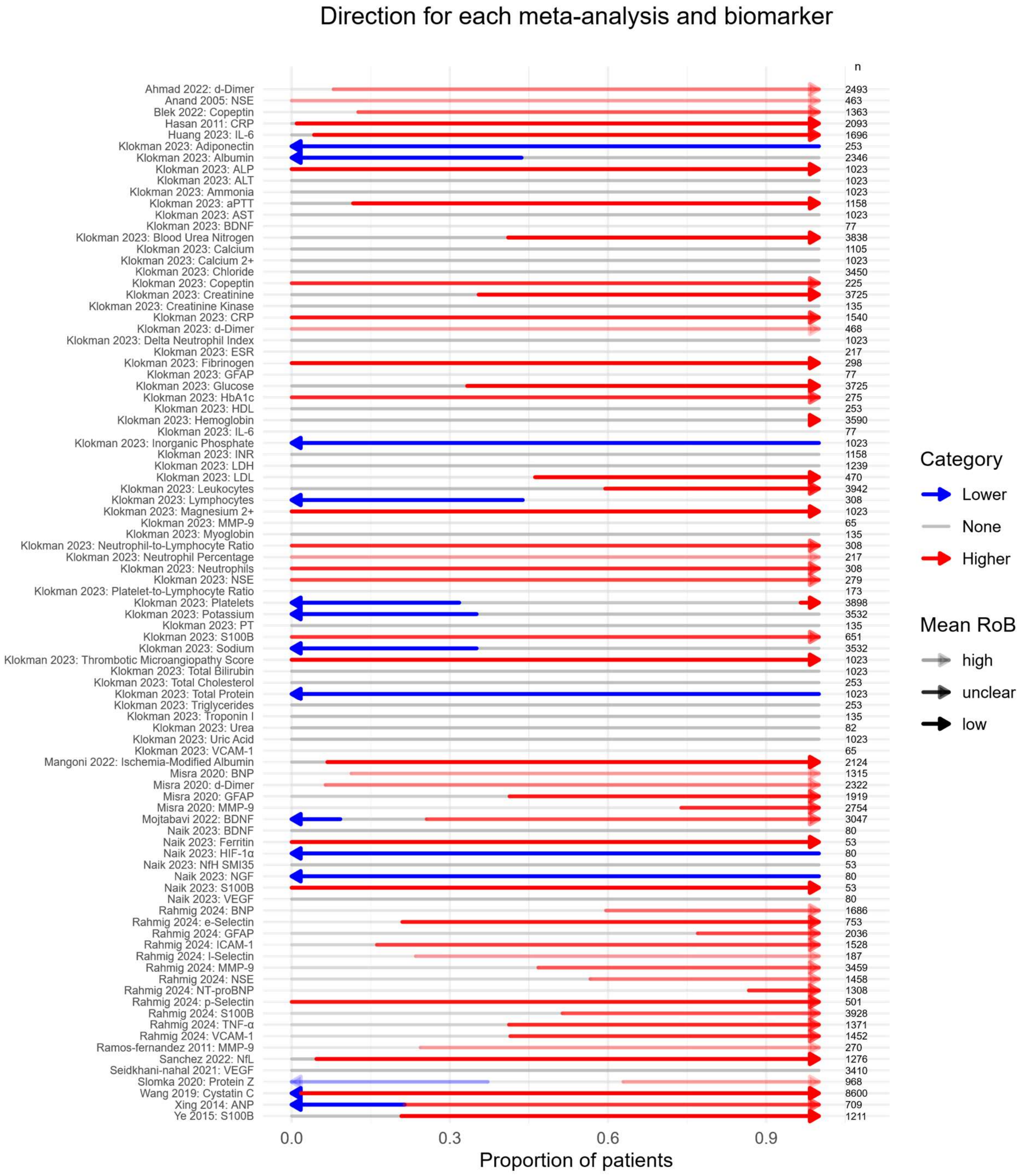

**Figure S2. Summary of the systematic review: direction of the meta-analyses for biochemistry biomarkers in the usual time-window**

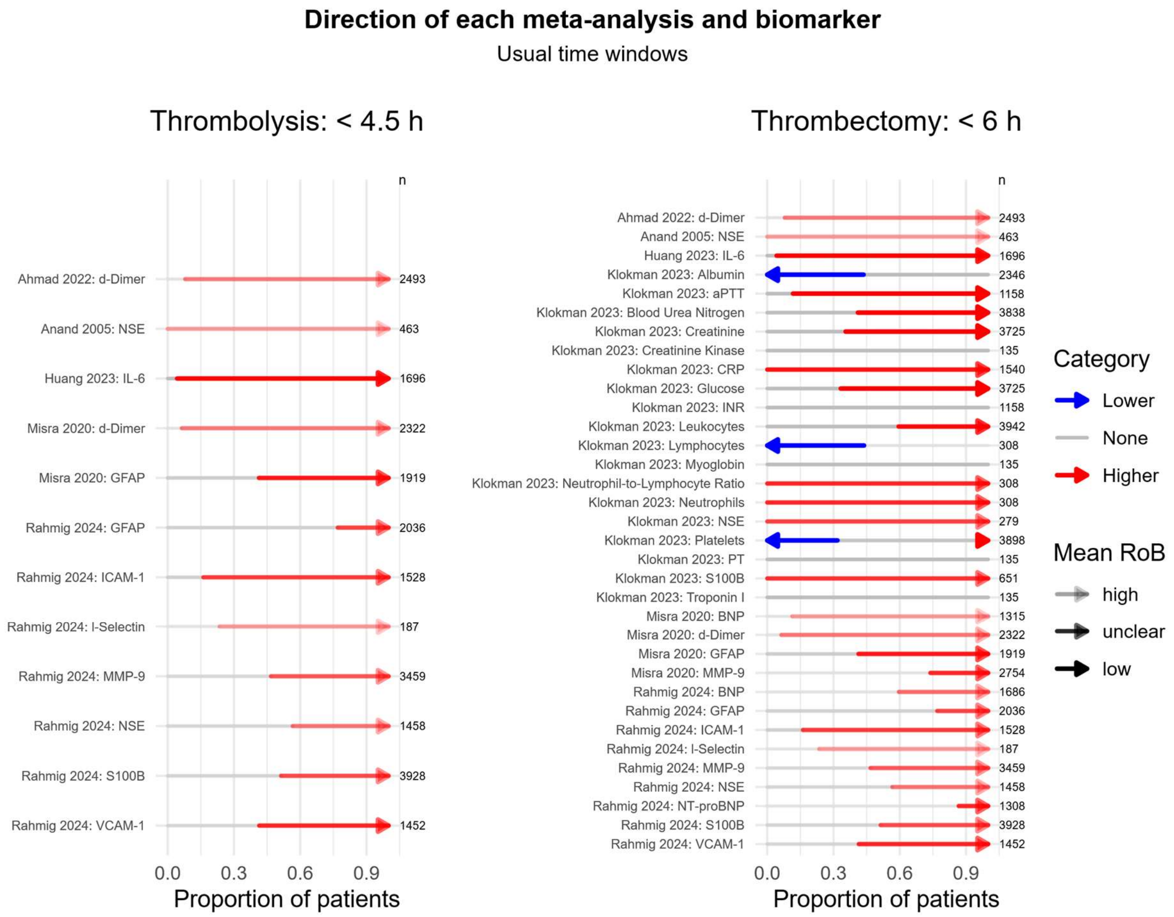

**Figure S3. Summary of the systematic review: direction of the meta-analyses for biochemistry biomarkers in the extended time-window**

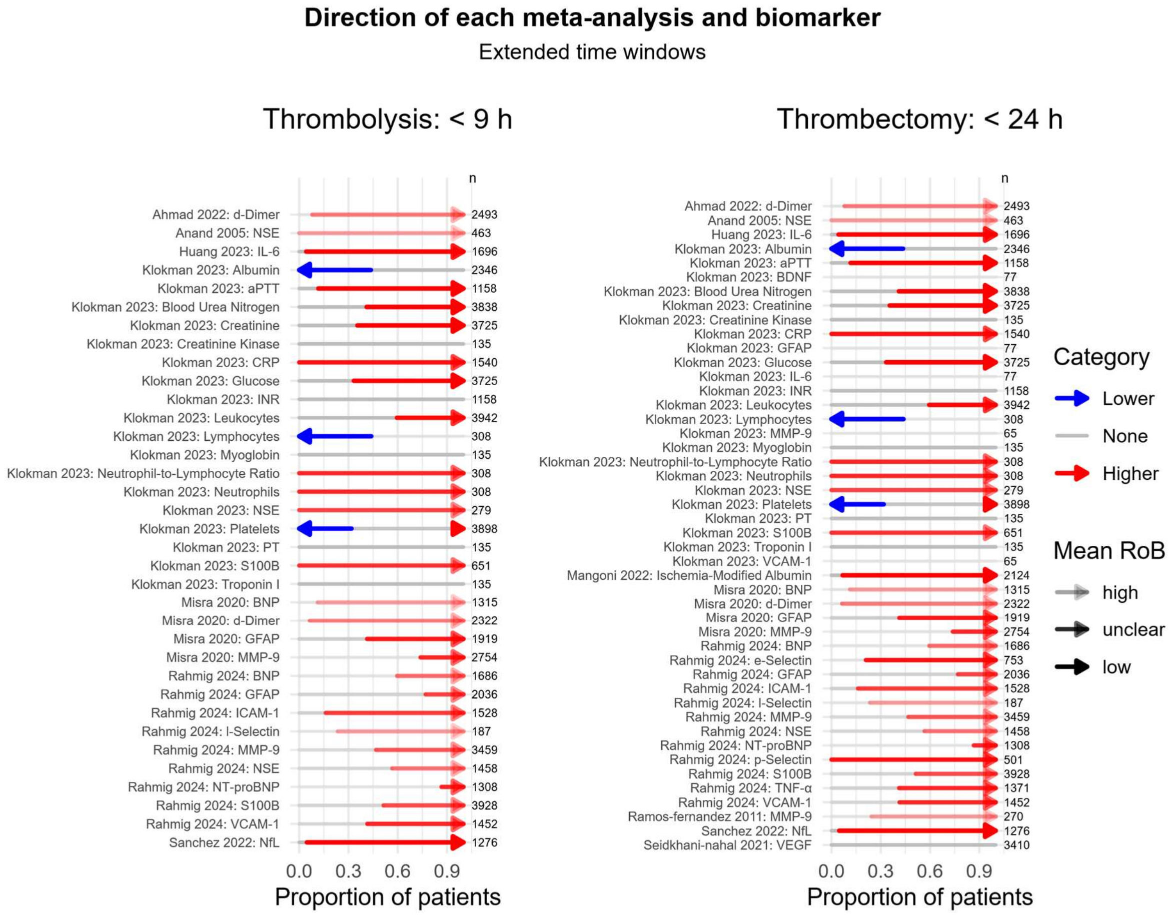

**Figures S4. Meta-analyses for each biomarkers, studies within 4.5 h (usual thrombolysis)**

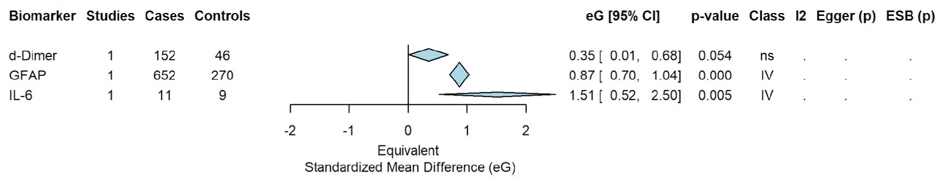

**Figure S5. Meta-analyses for each factor, studies within 6 hours (usual thrombectomy)**

| Biomarker | Studies | Cases | Controls | eG [95% CI] | p-value | Class | I2 | Egger (p) | ESB (p) |
|---|---|---|---|---|---|---|---|---|---|
| Lymphocytes | 1 | 35 | 100 | -0.59 [ -0.97, -0.20] | 0.008 | IV | . | . | . |
| aPTT | 1 | 35 | 100 | -0.34 [ -0.72, 0.04] | 0.098 | ns | . | . | . |
| Albumin | 1 | 35 | 100 | -0.33 [ -0.72, 0.06] | 0.111 | ns | . | . | . |
| Blood Urea Nitrogen | 1 | 35 | 100 | -0.04 [ -0.42, 0.34] | 0.839 | ns | . | . | . |
| INR | 1 | 35 | 100 | 0.00 [ -0.38, 0.38] | 1.000 | ns | . | . | . |
| Glucose | 1 | 35 | 100 | 0.25 [ -0.13, 0.63] | 0.215 | ns | . | . | . |
| BNP | 2 | 189 | 106 | 0.41 [ -0.06, 0.88] | 0.087 | ns | 74.3% | . | 0.605 |
| Platelets | 1 | 35 | 100 | 0.41 [ 0.02, 0.80] | 0.052 | ns | . | . | . |
| MMP-9 | 4 | 601 | 231 | 0.43 [ -0.24, 1.11] | 0.211 | ns | 88.1% | 0.101 | 0.774 |
| d-Dimer | 3 | 630 | 181 | 0.45 [ 0.26, 0.64] | 0.000 | IV | 25.9% | 0.634 | 0.108 |
| GFAP | 3 | 1282 | 501 | 0.45 [ -0.02, 0.92] | 0.061 | ns | 92.9% | 0.847 | 0.610 |
| c-Reactive Protein | 1 | 35 | 100 | 0.65 [ 0.26, 1.03] | 0.004 | IV | . | . | . |
| Leukocytes | 1 | 35 | 100 | 0.75 [ 0.35, 1.14] | 0.002 | IV | . | . | . |
| Creatinine | 1 | 35 | 100 | 0.86 [ 0.47, 1.26] | 0.000 | IV | . | . | . |
| Neutrophils | 1 | 35 | 100 | 0.99 [ 0.59, 1.40] | 0.000 | IV | . | . | . |
| Neutrophil-to-Lymphocyte Ratio | 1 | 35 | 100 | 1.43 [ 1.01, 1.85] | 0.000 | IV | . | . | . |
| IL-6 | 1 | 11 | 9 | 1.51 [ 0.52, 2.50] | 0.005 | IV | . | . | . |
| S100B | 1 | 21 | 64 | 2.12 [ 1.54, 2.71] | 0.000 | IV | . | . | . |
| NSE | 1 | 21 | 64 | 2.97 [ 2.30, 3.64] | 0.000 | IV | . | . | . |

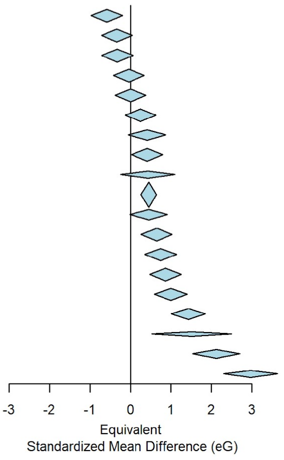

### Figure S6. Meta-analyses for each factor, studies within 9 h (extended thrombolysis)

| Biomarker | Studies | Cases | Controls | eG [95% CI] | p-value | Class | I2 | Egger (p) | ESB (p) |
|---|---|---|---|---|---|---|---|---|---|
| Lymphocytes | 1 | 35 | 100 | -0.59 [ -0.97, -0.20] | 0.008 | IV | . | . | . |
| aPTT | 1 | 35 | 100 | -0.34 [ -0.72, 0.04] | 0.098 | ns | . | . | . |
| Albumin | 1 | 35 | 100 | -0.33 [ -0.72, 0.06] | 0.111 | ns | . | . | . |
| Blood Urea Nitrogen | 1 | 35 | 100 | -0.04 [ -0.42, 0.34] | 0.839 | ns | . | . | . |
| INR | 1 | 35 | 100 | 0.00 [ -0.38, 0.38] | 1.000 | ns | . | . | . |
| Glucose | 1 | 35 | 100 | 0.25 [ -0.13, 0.63] | 0.215 | ns | . | . | . |
| BNP | 2 | 189 | 106 | 0.41 [ -0.06, 0.88] | 0.087 | ns | 74.3% | . | 0.605 |
| Platelets | 1 | 35 | 100 | 0.41 [ 0.02, 0.80] | 0.052 | ns | . | . | . |
| MMP-9 | 4 | 601 | 231 | 0.43 [ -0.24, 1.11] | 0.211 | ns | 88.1% | 0.101 | 0.774 |
| d-Dimer | 3 | 630 | 181 | 0.45 [ 0.26, 0.64] | 0.000 | IV | 25.9% | 0.634 | 0.108 |
| GFAP | 3 | 1282 | 501 | 0.45 [ -0.02, 0.92] | 0.061 | ns | 92.9% | 0.847 | 0.610 |
| c-Reactive Protein | 1 | 35 | 100 | 0.65 [ 0.26, 1.03] | 0.004 | IV | . | . | . |
| Leukocytes | 1 | 35 | 100 | 0.75 [ 0.35, 1.14] | 0.002 | IV | . | . | . |
| Creatinine | 1 | 35 | 100 | 0.86 [ 0.47, 1.26] | 0.000 | IV | . | . | . |
| Neutrophils | 1 | 35 | 100 | 0.99 [ 0.59, 1.40] | 0.000 | IV | . | . | . |
| Neutrophil-to-Lymphocyte Ratio | 1 | 35 | 100 | 1.43 [ 1.01, 1.85] | 0.000 | IV | . | . | . |
| IL-6 | 1 | 11 | 9 | 1.51 [ 0.52, 2.50] | 0.005 | IV | . | . | . |
| S100B | 2 | 64 | 138 | 2.82 [ 1.45, 4.19] | 0.000 | IV | 91.0% | . | 0.465 |
| NSE | 2 | 64 | 138 | 2.89 [ 2.48, 3.30] | 0.000 | IV | 0.0% | . | . |

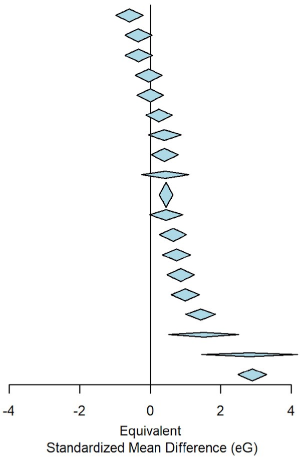

**Figure S7. Meta-analyses for each factor, studies within 24 h (extended thrombectomy)**

| Biomarker | Studies | Cases | Controls | eG [95% CI] | p-value | Class | I2 | Egger (p) | ESB (p) |
|---|---|---|---|---|---|---|---|---|---|
| VEGF | 6 | 457 | 352 | -4.44 [-10.65, 1.77] | 0.161 | ns | 99.3% | 0.189 | 0.098 |
| Lymphocytes | 1 | 35 | 100 | -0.59 [ -0.97, -0.20] | 0.004 | IV | . | . | . |
| aPTT | 1 | 35 | 100 | -0.34 [ -0.72, 0.04] | 0.087 | ns | . | . | . |
| Albumin | 1 | 35 | 100 | -0.33 [ -0.72, 0.06] | 0.101 | ns | . | . | . |
| Blood Urea Nitrogen | 1 | 35 | 100 | -0.04 [ -0.42, 0.34] | 0.838 | ns | . | . | . |
| INR | 1 | 35 | 100 | 0.00 [ -0.38, 0.38] | 1.000 | ns | . | . | . |
| Glucose | 1 | 35 | 100 | 0.25 [ -0.13, 0.63] | 0.206 | ns | . | . | . |
| BNP | 5 | 1070 | 245 | 0.41 [ 0.23, 0.58] | 0.000 | III | 18.1% | 0.452 | 0.033 |
| Platelets | 1 | 35 | 100 | 0.41 [ 0.02, 0.80] | 0.043 | IV | . | . | . |
| MMP-9 | 11 | 2075 | 679 | 0.46 [ 0.21, 0.71] | 0.000 | III | 77.6% | 0.084 | 0.707 |
| GFAP | 4 | 1361 | 558 | 0.48 [ 0.14, 0.82] | 0.005 | IV | 89.3% | 0.799 | 0.527 |
| c-Reactive Protein | 1 | 35 | 100 | 0.65 [ 0.26, 1.03] | 0.002 | IV | . | . | . |
| Leukocytes | 1 | 35 | 100 | 0.75 [ 0.35, 1.14] | 0.001 | IV | . | . | . |
| Creatinine | 1 | 35 | 100 | 0.86 [ 0.47, 1.26] | 0.000 | IV | . | . | . |
| Neutrophils | 1 | 35 | 100 | 0.99 [ 0.59, 1.40] | 0.000 | IV | . | . | . |
| IL-6 | 9 | 491 | 322 | 1.28 [ 0.67, 1.89] | 0.000 | IV | 87.7% | 0.045 | 0.035 |
| Neutrophil-to-Lymphocyte Ratio | 1 | 35 | 100 | 1.43 [ 1.01, 1.85] | 0.000 | IV | . | . | . |
| d-Dimer | 11 | 1827 | 542 | 1.89 [ 0.17, 3.62] | 0.031 | IV | 97.1% | 0.007 | 0.001 |
| S100B | 3 | 107 | 160 | 2.11 [ 0.52, 3.70] | 0.009 | IV | 96.0% | 0.245 | 0.307 |
| Ischemia-Modified Albumin | 18 | 1136 | 988 | 2.54 [ 1.66, 3.42] | 0.000 | II | 96.5% | 0.002 | 0.405 |
| NSE | 2 | 64 | 138 | 2.89 [ 2.48, 3.30] | 0.000 | IV | 0.0% | . | . |

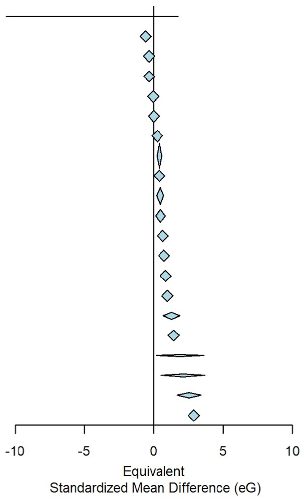

### Supplementary tables

### Table S1. Characteristics of the included meta-analyses for biochemistry biomarkers

| | | Higher in Stroke | | | | | | No difference | | | | | | Lower in Stroke | | | | | |
|---|---|---|---|---|---|---|---|---|---|---|---|---|---|---|---|---|---|---|---|
| Review | Biomarker | n | R | M | Subjects | Time to onset | Ctrl | n | R | M | Subjects | Time to onset | Ctrl | n | R | M | Subjects | Time to onset | Ctrl |
| Ahmad 2022 | d-Dimer | 9 | L: 2; U: 6; H: 1 | B: 9 | IS: 1824; C: 471 | <6 h: 3; <9 h: 3; <24 h: 9; | SM: 9 | 1 | U: 1 | B: 1 | IS: 152; C: 46 | <4.5 h: 1; <6 h: 1; <9 h: 1; <24 h: 1; | SM: 1; | 0 | | | | | |
| Anand 2005 | NSE | 5 | U: 5 | B: 5 | IS: 259; C: 204 | <4.5 h: 1; <6 h: 1; <9 h: 1; <24 h: 4; over: 1 | HC: 5 | 0 | | | | | | 0 | | | | | |
| Blek 2022 | Copeptin | 4 | L: 1; U: 3 | | IS: 315; C: 876 | over: 4 | HC: 2; OC: 1 | 1 | U: 1 | | IS: 109; C: 63 | over: 1 | HC: 1; | 0 | | | | | |
| Hasan 2011 | CRP | 6 | | | IS: 703; C: 1370 | over: 6 | HC: 6 | 1 | | | IS: 11; C: 9 | over: 1 | HC: 1; | 0 | | | | | |
| Huang 2023 | IL-6 | 14 | L: 12; U: 2 | | IS: 939; C: 685 | <4.5 h: 1; <6 h: 1; <9 h: 1; <24 h: 8; over: 6 | HC: 14 | 1 | L: 1 | | IS: 36; C: 36 | <24 h: 1; | HC: 1; | 0 | | | | | |
| Klokman 2023 | ALP | 1 | L: 1 | B: 1 | IS: 119; C: 904 | over: 1 | OC: 1 | 0 | | | | | | 0 | | | | | |
| Klokman 2023 | ALT | 0 | | | | | | 1 | L: 1 | B: 1 | IS: 119; C: 904 | over: 1 | OC: 1; | 0 | | | | | |
| Klokman 2023 | AST | 0 | | | | | | 1 | L: 1 | B: 1 | IS: 119; C: 904 | over: 1 | OC: 1; | 0 | | | | | |
| Klokman 2023 | Adiponectin | 0 | | | | | | 0 | | | | | | 1 | L: 1 | B: 1 | IS: 68; C: 185 | over: 1 | OC: 1; |
| Klokman 2023 | Albumin | 0 | | | | | | 2 | L: 2 | B: 2 | IS: 88; C: 1235 | <6 h: 1; <9 h: 1; <24 h: 1; over: 1 | OC: 2; | 1 | L: 1 | B: 1 | IS: 119; C: 904 | over: 1 | OC: 1; |
| Klokman 2023 | Ammonia | 0 | | | | | | 1 | L: 1 | B: 1 | IS: 119; C: 904 | over: 1 | OC: 1; | 0 | | | | | |
| Klokman 2023 | BDNF | 0 | | | | | | 1 | U: 1 | B: 1 | IS: 28; C: 49 | <24 h: 1; | OC: 1; | 0 | | | | | |
| Klokman 2023 | Blood Urea Nitrogen | 2 | L: 2 | B: 2 | IS: 174; C: 2088 | over: 2 | OC: 2 | 3 | L: 3 | B: 3 | IS: 156; C: 1420 | <6 h: 1; <9 h: 1; <24 h: 1; over: 2 | OC: 3; | 0 | | | | | |
| Klokman 2023 | CRP | 3 | L: 2; U: 1 | B: 3 | IS: 144; C: 1396 | <6 h: 1; <9 h: 1; <24 h: 1; over: 2 | OC: 3 | 0 | | | | | | 0 | | | | | |
| Klokman 2023 | Calcium | 0 | | | | | | 2 | L: 2 | B: 2 | IS: 150; C: 955 | over: 2 | OC: 2; | 0 | | | | | |
| Klokman 2023 | Calcium 2+ | 0 | | | | | | 1 | L: 1 | B: 1 | IS: 119; C: 904 | over: 1 | OC: 1; | 0 | | | | | |
| Klokman 2023 | Chloride | 0 | | | | | | 3 | L: 3 | B: 3 | IS: 227; C: 3223 | over: 3 | OC: 3; | 0 | | | | | |
| Klokman 2023 | Copeptin | 2 | L: 1; U: 1 | B: 2 | IS: 58; C: 167 | over: 2 | OC: 2 | 0 | | | | | | 0 | | | | | |
| Klokman 2023 | Creatinine | 4 | L: 4 | B: 4 | IS: 230; C: 2174 | <6 h: 1; <9 h: 1; <24 h: 1; over: 3 | OC: 4 | 2 | L: 2 | B: 2 | IS: 86; C: 1235 | over: 2 | OC: 2; | 0 | | | | | |
| Klokman 2023 | Creatinine Kinase | 0 | | | | | | 1 | L: 1 | B: 1 | IS: 35; C: 100 | <6 h: 1; <9 h: 1; <24 h: 1; | OC: 1; | 0 | | | | | |
| Klokman 2023 | Delta Neutrophil Index | 0 | | | | | | 1 | L: 1 | B: 1 | IS: 119; C: 904 | over: 1 | OC: 1; | 0 | | | | | |
| Klokman 2023 | ESR | 0 | | | | | | 1 | U: 1 | B: 1 | IS: 56; C: 161 | over: 1 | OC: 1; | 0 | | | | | |
| Klokman 2023 | Fibrinogen | 1 | L: 1 | B: 1 | IS: 126; C: 172 | over: 1 | OC: 1 | 0 | | | | | | 0 | | | | | |
| Klokman 2023 | GFAP | 0 | | | | | | 1 | U: 1 | B: 1 | IS: 28; C: 49 | <24 h: 1; | OC: 1; | 0 | | | | | |
| Klokman 2023 | Glucose | 3 | L: 3 | B: 3 | IS: 131; C: 2354 | over: 3 | OC: 3 | 3 | L: 3 | B: 3 | IS: 185; C: 1055 | <6 h: 1; <9 h: 1; <24 h: 1; over: 2 | OC: 3; | 0 | | | | | |
| Klokman 2023 | HDL | 0 | | | | | | 1 | L: 1 | B: 1 | IS: 68; C: 185 | over: 1 | OC: 1; | 0 | | | | | |
| Klokman 2023 | HbA1c | 2 | L: 1; U: 1 | B: 2 | IS: 79; C: 196 | over: 2 | OC: 2 | 0 | | | | | | 0 | | | | | |

| | | | | | | | | | | | | | | | | | | | |
|---|---|---|---|---|---|---|---|---|---|---|---|---|---|---|---|---|---|---|---|
| Klokman 2023 | Hemoglobin | 1 | L: 1 | B: 1 | IS: 23; C: 35 | over: 1 | OC: 1 | 4 | L: 4 | B: 4 | IS: 258; C: 3274 | over: 4 | OC: 4; | 0 | | | | | |
| Klokman 2023 | IL-6 | 0 | | | | | | 1 | U: 1 | B: 1 | IS: 28; C: 49 | <24 h: 1; | OC: 1; | 0 | | | | | |
| Klokman 2023 | INR | 0 | | | | | | 2 | L: 2 | B: 2 | IS: 154; C: 1004 | <6 h: 1; <9 h: 1; <24 h: 1; over: 1 | OC: 2; | 0 | | | | | |
| Klokman 2023 | Inorganic Phosphate | 0 | | | | | | 0 | | | | | | 1 | L: 1 | B: 1 | IS: 119; C: 904 | over: 1 | OC: 1; |
| Klokman 2023 | LDH | 0 | | | | | | 1 | L: 1 | B: 1 | IS: 55; C: 1184 | over: 1 | OC: 1; | 0 | | | | | |
| Klokman 2023 | LDL | 1 | L: 1 | B: 1 | IS: 68; C: 185 | over: 1 | OC: 1 | 1 | U: 1 | B: 1 | IS: 56; C: 161 | over: 1 | OC: 1; | 0 | | | | | |
| Klokman 2023 | Leukocytes | 4 | L: 3; U: 1 | B: 4 | IS: 167; C: 1431 | <6 h: 1; <9 h: 1; <24 h: 1; over: 3 | OC: 4 | 3 | L: 3 | B: 3 | IS: 205; C: 2139 | over: 3 | OC: 3; | 0 | | | | | |
| Klokman 2023 | Lymphocytes | 0 | | | | | | 1 | U: 1 | B: 1 | IS: 62; C: 111 | over: 1 | OC: 1; | 1 | L: 1 | B: 1 | IS: 35; C: 100 | <6 h: 1; <9 h: 1; <24 h: 1; | OC: 1; |
| Klokman 2023 | MMP-9 | 0 | | | | | | 1 | U: 1 | B: 1 | IS: 43; C: 22 | <24 h: 1; | OC: 1; | 0 | | | | | |
| Klokman 2023 | Magnesium 2+ | 1 | L: 1 | B: 1 | IS: 119; C: 904 | over: 1 | OC: 1 | 0 | | | | | | 0 | | | | | |
| Klokman 2023 | Myoglobin | 0 | | | | | | 1 | L: 1 | B: 1 | IS: 35; C: 100 | <6 h: 1; <9 h: 1; <24 h: 1; | OC: 1; | 0 | | | | | |
| Klokman 2023 | NSE | 3 | L: 1; U: 2 | B: 3 | IS: 92; C: 187 | <6 h: 1; <9 h: 2; <24 h: 3; | OC: 3 | 0 | | | | | | 0 | | | | | |
| Klokman 2023 | Neutrophil Percentage | 1 | U: 1 | B: 1 | IS: 56; C: 161 | over: 1 | OC: 1 | 0 | | | | | | 0 | | | | | |
| Klokman 2023 | Neutrophil-to-Lymphocyte Ratio | 2 | L: 1; U: 1 | B: 2 | IS: 97; C: 211 | <6 h: 1; <9 h: 1; <24 h: 1; over: 1 | OC: 2 | 0 | | | | | | 0 | | | | | |
| Klokman 2023 | Neutrophils | 2 | L: 1; U: 1 | B: 2 | IS: 97; C: 211 | <6 h: 1; <9 h: 1; <24 h: 1; over: 1 | OC: 2 | 0 | | | | | | 0 | | | | | |
| Klokman 2023 | PT | 0 | | | | | | 1 | L: 1 | B: 1 | IS: 35; C: 100 | <6 h: 1; <9 h: 1; <24 h: 1; | OC: 1; | 0 | | | | | |
| Klokman 2023 | Platelet-to-Lymphocyte Ratio | 0 | | | | | | 1 | U: 1 | B: 1 | IS: 62; C: 111 | over: 1 | OC: 1; | 0 | | | | | |
| Klokman 2023 | Platelets | 1 | L: 1 | B: 1 | IS: 35; C: 100 | <6 h: 1; <9 h: 1; <24 h: 1; | OC: 1 | 5 | L: 4; U: 1 | B: 5 | IS: 288; C: 2236 | over: 5 | OC: 5; | 1 | L: 1 | B: 1 | IS: 55; C: 1184 | over: 1 | OC: 1; |
| Klokman 2023 | Potassium | 0 | | | | | | 3 | L: 3 | B: 3 | IS: 203; C: 2090 | over: 3 | OC: 3; | 1 | L: 1 | B: 1 | IS: 55; C: 1184 | over: 1 | OC: 1; |
| Klokman 2023 | S100B | 7 | L: 3; U: 4 | B: 7 | IS: 224; C: 427 | <6 h: 1; <9 h: 2; <24 h: 4; over: 3 | OC: 7 | 0 | | | | | | 0 | | | | | |
| Klokman 2023 | Sodium | 0 | | | | | | 3 | L: 3 | B: 3 | IS: 203; C: 2090 | over: 3 | OC: 3; | 1 | L: 1 | B: 1 | IS: 55; C: 1184 | over: 1 | OC: 1; |
| Klokman 2023 | Thrombotic Microangiopathy Score | 1 | L: 1 | B: 1 | IS: 119; C: 904 | over: 1 | OC: 1 | 0 | | | | | | 0 | | | | | |
| Klokman 2023 | Total Bilirubin | 0 | | | | | | 1 | L: 1 | B: 1 | IS: 119; C: 904 | over: 1 | OC: 1; | 0 | | | | | |
| Klokman 2023 | Total Cholesterol | 0 | | | | | | 1 | L: 1 | B: 1 | IS: 68; C: 185 | over: 1 | OC: 1; | 0 | | | | | |
| Klokman 2023 | Total Protein | 0 | | | | | | 0 | | | | | | 1 | L: 1 | B: 1 | IS: 119; C: 904 | over: 1 | OC: 1; |
| Klokman 2023 | Triglycerides | 0 | | | | | | 1 | L: 1 | B: 1 | IS: 68; C: 185 | over: 1 | OC: 1; | 0 | | | | | |
| Klokman 2023 | Troponin I | 0 | | | | | | 1 | L: 1 | B: 1 | IS: 35; C: 100 | <6 h: 1; <9 h: 1; <24 h: 1; | OC: 1; | 0 | | | | | |
| Klokman 2023 | Urea | 0 | | | | | | 1 | L: 1 | B: 1 | IS: 31; C: 51 | over: 1 | OC: 1; | 0 | | | | | |
| Klokman 2023 | Uric Acid | 0 | | | | | | 1 | L: 1 | B: 1 | IS: 119; C: 904 | over: 1 | OC: 1; | 0 | | | | | |
| Klokman 2023 | VCAM-1 | 0 | | | | | | 1 | U: 1 | B: 1 | IS: 43; C: 22 | <24 h: 1; | OC: 1; | 0 | | | | | |
| Klokman 2023 | aPTT | 1 | L: 1 | B: 1 | IS: 119; C: 904 | over: 1 | OC: 1 | 1 | L: 1 | B: 1 | IS: 35; C: 100 | <6 h: 1; <9 h: 1; <24 h: 1; | OC: 1; | 0 | | | | | |
| Klokman 2023 | d-Dimer | 1 | U: 1 | B: 1 | IS: 38; C: 430 | over: 1 | OC: 1 | 0 | | | | | | 0 | | | | | |

| | | | | | | | | | | | | | | | | | | | |
|---|---|---|---|---|---|---|---|---|---|---|---|---|---|---|---|---|---|---|---|
| Mangoni 2022 | Ischemia-Modified Albumin | 16 | L: 16 | B: 16 | IS: 1045; C: 935 | <24 h: 16; | HC: 16 | 2 | L: 2 | B: 2 | IS: 91; C: 53 | <24 h: 2; | HC: 2; | 0 | | | | | |
| Misra 2020 | BNP | 4 | L: 1; U: 2; H: 1 | B: 4 | IS: 970; C: 196 | <6 h: 1; <9 h: 1; <24 h: 4; | SM: 2; HC: 2 | 1 | U: 1 | B: 1 | IS: 100; C: 49 | <6 h: 1; <9 h: 1; <24 h: 1; | SM: 1; | 0 | | | | | |
| Misra 2020 | GFAP | 5 | L: 3; U: 2 | B: 5 | IS: 647; C: 479 | <4.5 h: 2; <6 h: 4; <9 h: 4; <24 h: 5; | HC: 5; | 5 | L: 3; U: 2 | B: 5 | IS: 714; C: 79 | <4.5 h: 2; <6 h: 5; <9 h: 5; <24 h: 5; | SM: 5; | 0 | | | | | |
| Misra 2020 | MMP-9 | 4 | L: 3; H: 1 | B: 4 | IS: 495; C: 224 | <6 h: 1; <9 h: 1; <24 h: 4; | SM: 2; HC: 2; | 7 | L: 1; U: 5; H: 1 | B: 7 | IS: 1580; C: 455 | <6 h: 3; <9 h: 3; <24 h: 7; | SM: 3; HC: 4; | 0 | | | | | |
| Misra 2020 | d-Dimer | 9 | L: 2; U: 6; H: 1 | B: 9 | IS: 1705; C: 468 | <4.5 h: 1; <6 h: 3; <9 h: 3; <24 h: 9; | SM: 4; HC: 5; | 1 | U: 1 | B: 1 | IS: 100; C: 49 | <24 h: 1; | SM: 1; | 0 | | | | | |
| Mojtabavi 2022 | BDNF | 10 | L: 5; U: 3; H: 2 | B: 10 | IS: 933; C: 1334 | over: 10 | HC: 10; | 5 | L: 4; U: 1 | B: 5 | IS: 152; C: 347 | over: 5 | HC: 5; | 2 | L: 2 | B: 2 | IS: 156; C: 125 | over: 2 | HC: 2; |
| Naik 2023 | BDNF | 0 | | | | | | 1 | L: 1 | CSF: 1 | IS: 50; C: 30 | over: 1 | HC: 1; | 0 | | | | | |
| Naik 2023 | Ferritin | 1 | L: 1 | CSF: 1 | IS: 33; C: 20 | over: 1 | HC: 1; | 0 | | | | | | 0 | | | | | |
| Naik 2023 | HIF-1α | 0 | | | | | | 0 | | | | | | 1 | L: 1 | CSF: 1 | IS: 50; C: 30 | over: 1 | HC: 1; |
| Naik 2023 | NGF | 0 | | | | | | 0 | | | | | | 1 | L: 1 | CSF: 1 | IS: 50; C: 30 | over: 1 | HC: 1; |
| Naik 2023 | NfH SMI35 | 0 | | | | | | 1 | L: 1 | CSF: 1 | IS: 33; C: 20 | over: 1 | HC: 1; | 0 | | | | | |
| Naik 2023 | S100B | 1 | L: 1 | CSF: 1 | IS: 33; C: 20 | over: 1 | HC: 1; | 0 | | | | | | 0 | | | | | |
| Naik 2023 | VEGF | 0 | | | | | | 1 | L: 1 | CSF: 1 | IS: 50; C: 30 | over: 1 | HC: 1; | 0 | | | | | |
| Rahmig 2024 | BNP | 6 | L: 1; U: 5 | B: 6 | IS: 398; C: 283 | <6 h: 1; <9 h: 1; <24 h: 6; | SM: 2; HC: 3; OC: 1; | 1 | U: 1 | B: 1 | IS: 915; C: 90 | <24 h: 1; | SM: 1; | 0 | | | | | |
| Rahmig 2024 | GFAP | 3 | L: 1; U: 2 | B: 3 | IS: 301; C: 167 | <24 h: 3; | SM: 1; HC: 2; | 11 | L: 5; U: 6 | B: 11 | IS: 1118; C: 450 | <4.5 h: 6; <6 h: 9; <9 h: 10; <24 h: 11; | SM: 7; HC: 4; | 0 | | | | | |
| Rahmig 2024 | ICAM-1 | 8 | L: 3; U: 5 | B: 8 | IS: 726; C: 555 | <24 h: 8; | HC: 5; OC: 2; NC: 1 | 2 | L: 1; U: 1 | B: 2 | IS: 132; C: 115 | <4.5 h: 1; <6 h: 1; <9 h: 1; <24 h: 2; | HC: 1; NC: 1 | 0 | | | | | |
| Rahmig 2024 | MMP-9 | 8 | L: 2; U: 6 | B: 8 | IS: 1386; C: 455 | <6 h: 1; <9 h: 1; <24 h: 8; | SM: 3; HC: 5; | 10 | L: 3; U: 7 | B: 10 | IS: 868; C: 750 | <4.5 h: 1; <6 h: 3; <9 h: 4; <24 h: 10; | SM: 2; HC: 8; | 0 | | | | | |
| Rahmig 2024 | NSE | 7 | L: 1; U: 6 | B: 7 | IS: 330; C: 302 | <4.5 h: 1; <6 h: 1; <9 h: 1; <24 h: 7; | SM: 1; HC: 6; | 3 | L: 1; U: 2 | B: 3 | IS: 601; C: 225 | <6 h: 1; <9 h: 1; <24 h: 3; | SM: 2; OC: 1; | 0 | | | | | |
| Rahmig 2024 | NT-proBNP | 2 | L: 1; U: 1 | B: 2 | IS: 87; C: 87 | <24 h: 2; | HC: 2; | 1 | U: 1 | B: 1 | IS: 941; C: 193 | <6 h: 1; <9 h: 1; <24 h: 1; | SM: 1; | 0 | | | | | |
| Rahmig 2024 | S100B | 10 | L: 3; U: 7 | B: 10 | IS: 989; C: 922 | <4.5 h: 1; <6 h: 2; <9 h: 2; <24 h: 10; | SM: 5; HC: 4; NC: 1 | 5 | L: 2; U: 3 | B: 5 | IS: 1547; C: 470 | <6 h: 1; <9 h: 1; <24 h: 5; | SM: 3; HC: 2; | 0 | | | | | |
| Rahmig 2024 | TNF-α | 6 | L: 3; U: 3 | B: 6 | IS: 410; C: 396 | <24 h: 6; | HC: 2; NS: 1; OC: 1; NC: 2 | 4 | L: 2; U: 2 | B: 4 | IS: 332; C: 233 | <24 h: 4; | SM: 2; HC: 2; | 0 | | | | | |
| Rahmig 2024 | VCAM-1 | 6 | L: 3; U: 3 | B: 6 | IS: 416; C: 434 | <4.5 h: 1; <6 h: 1; <9 h: 1; <24 h: 6; | HC: 4; OC: 1; NC: 1 | 3 | L: 1; U: 2 | B: 3 | IS: 391; C: 211 | <24 h: 3; | HC: 1; OC: 1; NC: 1 | 0 | | | | | |
| Rahmig 2024 | e-Selectin | 3 | L: 2; U: 1 | B: 3 | IS: 294; C: 301 | <24 h: 3; | HC: 2; OC: 1; | 2 | U: 2 | B: 2 | IS: 91; C: 67 | <24 h: 2; | HC: 1; OC: 1; | 0 | | | | | |
| Rahmig 2024 | l-Selectin | 1 | U: 1 | B: 1 | IS: 67; C: 76 | <24 h: 1; | HC: 1; | 1 | U: 1 | B: 1 | IS: 22; C: 22 | <4.5 h: 1; <6 h: 1; <9 h: 1; <24 h: 1; | HC: 1; | 0 | | | | | |
| Rahmig 2024 | p-Selectin | 3 | L: 2; U: 1 | B: 3 | IS: 252; C: 249 | <24 h: 3; | HC: 1; OC: 1; NC: 1 | 0 | | | | | | 0 | | | | | |
| Ramos-Fernandez 2011 | MMP-9 | 2 | U: 2 | | IS: 107; C: 97 | <24 h: 2; | HC: 2; | 1 | U: 1 | | IS: 29; C: 37 | <24 h: 1; | HC: 1; | 0 | | | | | |
| Sanchez 2022 | NfL | 2 | L: 2 | B: 2 | IS: 568; C: 648 | over: 2 | HC: 2; | 1 | L: 1 | B: 1 | IS: 31; C: 29 | <9 h: 1; <24 h: 1; | HC: 1; | 0 | | | | | |
| Seidkhani-Nahal 2021 | VEGF | 0 | | | | | | 42 | | B: 42 | IS: 1855; C: 1555 | <24 h: 16; over: 26 | HC: 42; | 0 | | | | | |
| Slomka 2020 | Protein Z | 1 | U: 1 | B: 1 | IS: 173; C: 186 | over: 1 | HC: 1; | 1 | U: 1 | B: 1 | IS: 124; C: 125 | over: 1 | HC: 1; | 1 | U: 1 | B: 1 | IS: 154; C: 206 | over: 1 | HC: 1; |
| Wang 2019 | Cystatin C | 8 | L: 8 | B: 8 | IS: 3690; C: 4756 | over: 8 | HC: 8; | 0 | | | | | | 1 | L: 1 | B: 1 | IS: 83; C: 71 | over: 1 | HC: 1; |
| Xing 2014 | ANP | 7 | L: 3; U: 4 | B: 7 | IS: 328; C: 229 | over: 7 | HC: 7; | 0 | | | | | | 1 | L: 1 | B: 1 | IS: 52; C: 100 | over: 1 | HC: 1; |
| Ye 2015 | S100B | 8 | L: 8 | B: 8 | IS: 637; C: 322 | over: 8 | HC: 8; | 2 | L: 2 | B: 2 | IS: 136; C: 116 | over: 2 | HC: 2; | 0 | | | | | |

### Table S2. Characteristics of the included meta-analyses for miRNA biomarkers

| | | Higher in Stroke | | | | | | Mixed or no difference | | | | | | Lower in Stroke | | | | | |
|---|---|---|---|---|---|---|---|---|---|---|---|---|---|---|---|---|---|---|---|
| Review | Biomarker | n | R | M | Subjects | Time to onset | Ctrl | n | R | M | Subjects | Time to onset | Ctrl | n | R | M | Subjects | Time to onset | Ctrl |
| barrera-vazquez 2022 | Let-7b | 1 | | B: 1 | IS: 87; C: 13 | <24 h: 1 | NS: 1 | 0 | | | | | | 0 | | | | | |
| barrera-vazquez 2022 | Let-7c | 1 | | CSF: 1 | IS: 10; C: 10 | <24 h: 1 | NS: 1 | 0 | | | | | | 0 | | | | | |
| barrera-vazquez 2022 | Let-7d-3p | 0 | | | | | | 0 | | | | | | 1 | | B: 1 | IS: 80; C: 2360 | <24 h: 1 | NS: 1 |
| barrera-vazquez 2022 | Let-7e | 1 | | B: 1 | IS: 87; C: 13 | <24 h: 1 | NS: 1 | 0 | | | | | | 0 | | | | | |
| barrera-vazquez 2022 | Let-7i-5p | 1 | | B: 1 | IS: 40; C: 39 | <24 h: 1 | NS: 1 | 0 | | | | | | 1 | | B: 1 | IS: 46; C: 39 | <24 h: 1 | NS: 1 |
| barrera-vazquez 2022 | miR-101 | 0 | | | | | | 0 | | | | | | 1 | | B: 1 | IS: 106; C: 110 | <24 h: 1 | NS: 1 |
| barrera-vazquez 2022 | miR-106b-5p | 1 | | B: 1 | IS: 33; C: 23 | <24 h: 1 | NS: 1 | 0 | | | | | | 0 | | | | | |
| barrera-vazquez 2022 | miR-107 | 3 | | B: 2; CSF: 1 | IS: 168; C: 102 | <24 h: 3 | NS: 3 | 0 | | | | | | 0 | | | | | |
| barrera-vazquez 2022 | miR-124 | 0 | | | | | | 0 | | | | | | 1 | | B: 1 | IS: 54; C: 51 | <24 h: 1 | NS: 1 |
| barrera-vazquez 2022 | miR-124-3p | 3 | | B: 2; CSF: 1 | IS: 62; C: 21 | <24 h: 3 | NS: 3 | 0 | | | | | | 2 | | B: 2 | IS: 55; C: 0 | <24 h: 2 | NS: 2 |
| barrera-vazquez 2022 | miR-1246 | 1 | | CSF: 1 | IS: 21; C: 21 | <24 h: 1 | NS: 1 | 0 | | | | | | 0 | | | | | |
| barrera-vazquez 2022 | miR-125a | 1 | | B: 1 | IS: 87; C: 13 | <24 h: 1 | NS: 1 | 0 | | | | | | 0 | | | | | |
| barrera-vazquez 2022 | miR-125a-5p | 1 | | B: 1 | IS: 20; C: 20 | <24 h: 1 | NS: 1 | 0 | | | | | | 0 | | | | | |
| barrera-vazquez 2022 | miR-125b | 1 | | B: 1 | IS: 87; C: 13 | <24 h: 1 | NS: 1 | 0 | | | | | | 0 | | | | | |
| barrera-vazquez 2022 | miR-125b-5p | 2 | | B: 2 | IS: 49; C: 20 | <24 h: 2 | NS: 2 | 0 | | | | | | 1 | | B: 1 | IS: 55; C: 0 | <24 h: 1 | NS: 1 |
| barrera-vazquez 2022 | miR-126 | 0 | | | | | | 0 | | | | | | 2 | | B: 2 | IS: 254; C: 258 | <24 h: 2 | NS: 2 |
| barrera-vazquez 2022 | miR-1264 | 1 | | B: 1 | | <24 h: 1 | NS: 1 | 0 | | | | | | 0 | | | | | |
| barrera-vazquez 2022 | miR-1271-5p-B1 | 0 | | | | | | 0 | | | | | | 1 | | B: 1 | IS: 80; C: 2360 | <24 h: 1 | NS: 1 |
| barrera-vazquez 2022 | miR-1273g-3p | 0 | | | | | | 0 | | | | | | 1 | | B: 1 | IS: 17; C: 25 | <24 h: 1 | NS: 1 |
| barrera-vazquez 2022 | miR-1279 | 0 | | | | | | 0 | | | | | | 1 | | B: 1 | | <24 h: 1 | NS: 1 |
| barrera-vazquez 2022 | miR-128-3p | 2 | | B: 1; CSF: 1 | IS: 21; C: 21 | <24 h: 2 | NS: 2 | 0 | | | | | | 0 | | | | | |

| | | | | | | | | | | | | |
|---|---|---|---|---|---|---|---|---|---|---|---|---|
| barrera-vazquez 2022 | miR-128b | 1 | B: 1 | IS: 114; C: 58 | <24 h: 1 | NS: 1 | 0 | 0 | | | | |
| barrera-vazquez 2022 | miR-129-1-3p | 0 | | | | | 0 | 1 | B: 1 | IS: 80; C: 2360 | <24 h: 1 | NS: 1 |
| barrera-vazquez 2022 | miR-129-2-3p | 0 | | | | | 0 | 1 | B: 1 | IS: 270; C: 270 | <24 h: 1 | NS: 1 |
| barrera-vazquez 2022 | miR-130a | 0 | | | | | 0 | 2 | B: 2 | IS: 254; C: 258 | <24 h: 2 | NS: 2 |
| barrera-vazquez 2022 | miR-130a-3p | 1 | B: 1 | IS: 33; C: 23 | <24 h: 1 | NS: 1 | 0 | 0 | | | | |
| barrera-vazquez 2022 | miR-130b-3p | 1 | B: 1 | IS: 33; C: 23 | <24 h: 1 | NS: 1 | 0 | 0 | | | | |
| barrera-vazquez 2022 | miR-134 | 1 | E: 1 | IS: 50; C: 50 | <24 h: 1 | NS: 1 | 0 | 0 | | | | |
| barrera-vazquez 2022 | miR-140-3p | 1 | B: 1 | IS: 33; C: 23 | <24 h: 1 | NS: 1 | 0 | 0 | | | | |
| barrera-vazquez 2022 | miR-140-5p | 1 | B: 1 | IS: 10; C: 10 | <24 h: 1 | NS: 1 | 0 | 0 | | | | |
| barrera-vazquez 2022 | miR-143-3p | 1 | B: 1 | IS: 20; C: 20 | <24 h: 1 | NS: 1 | 0 | 0 | | | | |
| barrera-vazquez 2022 | miR-145 | 1 | B: 1 | IS: 30; C: 30 | <24 h: 1 | NS: 1 | 0 | 0 | | | | |
| barrera-vazquez 2022 | miR-146a-5p | 0 | | | | | 0 | 1 | E: 1 | | <24 h: 1 | NS: 1 |
| barrera-vazquez 2022 | miR-146b | 1 | B: 1 | IS: 30; C: 30 | <24 h: 1 | NS: 1 | 0 | 0 | | | | |
| barrera-vazquez 2022 | miR-148b-3p | 0 | | | | | 0 | 1 | B: 1 | IS: 77; C: 42 | <24 h: 1 | NS: 1 |
| barrera-vazquez 2022 | miR-151a-3p | 2 | B: 2 | IS: 43; C: 33 | <24 h: 2 | NS: 2 | 0 | 0 | | | | |
| barrera-vazquez 2022 | miR-151b | 1 | B: 1 | IS: 77; C: 42 | <24 h: 1 | NS: 1 | 0 | 0 | | | | |
| barrera-vazquez 2022 | miR-153 | 1 | B: 1 | IS: 114; C: 58 | <24 h: 1 | NS: 1 | 0 | 0 | | | | |
| barrera-vazquez 2022 | miR-155 | 1 | B: 1 | | <24 h: 1 | NS: 1 | 0 | 0 | | | | |
| barrera-vazquez 2022 | miR-15a | 1 | B: 1 | IS: 106; C: 120 | <24 h: 1 | NS: 1 | 0 | 1 | B: 1 | IS: 46; C: 39 | <24 h: 1 | NS: 1 |
| barrera-vazquez 2022 | miR-16 | 2 | B: 2 | IS: 178; C: 120 | <24 h: 2 | NS: 2 | 0 | 0 | | | | |
| barrera-vazquez 2022 | miR-16-1-3p | 1 | B: 1 | IS: 80; C: 2360 | <24 h: 1 | NS: 1 | 0 | 0 | | | | |
| barrera-vazquez 2022 | miR-16-5p | 1 | B: 1 | IS: 33; C: 23 | <24 h: 1 | NS: 1 | 0 | 0 | | | | |
| barrera-vazquez 2022 | miR-17-5p | 1 | B: 1 | IS: 106; C: 120 | <24 h: 1 | NS: 1 | 0 | 0 | | | | |
| barrera-vazquez 2022 | miR-17-92 | 0 | | | | | 0 | 1 | E: 1 | | <24 h: 1 | NS: 1 |

| | | | | | | | | | | | | |
|---|---|---|---|---|---|---|---|---|---|---|---|---|
| barrera-vazquez 2022 | miR-181a-3p | 1 | B: 1 | IS: 10; C: 10 | <24 h: 1 | NS: 1 | 0 | 0 | | | | |
| barrera-vazquez 2022 | miR-181a-5p | 1 | B: 1 | IS: 227; C: 92 | <24 h: 1 | NS: 1 | 0 | 0 | | | | |
| barrera-vazquez 2022 | miR-181b-3p | 1 | B: 1 | IS: 5; C: 5 | <24 h: 1 | NS: 1 | 0 | 0 | | | | |
| barrera-vazquez 2022 | miR-181d-5p | 1 | B: 1 | IS: 5; C: 5 | <24 h: 1 | NS: 1 | 0 | 0 | | | | |
| barrera-vazquez 2022 | miR-185 | 2 | B: 2 | IS: 254; C: 258 | <24 h: 2 | NS: 2 | 0 | 0 | | | | |
| barrera-vazquez 2022 | miR-18b-5p | 0 | | | | | 0 | 1 | B: 1 | IS: 10; C: 10 | <24 h: 1 | NS: 1 |
| barrera-vazquez 2022 | miR-1908 | 0 | | | | | 0 | 1 | B: 1 | IS: 87; C: 13 | <24 h: 1 | NS: 1 |
| barrera-vazquez 2022 | miR-192-5p | 1 | B: 1 | IS: 29; C: 0 | <24 h: 1 | NS: 1 | 0 | 1 | B: 1 | IS: 55; C: 0 | <24 h: 1 | NS: 1 |
| barrera-vazquez 2022 | miR-193b-3p | 0 | | | | | 0 | 1 | B: 1 | IS: 80; C: 2360 | <24 h: 1 | NS: 1 |
| barrera-vazquez 2022 | miR-19a | 0 | | | | | 0 | 1 | B: 1 | IS: 106; C: 110 | <24 h: 1 | NS: 1 |
| barrera-vazquez 2022 | miR-21 | 1 | B: 1 | IS: 30; C: 30 | <24 h: 1 | NS: 1 | 0 | 0 | | | | |
| barrera-vazquez 2022 | miR-21-5p | 1 | B: 1 | IS: 227; C: 92 | <24 h: 1 | NS: 1 | 0 | 0 | | | | |
| barrera-vazquez 2022 | miR-218 | 2 | B: 2 | IS: 254; C: 258 | <24 h: 2 | NS: 2 | 0 | 0 | | | | |
| barrera-vazquez 2022 | miR-221 | 0 | | | | | 0 | 1 | B: 1 | IS: 30; C: 30 | <24 h: 1 | NS: 1 |
| barrera-vazquez 2022 | miR-221-3p | 1 | CSF: 1 | IS: 10; C: 10 | <24 h: 1 | NS: 1 | 0 | 1 | B: 1 | IS: 78; C: 39 | <24 h: 1 | NS: 1 |
| barrera-vazquez 2022 | miR-222 | 2 | B: 2 | IS: 254; C: 258 | <24 h: 2 | NS: 2 | 0 | 0 | | | | |
| barrera-vazquez 2022 | miR-223 | 1 | E: 1 | IS: 50; C: 33 | <24 h: 1 | NS: 1 | 0 | 0 | | | | |
| barrera-vazquez 2022 | miR-223-3p | 1 | B: 1 | IS: 33; C: 23 | <24 h: 1 | NS: 1 | 0 | 0 | | | | |
| barrera-vazquez 2022 | miR-23a | 0 | | | | | 0 | 1 | B: 1 | IS: 30; C: 30 | <24 h: 1 | NS: 1 |
| barrera-vazquez 2022 | miR-23b-3p | 1 | B: 1 | IS: 227; C: 92 | <24 h: 1 | NS: 1 | 0 | 0 | | | | |
| barrera-vazquez 2022 | miR-25-3p | 1 | B: 1 | IS: 33; C: 23 | <24 h: 1 | NS: 1 | 0 | 0 | | | | |
| barrera-vazquez 2022 | miR-26b-5p | 1 | B: 1 | IS: 80; C: 2360 | <24 h: 1 | NS: 1 | 0 | 0 | | | | |
| barrera-vazquez 2022 | miR-27b-3p | 1 | B: 1 | IS: 77; C: 42 | <24 h: 1 | NS: 1 | 0 | 0 | | | | |
| barrera-vazquez 2022 | miR-28-5p | 1 | B: 1 | IS: 55; C: 2360 | <24 h: 1 | NS: 1 | 0 | 0 | | | | |

| | | | | | | | | | | | | |
|---|---|---|---|---|---|---|---|---|---|---|---|---|
| barrera-vazquez 2022 | miR-296 | 0 | | | | | 0 | 1 | B: 1 | IS: 106; C: 110 | <24 h: 1 | NS: 1 |
| barrera-vazquez 2022 | miR-29b | 1 | B: 1 | IS: 30; C: 30 | <24 h: 1 | NS: 1 | 0 | 0 | | | | |
| barrera-vazquez 2022 | miR-29b-2-5p | 1 | B: 1 | IS: 80; C: 2360 | <24 h: 1 | NS: 1 | 0 | 0 | | | | |
| barrera-vazquez 2022 | miR-29b-3p | 1 | B: 1 | IS: 227; C: 92 | <24 h: 1 | NS: 1 | 0 | 0 | | | | |
| barrera-vazquez 2022 | miR-30a-5p | 1 | E: 1 | IS: 33; C: 24 | <24 h: 1 | NS: 1 | 0 | 0 | | | | |
| barrera-vazquez 2022 | miR-30d-5p | 1 | B: 1 | IS: 17; C: 25 | <24 h: 1 | NS: 1 | 0 | 0 | | | | |
| barrera-vazquez 2022 | miR-31-5p | 1 | B: 1 | IS: 10; C: 10 | <24 h: 1 | NS: 1 | 0 | 0 | | | | |
| barrera-vazquez 2022 | miR-320b | 3 | B: 3 | IS: 152; C: 4745 | <24 h: 3 | NS: 3 | 0 | 0 | | | | |
| barrera-vazquez 2022 | miR-320d | 1 | B: 1 | IS: 17; C: 25 | <24 h: 1 | NS: 1 | 0 | 0 | | | | |
| barrera-vazquez 2022 | miR-320e | 1 | B: 1 | IS: 17; C: 25 | <24 h: 1 | NS: 1 | 0 | 0 | | | | |
| barrera-vazquez 2022 | miR-324-3p | 1 | B: 1 | IS: 55; C: 2360 | <24 h: 1 | NS: 1 | 0 | 0 | | | | |
| barrera-vazquez 2022 | miR-328-3p | 0 | | | | | 0 | 1 | B: 1 | IS: 39; C: 20 | <24 h: 1 | NS: 1 |
| barrera-vazquez 2022 | miR-335-3p | 0 | | | | | 0 | 1 | B: 1 | IS: 17; C: 25 | <24 h: 1 | NS: 1 |
| barrera-vazquez 2022 | miR-378 | 0 | | | | | 0 | 1 | B: 1 | IS: 106; C: 110 | <24 h: 1 | NS: 1 |
| barrera-vazquez 2022 | miR-382-5p | 0 | | | | | 0 | 1 | B: 1 | IS: 78; C: 39 | <24 h: 1 | NS: 1 |
| barrera-vazquez 2022 | miR-424 | 1 | Ce: 1 | IS: 40; C: 27 | <24 h: 1 | NS: 1 | 0 | 0 | | | | |
| barrera-vazquez 2022 | miR-4454 | 0 | | | | | 0 | 2 | B: 2 | IS: 50; C: 48 | <24 h: 2 | NS: 2 |
| barrera-vazquez 2022 | miR-4634 | 1 | B: 1 | IS: 17; C: 25 | <24 h: 1 | NS: 1 | 0 | 0 | | | | |
| barrera-vazquez 2022 | miR-483-3p | 0 | | | | | 0 | 1 | B: 1 | IS: 55; C: 2360 | <24 h: 1 | NS: 1 |
| barrera-vazquez 2022 | miR-484 | 1 | B: 1 | IS: 33; C: 23 | <24 h: 1 | NS: 1 | 0 | 1 | B: 1 | IS: 80; C: 2360 | <24 h: 1 | NS: 1 |
| barrera-vazquez 2022 | miR-5100 | 0 | | | | | 0 | 1 | B: 1 | IS: 17; C: 25 | <24 h: 1 | NS: 1 |
| barrera-vazquez 2022 | miR-523-3p | 1 | CSF: 1 | IS: 10; C: 10 | <24 h: 1 | NS: 1 | 0 | 0 | | | | |
| barrera-vazquez 2022 | miR-570 | 0 | | | | | 0 | 1 | B: 1 | | <24 h: 1 | NS: 1 |
| barrera-vazquez 2022 | miR-574-3p | 1 | B: 1 | IS: 55; C: 2360 | <24 h: 1 | NS: 1 | 0 | 0 | | | | |

| | | | | | | | | | | | | | | | | | | | |
|---|---|---|---|---|---|---|---|---|---|---|---|---|---|---|---|---|---|---|---|
| barrera-vazquez 2022 | miR-593 | 0 | | | | | | 0 | | | | | | 1 | | B: 1 | | <24 h: 1 | NS: 1 |
| barrera-vazquez 2022 | miR-605-3p | 1 | | B: 1 | IS: 5; C: 5 | <24 h: 1 | NS: 1 | 0 | | | | | | 0 | | | | | |
| barrera-vazquez 2022 | miR-605-5p | 1 | | B: 1 | IS: 5; C: 5 | <24 h: 1 | NS: 1 | 0 | | | | | | 0 | | | | | |
| barrera-vazquez 2022 | miR-625-3p | 0 | | | | | | 0 | | | | | | 1 | | B: 1 | IS: 55; C: 2360 | <24 h: 1 | NS: 1 |
| barrera-vazquez 2022 | miR-629-3p | 0 | | | | | | 0 | | | | | | 1 | | B: 1 | IS: 80; C: 2360 | <24 h: 1 | NS: 1 |
| barrera-vazquez 2022 | miR-630 | 0 | | | | | | 0 | | | | | | 1 | | B: 1 | | <24 h: 1 | NS: 1 |
| barrera-vazquez 2022 | miR-638 | 0 | | | | | | 0 | | | | | | 1 | | B: 1 | IS: 22; C: 36 | <24 h: 1 | NS: 1 |
| barrera-vazquez 2022 | miR-653 | 0 | | | | | | 0 | | | | | | 1 | | B: 1 | | <24 h: 1 | NS: 1 |
| barrera-vazquez 2022 | miR-7-2-3p | 1 | | B: 1 | IS: 87; C: 13 | <24 h: 1 | NS: 1 | 0 | | | | | | 0 | | | | | |
| barrera-vazquez 2022 | miR-9-3p | 1 | | CSF: 1 | IS: 21; C: 21 | <24 h: 1 | NS: 1 | 0 | | | | | | 0 | | | | | |
| barrera-vazquez 2022 | miR-9-5p | 1 | | B: 1 | | <24 h: 1 | NS: 1 | 0 | | | | | | 0 | | | | | |
| barrera-vazquez 2022 | miR-93-5p | 1 | | B: 1 | IS: 33; C: 23 | <24 h: 1 | NS: 1 | 0 | | | | | | 0 | | | | | |
| bejleri 2021 | Let-7b | 0 | | | | | | 1 | H: 1 | B: 1 | IS: 38; C: 50 | <24 h: 1 | HC: 1 | 0 | | | | | |
| bejleri 2021 | Let-7d-3p | 0 | | | | | | 0 | | | | | | 1 | U: 1 | B: 1 | IS: 169; C: 24 | <24 h: 1 | HC: 1 |
| bejleri 2021 | Let-7e | 1 | U: 1 | B: 1 | IS: 11; C: 10 | <24 h: 1 | HC: 1 | 0 | | | | | | 0 | | | | | |
| bejleri 2021 | Let-7e-5p | 1 | H: 1 | B: 1 | IS: 346; C: 346 | <24 h: 1 | HC: 1 | 0 | | | | | | 0 | | | | | |
| bejleri 2021 | miR-101 | 0 | | | | | | 0 | | | | | | 2 | H: 2 | B: 2 | IS: 212; C: 220 | <24 h: 2 | HC: 2 |
| bejleri 2021 | miR-106b-5p | 2 | U: 2 | B: 2 | IS: 253; C: 198 | <24 h: 2 | HC: 2 | 0 | | | | | | 0 | | | | | |
| bejleri 2021 | miR-124 | 0 | | | | | | 0 | | | | | | 1 | L: 1 | B: 1 | IS: 31; C: 11 | <24 h: 1 | HC: 1 |
| bejleri 2021 | miR-1246 | 1 | U: 1 | B: 1 | IS: 117; C: 82 | <24 h: 1 | HC: 1 | 0 | | | | | | 0 | | | | | |
| bejleri 2021 | miR-125a-5p | 1 | U: 1 | B: 1 | IS: 260; C: 160 | <24 h: 1 | HC: 1 | 0 | | | | | | 0 | | | | | |
| bejleri 2021 | miR-125b-2-3p | 1 | U: 1 | B: 1 | IS: 169; C: 24 | <24 h: 1 | HC: 1 | 0 | | | | | | 0 | | | | | |
| bejleri 2021 | miR-125b-5p | 1 | U: 1 | B: 1 | IS: 260; C: 160 | <24 h: 1 | HC: 1 | 0 | | | | | | 0 | | | | | |
| bejleri 2021 | miR-126 | 0 | | | | | | 0 | | | | | | 3 | H: 3 | B: 3 | IS: 292; C: 308 | <24 h: 3 | HC: 3 |

| | | | | | | | | | | | | | | |
|---|---|---|---|---|---|---|---|---|---|---|---|---|---|---|
| bejleri 2021 | miR-1261 | 1 | U: 1 | B: 1 | IS: 169; C: 24 | <24 h: 1 | HC: 1 | 0 | 0 | | | | | |
| bejleri 2021 | miR-1299 | 0 | | | | | | 0 | 1 | U: 1 | B: 1 | IS: 169; C: 24 | <24 h: 1 | HC: 1 |
| bejleri 2021 | miR-130a | 0 | | | | | | 0 | 3 | U: 1; H: 2 | B: 3 | IS: 423; C: 282 | <24 h: 3 | HC: 3 |
| bejleri 2021 | miR-1321 | 1 | U: 1 | B: 1 | IS: 169; C: 24 | <24 h: 1 | HC: 1 | 0 | 0 | | | | | |
| bejleri 2021 | miR-134 | 1 | H: 1 | B: 1 | IS: 50; C: 50 | <24 h: 1 | HC: 1 | 0 | 0 | | | | | |
| bejleri 2021 | miR-143-3p | 1 | U: 1 | B: 1 | IS: 260; C: 160 | <24 h: 1 | HC: 1 | 0 | 0 | | | | | |
| bejleri 2021 | miR-145 | 1 | H: 1 | B: 1 | IS: 146; C: 96 | <24 h: 1 | HC: 1 | 0 | 0 | | | | | |
| bejleri 2021 | miR-146b | 1 | H: 1 | B: 1 | IS: 128; C: 102 | <24 h: 1 | HC: 1 | 0 | 0 | | | | | |
| bejleri 2021 | miR-16 | 2 | U: 2 | B: 2 | IS: 114; C: 53 | <6 h: 1; <9 h: 1; <24 h: 2 | HC: 2 | 0 | 0 | | | | | |
| bejleri 2021 | miR-185 | 2 | H: 2 | B: 2 | IS: 254; C: 258 | <24 h: 2 | HC: 2 | 0 | 0 | | | | | |
| bejleri 2021 | miR-186-5p | 0 | | | | | | 0 | 1 | U: 1 | B: 1 | IS: 11; C: 10 | <24 h: 1 | HC: 1 |
| bejleri 2021 | miR-206 | 1 | H: 1 | B: 1 | IS: 106; C: 110 | <24 h: 1 | HC: 1 | 0 | 0 | | | | | |
| bejleri 2021 | miR-208a | 0 | | | | | | 0 | 1 | U: 1 | B: 1 | IS: 169; C: 24 | <24 h: 1 | HC: 1 |
| bejleri 2021 | miR-218 | 2 | H: 2 | B: 2 | IS: 254; C: 258 | <24 h: 2 | HC: 2 | 0 | 0 | | | | | |
| bejleri 2021 | miR-219 | 0 | | | | | | 0 | 1 | L: 1 | B: 1 | IS: 31; C: 11 | <24 h: 1 | HC: 1 |
| bejleri 2021 | miR-22-5p | 0 | | | | | | 0 | 1 | U: 1 | B: 1 | IS: 169; C: 24 | <24 h: 1 | HC: 1 |
| bejleri 2021 | miR-221 | 0 | | | | | | 0 | 1 | H: 1 | B: 1 | IS: 146; C: 96 | <24 h: 1 | HC: 1 |
| bejleri 2021 | miR-221-3p | 0 | | | | | | 0 | 1 | U: 1 | B: 1 | IS: 78; C: 39 | <24 h: 1 | HC: 1 |
| bejleri 2021 | miR-222 | 2 | H: 2 | B: 2 | IS: 254; C: 258 | <24 h: 2 | HC: 2 | 0 | 0 | | | | | |
| bejleri 2021 | miR-23a | 0 | | | | | | 0 | 1 | H: 1 | B: 1 | IS: 146; C: 96 | <24 h: 1 | HC: 1 |
| bejleri 2021 | miR-27a-5p | 1 | U: 1 | B: 1 | IS: 169; C: 24 | <24 h: 1 | HC: 1 | 0 | 0 | | | | | |
| bejleri 2021 | miR-30a | 0 | | | | | | 0 | 1 | H: 1 | B: 1 | IS: 38; C: 50 | <24 h: 1 | HC: 1 |
| bejleri 2021 | miR-30c | 0 | | | | | | 0 | 1 | U: 1 | B: 1 | IS: 169; C: 24 | <24 h: 1 | HC: 1 |
| bejleri 2021 | miR-32-3p | 1 | U: 1 | B: 1 | IS: 117; C: 82 | <24 h: 1 | HC: 1 | 0 | 0 | | | | | |

| | | | | | | | | | | | | | | |
|---|---|---|---|---|---|---|---|---|---|---|---|---|---|---|
| bejleri 2021 | miR-32-5p | 0 | | | | | | 0 | 1 | U: 1 | B: 1 | IS: 11; C: 10 | <24 h: 1 | HC: 1 |
| bejleri 2021 | miR-320b | 0 | | | | | | 0 | 1 | U: 1 | B: 1 | IS: 169; C: 24 | <24 h: 1 | HC: 1 |
| bejleri 2021 | miR-320d | 0 | | | | | | 0 | 2 | U: 2 | B: 2 | IS: 305; C: 140 | <24 h: 2 | HC: 2 |
| bejleri 2021 | miR-320e | 0 | | | | | | 0 | 1 | U: 1 | B: 1 | IS: 136; C: 116 | <24 h: 1 | HC: 1 |
| bejleri 2021 | miR-340 | 0 | | | | | | 0 | 1 | U: 1 | B: 1 | IS: 169; C: 24 | <24 h: 1 | HC: 1 |
| bejleri 2021 | miR-340-5p | 0 | | | | | | 0 | 1 | U: 1 | B: 1 | IS: 11; C: 10 | <24 h: 1 | HC: 1 |
| bejleri 2021 | miR-378 | 0 | | | | | | 0 | 2 | H: 2 | B: 2 | IS: 212; C: 220 | <24 h: 2 | HC: 2 |
| bejleri 2021 | miR-382-5p | 0 | | | | | | 0 | 1 | U: 1 | B: 1 | IS: 78; C: 39 | <24 h: 1 | HC: 1 |
| bejleri 2021 | miR-422a | 1 | U: 1 | B: 1 | IS: 169; C: 24 | <24 h: 1 | HC: 1 | 0 | 0 | | | | | |
| bejleri 2021 | miR-423-3p | 0 | | | | | | 0 | 1 | U: 1 | B: 1 | IS: 169; C: 24 | <24 h: 1 | HC: 1 |
| bejleri 2021 | miR-424 | 1 | U: 1 | Ce: 1 | IS: 40; C: 27 | <6 h: 1; <9 h: 1; <24 h: 1 | HC: 1 | 0 | 0 | | | | | |
| bejleri 2021 | miR-4306 | 1 | U: 1 | B: 1 | IS: 136; C: 116 | <24 h: 1 | HC: 1 | 0 | 0 | | | | | |
| bejleri 2021 | miR-488 | 1 | U: 1 | B: 1 | IS: 169; C: 24 | <24 h: 1 | HC: 1 | 0 | 0 | | | | | |
| bejleri 2021 | miR-502-5p | 0 | | | | | | 0 | 1 | U: 1 | B: 1 | IS: 169; C: 24 | <24 h: 1 | HC: 1 |
| bejleri 2021 | miR-532-5p | 0 | | | | | | 0 | 1 | U: 1 | B: 1 | IS: 117; C: 82 | <24 h: 1 | HC: 1 |
| bejleri 2021 | miR-549 | 1 | U: 1 | B: 1 | IS: 169; C: 24 | <24 h: 1 | HC: 1 | 0 | 0 | | | | | |
| bejleri 2021 | miR-574-3p | 0 | | | | | | 0 | 1 | U: 1 | B: 1 | IS: 169; C: 24 | <24 h: 1 | HC: 1 |
| bejleri 2021 | miR-574-5p | 0 | | | | | | 0 | 1 | U: 1 | B: 1 | IS: 169; C: 24 | <24 h: 1 | HC: 1 |
| bejleri 2021 | miR-579-3p | 0 | | | | | | 0 | 1 | U: 1 | B: 1 | IS: 11; C: 10 | <24 h: 1 | HC: 1 |
| bejleri 2021 | miR-617 | 1 | U: 1 | B: 1 | IS: 169; C: 24 | <24 h: 1 | HC: 1 | 0 | 0 | | | | | |
| bejleri 2021 | miR-627 | 1 | U: 1 | B: 1 | IS: 169; C: 24 | <24 h: 1 | HC: 1 | 0 | 0 | | | | | |
| bejleri 2021 | miR-886-5p | 0 | | | | | | 0 | 1 | U: 1 | B: 1 | IS: 169; C: 24 | <24 h: 1 | HC: 1 |
| bejleri 2021 | miR-9 | 0 | | | | | | 0 | 1 | L: 1 | B: 1 | IS: 31; C: 11 | <24 h: 1 | HC: 1 |
| bejleri 2021 | miR-92a | 0 | | | | | | 0 | 1 | U: 1 | B: 1 | IS: 169; C: 24 | <24 h: 1 | HC: 1 |

| | | | | | | | | | | | | | | | | | | | |
|---|---|---|---|---|---|---|---|---|---|---|---|---|---|---|---|---|---|---|---|
| bejleri 2021 | miR-93 | 0 | | | | | | 0 | | | | | | 2 | U: 2 | B: 2 | IS: 202; C: 44 | <6 h: 1; <9 h: 1; <24 h: 2 | HC: 2 |
| bejleri 2021 | miR-Let-7i | 0 | | | | | | 0 | | | | | | 1 | H: 1 | B: 1 | IS: 106; C: 106 | <24 h: 1 | HC: 1 |
| bejleri 2021 | miRNA-335 | 0 | | | | | | 0 | | | | | | 1 | L: 1 | B: 1 | IS: 168; C: 104 | <24 h: 1 | HC: 1 |
| dewdney 2017 | Let-7b | 0 | | | | | | 1 | L: 1 | B: 1 | IS: 38; C: 50 | <24 h: 1 | HC: 1 | 0 | | | | | |
| dewdney 2017 | Let-7e | 1 | U: 1 | B: 1 | IS: 72; C: 51 | <24 h: 1 | HC: 1 | 0 | | | | | | 0 | | | | | |
| dewdney 2017 | miR-106b-5p | 2 | U: 1; H: 1 | B: 2 | IS: 129; C: 166 | <24 h: 2 | HC: 2 | 0 | | | | | | 0 | | | | | |
| dewdney 2017 | miR-1246 | 1 | U: 1 | B: 1 | IS: 53; C: 50 | <24 h: 1 | HC: 1 | 0 | | | | | | 0 | | | | | |
| dewdney 2017 | miR-125b-2 | 1 | H: 1 | B: 1 | IS: 45; C: 24 | <24 h: 1 | HC: 1 | 0 | | | | | | 0 | | | | | |
| dewdney 2017 | miR-126 | 0 | | | | | | 0 | | | | | | 1 | L: 1 | B: 1 | IS: 38; C: 50 | <24 h: 1 | HC: 1 |
| dewdney 2017 | miR-145 | 1 | H: 1 | B: 1 | IS: 146; C: 96 | <24 h: 1 | HC: 1 | 0 | | | | | | 0 | | | | | |
| dewdney 2017 | miR-16 | 1 | U: 1 | B: 1 | IS: 74; C: 23 | <24 h: 1 | HC: 1 | 0 | | | | | | 0 | | | | | |
| dewdney 2017 | miR-21 | 0 | | | | | | 0 | | | | | | 1 | U: 1 | B: 1 | IS: 68; C: 21 | <24 h: 1 | HC: 1 |
| dewdney 2017 | miR-221 | 0 | | | | | | 0 | | | | | | 1 | H: 1 | B: 1 | IS: 146; C: 96 | <24 h: 1 | HC: 1 |
| dewdney 2017 | miR-23a | 0 | | | | | | 0 | | | | | | 1 | H: 1 | B: 1 | IS: 146; C: 96 | <24 h: 1 | HC: 1 |
| dewdney 2017 | miR-24 | 0 | | | | | | 0 | | | | | | 1 | U: 1 | B: 1 | IS: 68; C: 21 | <24 h: 1 | HC: 1 |
| dewdney 2017 | miR-27a | 1 | H: 1 | B: 1 | IS: 45; C: 24 | <24 h: 1 | HC: 1 | 0 | | | | | | 0 | | | | | |
| dewdney 2017 | miR-30a | 0 | | | | | | 0 | | | | | | 1 | L: 1 | B: 1 | IS: 38; C: 50 | <24 h: 1 | HC: 1 |
| dewdney 2017 | miR-32-3p | 1 | U: 1 | B: 1 | IS: 53; C: 50 | <24 h: 1 | HC: 1 | 0 | | | | | | 0 | | | | | |
| dewdney 2017 | miR-320d | 0 | | | | | | 0 | | | | | | 1 | H: 1 | B: 1 | IS: 76; C: 116 | <24 h: 1 | HC: 1 |
| dewdney 2017 | miR-320e | 0 | | | | | | 0 | | | | | | 1 | H: 1 | B: 1 | IS: 76; C: 116 | <24 h: 1 | HC: 1 |
| dewdney 2017 | miR-422a | 1 | H: 1 | B: 1 | IS: 45; C: 24 | <24 h: 1 | HC: 1 | 0 | | | | | | 0 | | | | | |
| dewdney 2017 | miR-4306 | 1 | H: 1 | B: 1 | IS: 76; C: 116 | <24 h: 1 | HC: 1 | 0 | | | | | | 0 | | | | | |
| dewdney 2017 | miR-488 | 1 | H: 1 | B: 1 | IS: 45; C: 24 | <24 h: 1 | HC: 1 | 0 | | | | | | 0 | | | | | |
| dewdney 2017 | miR-532-5p | 0 | | | | | | 0 | | | | | | 1 | U: 1 | B: 1 | IS: 53; C: 50 | <24 h: 1 | HC: 1 |

| | | | | | | | | | | | | | | | | | | | |
|---|---|---|---|---|---|---|---|---|---|---|---|---|---|---|---|---|---|---|---|
| dewdney 2017 | miR-627 | 1 | H: 1 | B: 1 | IS: 45; C: 24 | <24 h: 1 | HC: 1 | 0 | | | | | | 0 | | | | | |
| klokman 2023 | miR-125a-5p | 1 | L: 1 | B: 1 | IS: 23; C: 35 | over: 1 | OC: 1 | 0 | | | | | | 0 | | | | | |
| klokman 2023 | miR-125b-5p | 1 | L: 1 | B: 1 | IS: 23; C: 35 | over: 1 | OC: 1 | 0 | | | | | | 0 | | | | | |
| klokman 2023 | miR-143-3p | 1 | L: 1 | B: 1 | IS: 23; C: 35 | over: 1 | OC: 1 | 0 | | | | | | 0 | | | | | |
| klokman 2023 | miR-342-3p | 0 | | | | | | 1 | L: 1 | B: 1 | IS: 23; C: 35 | over: 1 | OC: 1 | 0 | | | | | |
| klokman 2023 | miR-376a-3p | 0 | | | | | | 1 | L: 1 | B: 1 | IS: 23; C: 35 | over: 1 | OC: 1 | 0 | | | | | |
| klokman 2023 | miR-433-5p | 1 | L: 1 | B: 1 | IS: 23; C: 35 | over: 1 | OC: 1 | 0 | | | | | | 0 | | | | | |
| loggini 2024 | miR-1255b-5p | 0 | | | | | | 0 | | | | | | 1 | U: 1 | B: 1 | IS: 22; C: 22 | <24 h: 1 | HC: 1 |
| loggini 2024 | miR-125a-5p | 1 | L: 1 | B: 1 | IS: 200; C: 100 | <24 h: 1 | HC: 1 | 0 | | | | | | 0 | | | | | |
| loggini 2024 | miR-125b-5p | 1 | L: 1 | B: 1 | IS: 200; C: 100 | <24 h: 1 | HC: 1 | 0 | | | | | | 0 | | | | | |
| loggini 2024 | miR-143-3p | 1 | L: 1 | B: 1 | IS: 200; C: 100 | <24 h: 1 | HC: 1 | 0 | | | | | | 0 | | | | | |
| loggini 2024 | miR-30a-5p | 1 | L: 1 | B: 1 | IS: 15; C: 24 | <6 h: 1; <9 h: 1; <24 h: 1 | HC: 1 | 0 | | | | | | 0 | | | | | |
| loggini 2024 | miR-4523 | 0 | | | | | | 0 | | | | | | 1 | U: 1 | B: 1 | IS: 22; C: 22 | <24 h: 1 | HC: 1 |
| loggini 2024 | miR-505â€“5p | 1 | U: 1 | B: 1 | IS: 22; C: 22 | <24 h: 1 | HC: 1 | 0 | | | | | | 0 | | | | | |
| loggini 2024 | miR-550b-2-5p | 0 | | | | | | 0 | | | | | | 1 | U: 1 | B: 1 | IS: 22; C: 22 | <24 h: 1 | HC: 1 |
| loggini 2024 | miR-6795-3p | 0 | | | | | | 0 | | | | | | 1 | U: 1 | B: 1 | IS: 22; C: 22 | <24 h: 1 | HC: 1 |
| loggini 2024 | miRNA-221-3p | 0 | | | | | | 0 | | | | | | 1 | L: 1 | B: 1 | IS: 78; C: 39 | <6 h: 1; <9 h: 1; <24 h: 1 | HC: 1 |
| loggini 2024 | miRNA-382-5p | 0 | | | | | | 0 | | | | | | 1 | L: 1 | B: 1 | IS: 78; C: 39 | <6 h: 1; <9 h: 1; <24 h: 1 | HC: 1 |
| naik 2023 | Let-7c | 1 | L: 1 | CSF: 1 | IS: 10; C: 10 | over: 1 | HC: 1 | 0 | | | | | | 0 | | | | | |
| naik 2023 | Let-7e | 1 | L: 1 | CSF: 1 | IS: 28; C: 12 | over: 1 | HC: 1 | 0 | | | | | | 0 | | | | | |
| naik 2023 | miR-221-3p | 1 | L: 1 | CSF: 1 | IS: 10; C: 10 | over: 1 | HC: 1 | 0 | | | | | | 0 | | | | | |
| naik 2023 | miR-338 | 1 | L: 1 | CSF: 1 | IS: 9; C: 12 | over: 1 | HC: 1 | 0 | | | | | | 0 | | | | | |
| naik 2023 | miR-523-3p | 1 | L: 1 | CSF: 1 | IS: 10; C: 10 | over: 1 | HC: 1 | 0 | | | | | | 0 | | | | | |
| wang 2023 | Let-7e-5p | 2 | | B: 2 | IS: 312; C: 313 | <24 h: 2 | HC: 2 | 0 | | | | | | 0 | | | | | |

| | | | | | | | | | | | | |
|---|---|---|---|---|---|---|---|---|---|---|---|---|
| wang 2023 | Let-7f-5p | 0 | | | | | 0 | 1 | B: 1 | IS: 28; C: 35 | <24 h: 1 | HC: 1 |
| wang 2023 | Let-7i-5p | 0 | | | | | 0 | 1 | Ce: 1 | IS: 24; C: 24 | <24 h: 1 | HC: 1 |
| wang 2023 | miR-106b-5p | 1 | B: 1 | IS: 136; C: 116 | <24 h: 1 | HC: 1 | 0 | 0 | | | | |
| wang 2023 | miR-122-5p | 1 | B: 1 | IS: 34; C: 11 | <24 h: 1 | HC: 1 | 0 | 1 | Ce: 1 | IS: 24; C: 24 | <24 h: 1 | HC: 1 |
| wang 2023 | miR-1229-3p | 1 | B: 1 | IS: 10; C: 11 | <24 h: 1 | HC: 1 | 0 | 0 | | | | |
| wang 2023 | miR-1238-5p | 1 | B: 1 | IS: 10; C: 11 | <24 h: 1 | HC: 1 | 0 | 0 | | | | |
| wang 2023 | miR-124-3p | 0 | | | | | 0 | 2 | B: 2 | IS: 41; C: 21 | <24 h: 2 | HC: 2 |
| wang 2023 | miR-1246 | 1 | B: 1 | IS: 117; C: 82 | <24 h: 1 | HC: 1 | 0 | 0 | | | | |
| wang 2023 | miR-126-3p | 0 | | | | | 0 | 5 | B: 5 | IS: 324; C: 330 | <24 h: 5 | HC: 5 |
| wang 2023 | miR-1270 | 1 | B: 1 | IS: 10; C: 11 | <24 h: 1 | HC: 1 | 0 | 0 | | | | |
| wang 2023 | miR-1275 | 1 | B: 1 | IS: 279; C: 279 | <24 h: 1 | HC: 1 | 0 | 0 | | | | |
| wang 2023 | miR-128-3p | 1 | B: 1 | IS: 80; C: 60 | <24 h: 1 | HC: 1 | 0 | 0 | | | | |
| wang 2023 | miR-1294 | 1 | B: 1 | IS: 10; C: 11 | <24 h: 1 | HC: 1 | 0 | 0 | | | | |
| wang 2023 | miR-1299 | 1 | B: 1 | IS: 117; C: 82 | <24 h: 1 | HC: 1 | 0 | 0 | | | | |
| wang 2023 | miR-1301-3p | 1 | B: 1 | IS: 10; C: 11 | <24 h: 1 | HC: 1 | 0 | 0 | | | | |
| wang 2023 | miR-130a-3p | 1 | Ce: 1 | IS: 19; C: 20 | <24 h: 1 | HC: 1 | 0 | 2 | B: 2 | IS: 254; C: 258 | <24 h: 2 | HC: 2 |
| wang 2023 | miR-140-5p | 0 | | | | | 0 | 1 | B: 1 | IS: 10; C: 11 | <24 h: 1 | HC: 1 |
| wang 2023 | miR-142-3p | 0 | | | | | 0 | 2 | B: 2 | IS: 57; C: 107 | <24 h: 2 | HC: 2 |
| wang 2023 | miR-144-3p | 1 | B: 1 | IS: 19; C: 5 | <24 h: 1 | HC: 1 | 0 | 1 | B: 1 | IS: 10; C: 11 | <24 h: 1 | HC: 1 |
| wang 2023 | miR-145-5p | 1 | B: 1 | IS: 146; C: 96 | <24 h: 1 | HC: 1 | 0 | 0 | | | | |
| wang 2023 | miR-146a-5p | 0 | | | | | 0 | 2 | B: 2 | IS: 104; C: 52 | <24 h: 2 | HC: 2 |
| wang 2023 | miR-146b-5p | 1 | B: 1 | IS: 128; C: 102 | <24 h: 1 | HC: 1 | 0 | 0 | | | | |
| wang 2023 | miR-148a-3p | 0 | | | | | 0 | 1 | Ce: 1 | IS: 24; C: 24 | <24 h: 1 | HC: 1 |
| wang 2023 | miR-149-5p | 0 | | | | | 0 | 1 | B: 1 | IS: 47; C: 96 | <24 h: 1 | HC: 1 |

| | | | | | | | | | | | | | |
|---|---|---|---|---|---|---|---|---|---|---|---|---|---|
| wang 2023 | miR-15a-5p | 1 | B: 1 | IS: 106; C: 120 | <24 h: 1 | HC: 1 | 0 | | 0 | | | | |
| wang 2023 | miR-16-5p | 2 | B: 2 | IS: 125; C: 125 | <24 h: 2 | HC: 2 | 0 | | 0 | | | | |
| wang 2023 | miR-17-5p | 2 | B: 2 | IS: 245; C: 154 | <24 h: 2 | HC: 2 | 0 | | 0 | | | | |
| wang 2023 | miR-185-5p | 3 | B: 3 | IS: 396; C: 308 | <24 h: 3 | HC: 3 | 0 | | 1 | B: 1 | IS: 60; C: 30 | <24 h: 1 | HC: 1 |
| wang 2023 | miR-186-5p | 0 | | | | | 0 | | 1 | B: 1 | IS: 10; C: 11 | <24 h: 1 | HC: 1 |
| wang 2023 | miR-18b-5p | 0 | | | | | 0 | | 1 | B: 1 | IS: 10; C: 11 | <24 h: 1 | HC: 1 |
| wang 2023 | miR-1913 | 0 | | | | | 0 | | 1 | B: 1 | IS: 117; C: 82 | <24 h: 1 | HC: 1 |
| wang 2023 | miR-195-5p | 1 | B: 1 | IS: 18; C: 20 | <24 h: 1 | HC: 1 | 0 | | 0 | | | | |
| wang 2023 | miR-19a-3p | 1 | B: 1 | IS: 28; C: 35 | <24 h: 1 | HC: 1 | 0 | | 2 | B: 1; Ce: 1 | IS: 34; C: 35 | <24 h: 2 | HC: 2 |
| wang 2023 | miR-20b-5p | 1 | B: 1 | IS: 139; C: 34 | <24 h: 1 | HC: 1 | 0 | | 0 | | | | |
| wang 2023 | miR-21-5p | 1 | B: 1 | IS: 19; C: 5 | <24 h: 1 | HC: 1 | 0 | | 0 | | | | |
| wang 2023 | miR-210-3p | 0 | | | | | 0 | | 1 | B: 1 | IS: 112; C: 60 | <24 h: 1 | HC: 1 |
| wang 2023 | miR-211-5p | 1 | B: 1 | IS: 34; C: 11 | <24 h: 1 | HC: 1 | 0 | | 0 | | | | |
| wang 2023 | miR-218-5p | 1 | B: 1 | IS: 106; C: 110 | <24 h: 1 | HC: 1 | 0 | | 1 | B: 1 | IS: 10; C: 10 | <24 h: 1 | HC: 1 |
| wang 2023 | miR-219-5p | 1 | B: 1 | IS: 148; C: 148 | <24 h: 1 | HC: 1 | 0 | | 0 | | | | |
| wang 2023 | miR-22-3p | 1 | B: 1 | IS: 10; C: 10 | <24 h: 1 | HC: 1 | 0 | | 1 | B: 1 | IS: 34; C: 11 | <24 h: 1 | HC: 1 |
| wang 2023 | miR-221-3p | 0 | | | | | 0 | | 2 | B: 2 | IS: 214; C: 135 | <24 h: 2 | HC: 2 |
| wang 2023 | miR-222-3p | 2 | B: 2 | IS: 254; C: 258 | <24 h: 2 | HC: 2 | 0 | | 0 | | | | |
| wang 2023 | miR-223-3p | 1 | B: 1 | IS: 19; C: 5 | <24 h: 1 | HC: 1 | 0 | | 0 | | | | |
| wang 2023 | miR-224-3p | 0 | | | | | 0 | | 1 | B: 1 | IS: 117; C: 82 | <24 h: 1 | HC: 1 |
| wang 2023 | miR-23a-3p | 1 | B: 1 | IS: 10; C: 10 | <24 h: 1 | HC: 1 | 0 | | 2 | B: 2 | IS: 180; C: 107 | <24 h: 2 | HC: 2 |
| wang 2023 | miR-27b-3p | 1 | B: 1 | IS: 139; C: 34 | <24 h: 1 | HC: 1 | 0 | | 0 | | | | |
| wang 2023 | miR-301a-3p | 0 | | | | | 0 | | 1 | B: 1 | IS: 10; C: 11 | <24 h: 1 | HC: 1 |
| wang 2023 | miR-30a-5p | 0 | | | | | 0 | | 2 | B: 2 | IS: 48; C: 60 | <24 h: 2 | HC: 2 |

| | | | | | | | | | | | | |
|---|---|---|---|---|---|---|---|---|---|---|---|---|
| wang 2023 | miR-30d-5p | 0 | | | | | 0 | 1 | B: 1 | IS: 34; C: 11 | <24 h: 1 | HC: 1 |
| wang 2023 | miR-3149 | 1 | B: 1 | IS: 117; C: 82 | <24 h: 1 | HC: 1 | 0 | 0 | | | | |
| wang 2023 | miR-32-3p | 1 | B: 1 | IS: 117; C: 82 | <24 h: 1 | HC: 1 | 0 | 0 | | | | |
| wang 2023 | miR-32-5p | 0 | | | | | 0 | 1 | B: 1 | IS: 10; C: 11 | <24 h: 1 | HC: 1 |
| wang 2023 | miR-320a | 1 | Ce: 1 | IS: 19; C: 20 | <24 h: 1 | HC: 1 | 0 | 0 | | | | |
| wang 2023 | miR-320a-3p | 1 | B: 1 | IS: 19; C: 5 | <24 h: 1 | HC: 1 | 0 | 0 | | | | |
| wang 2023 | miR-320d | 0 | | | | | 0 | 2 | B: 1; Ce: 1 | IS: 160; C: 140 | <24 h: 2 | HC: 2 |
| wang 2023 | miR-320e | 0 | | | | | 0 | 1 | B: 1 | IS: 136; C: 116 | <24 h: 1 | HC: 1 |
| wang 2023 | miR-330-3p | 0 | | | | | 0 | 1 | B: 1 | IS: 10; C: 10 | <24 h: 1 | HC: 1 |
| wang 2023 | miR-335-5p | 0 | | | | | 0 | 1 | B: 1 | IS: 10; C: 11 | <24 h: 1 | HC: 1 |
| wang 2023 | miR-33a-5p | 0 | | | | | 0 | 1 | B: 1 | IS: 10; C: 10 | <24 h: 1 | HC: 1 |
| wang 2023 | miR-340-5p | 0 | | | | | 0 | 1 | B: 1 | IS: 10; C: 11 | <24 h: 1 | HC: 1 |
| wang 2023 | miR-34a-5p | 1 | B: 1 | IS: 102; C: 97 | <24 h: 1 | HC: 1 | 0 | 0 | | | | |
| wang 2023 | miR-362-3p | 0 | | | | | 0 | 1 | B: 1 | IS: 10; C: 11 | <24 h: 1 | HC: 1 |
| wang 2023 | miR-363-3p | 1 | Ce: 1 | IS: 24; C: 24 | <24 h: 1 | HC: 1 | 0 | 0 | | | | |
| wang 2023 | miR-376c-3p | 1 | Ce: 1 | IS: 19; C: 20 | <24 h: 1 | HC: 1 | 0 | 0 | | | | |
| wang 2023 | miR-377-5p | 0 | | | | | 0 | 1 | B: 1 | IS: 117; C: 82 | <24 h: 1 | HC: 1 |
| wang 2023 | miR-378a-5p | 0 | | | | | 0 | 1 | B: 1 | IS: 106; C: 110 | <24 h: 1 | HC: 1 |
| wang 2023 | miR-379-5p | 0 | | | | | 0 | 1 | B: 1 | IS: 47; C: 96 | <24 h: 1 | HC: 1 |
| wang 2023 | miR-382-5p | 0 | | | | | 0 | 1 | B: 1 | IS: 68; C: 39 | <24 h: 1 | HC: 1 |
| wang 2023 | miR-411-5p | 0 | | | | | 0 | 1 | B: 1 | IS: 47; C: 96 | <24 h: 1 | HC: 1 |
| wang 2023 | miR-423-5p | 1 | B: 1 | IS: 117; C: 82 | <24 h: 1 | HC: 1 | 0 | 0 | | | | |
| wang 2023 | miR-424-5p | 1 | B: 1 | IS: 142; C: 50 | <24 h: 1 | HC: 1 | 0 | 0 | | | | |
| wang 2023 | miR-4306 | 1 | B: 1 | IS: 136; C: 116 | <24 h: 1 | HC: 1 | 0 | 0 | | | | |

| | | | | | | | | | | | | |
|---|---|---|---|---|---|---|---|---|---|---|---|---|
| wang 2023 | miR-432-5p | 1 | Ce: 1 | IS: 19; C: 20 | <24 h: 1 | HC: 1 | 0 | 0 | | | | |
| wang 2023 | miR-4429 | 0 | | | | | 0 | 1 | Ce: 1 | IS: 24; C: 24 | <24 h: 1 | HC: 1 |
| wang 2023 | miR-4446-3p | 0 | | | | | 0 | 1 | B: 1 | IS: 47; C: 96 | <24 h: 1 | HC: 1 |
| wang 2023 | miR-451a | 3 | B: 3 | IS: 182; C: 198 | <24 h: 3 | HC: 3 | 0 | 0 | | | | |
| wang 2023 | miR-4656 | 1 | Ce: 1 | IS: 19; C: 20 | <24 h: 1 | HC: 1 | 0 | 0 | | | | |
| wang 2023 | miR-4739 | 1 | B: 1 | IS: 117; C: 82 | <24 h: 1 | HC: 1 | 0 | 0 | | | | |
| wang 2023 | miR-485-3p | 0 | | | | | 0 | 1 | B: 1 | IS: 47; C: 96 | <24 h: 1 | HC: 1 |
| wang 2023 | miR-487 | 1 | Ce: 1 | IS: 19; C: 20 | <24 h: 1 | HC: 1 | 0 | 0 | | | | |
| wang 2023 | miR-487b-3p | 1 | Ce: 1 | IS: 24; C: 24 | <24 h: 1 | HC: 1 | 0 | 0 | | | | |
| wang 2023 | miR-503-5p | 1 | Ce: 1 | IS: 19; C: 20 | <24 h: 1 | HC: 1 | 0 | 0 | | | | |
| wang 2023 | miR-505-5p | 1 | B: 1 | IS: 10; C: 11 | <24 h: 1 | HC: 1 | 0 | 0 | | | | |
| wang 2023 | miR-517b-3p | 0 | | | | | 0 | 1 | B: 1 | IS: 10; C: 11 | <24 h: 1 | HC: 1 |
| wang 2023 | miR-518b | 0 | | | | | 0 | 1 | B: 1 | IS: 117; C: 82 | <24 h: 1 | HC: 1 |
| wang 2023 | miR-532-5p | 0 | | | | | 0 | 1 | B: 1 | IS: 117; C: 82 | <24 h: 1 | HC: 1 |
| wang 2023 | miR-544a | 1 | B: 1 | IS: 10; C: 11 | <24 h: 1 | HC: 1 | 0 | 0 | | | | |
| wang 2023 | miR-574-5p | 0 | | | | | 0 | 1 | B: 1 | IS: 47; C: 96 | <24 h: 1 | HC: 1 |
| wang 2023 | miR-579-3p | 0 | | | | | 0 | 1 | B: 1 | IS: 10; C: 11 | <24 h: 1 | HC: 1 |
| wang 2023 | miR-628-5p | 1 | B: 1 | IS: 10; C: 11 | <24 h: 1 | HC: 1 | 0 | 0 | | | | |
| wang 2023 | miR-660-5p | 0 | | | | | 0 | 1 | B: 1 | IS: 10; C: 11 | <24 h: 1 | HC: 1 |
| wang 2023 | miR-664a-5p | 1 | B: 1 | IS: 10; C: 11 | <24 h: 1 | HC: 1 | 0 | 0 | | | | |
| wang 2023 | miR-6721-5p | 0 | | | | | 0 | 1 | B: 1 | IS: 47; C: 96 | <24 h: 1 | HC: 1 |
| wang 2023 | miR-676-3p | 0 | | | | | 0 | 1 | B: 1 | IS: 47; C: 96 | <24 h: 1 | HC: 1 |
| wang 2023 | miR-874-3p | 0 | | | | | 0 | 1 | Ce: 1 | IS: 19; C: 20 | <24 h: 1 | HC: 1 |
| wang 2023 | miR-877-5p | 1 | B: 1 | IS: 10; C: 11 | <24 h: 1 | HC: 1 | 0 | 0 | | | | |

| | | | | | | | | | | | | | | |
|---|---|---|---|---|---|---|---|---|---|---|---|---|---|---|
| wang 2023 | miR-9-5p | 0 | | | | | | 0 | 2 | | B: 2 | IS: 41; C: 21 | <24 h: 2 | HC: 2 |
| wang 2023 | miR-93-5p | 1 | | B: 1 | IS: 139; C: 34 | <24 h: 1 | HC: 1 | 0 | 0 | | | | | |
| zhang 2025 | Let-7e | 1 | U: 1 | B: 1 | IS: 51; C: 33 | <24 h: 1 | HC: 1 | 0 | 0 | | | | | |
| zhang 2025 | miR-106b-5p | 1 | U: 1 | B: 1 | IS: 76; C: 116 | <24 h: 1 | HC: 1 | 0 | 0 | | | | | |
| zhang 2025 | miR-124 | 0 | | | | | | 0 | 1 | U: 1 | B: 1 | IS: 108; C: 108 | <24 h: 1 | HC: 1 |
| zhang 2025 | miR-125a-5p | 1 | L: 1 | B: 1 | IS: 200; C: 100 | <24 h: 1 | HC: 1 | 0 | 0 | | | | | |
| zhang 2025 | miR-125b | 1 | U: 1 | B: 1 | IS: 51; C: 33 | <24 h: 1 | HC: 1 | 0 | 0 | | | | | |
| zhang 2025 | miR-125b-5p | 1 | L: 1 | B: 1 | IS: 200; C: 100 | <24 h: 1 | HC: 1 | 0 | 0 | | | | | |
| zhang 2025 | miR-143-3p | 1 | L: 1 | B: 1 | IS: 200; C: 100 | <24 h: 1 | HC: 1 | 0 | 0 | | | | | |
| zhang 2025 | miR-155 | 1 | U: 1 | B: 1 | IS: 46; C: 50 | <24 h: 1 | HC: 1 | 0 | 0 | | | | | |
| zhang 2025 | miR-16 | 2 | U: 2 | B: 2 | IS: 13; C: 46 | <6 h: 2; <9 h: 2; <24 h: 2 | HC: 2 | 0 | 0 | | | | | |
| zhang 2025 | miR-320d | 0 | | | | | | 0 | 1 | U: 1 | B: 1 | IS: 76; C: 116 | <24 h: 1 | HC: 1 |
| zhang 2025 | miR-320e | 0 | | | | | | 0 | 1 | U: 1 | B: 1 | IS: 76; C: 116 | <24 h: 1 | HC: 1 |
| zhang 2025 | miR-4306 | 1 | U: 1 | B: 1 | IS: 76; C: 116 | <24 h: 1 | HC: 1 | 0 | 0 | | | | | |

**Table S3. Characteristics of the included meta-analyses for CircRNA biomarkers**

| | | Higher in Stroke | | | | | | Lower in Stroke | | | | | |
|---|---|---|---|---|---|---|---|---|---|---|---|---|---|
| **Review** | **Biomarker** | **n** | **R** | **M** | **Subjects** | **Time to onset** | **Ctrl** | **n** | **R** | **M** | **Subjects** | **Time to onset** | **Ctrl** |
| loggini 2024 | Circ_0001460 | 1 | U: 1 | Ce: 1 | IS: 118; C: 118 | <24 h: 1 | HC: 1 | 0 | | | | | |
| loggini 2024 | Circ_0001599 | 0 | | | | | | 1 | U: 1 | Ce: 1 | IS: 118; C: 118 | <24 h: 1 | HC: 1 |
| loggini 2024 | Circ_0004338 | 0 | | | | | | 1 | U: 1 | Ce: 1 | IS: 118; C: 118 | <24 h: 1 | HC: 1 |
| loggini 2024 | Circ_0007637 | 0 | | | | | | 1 | U: 1 | Ce: 1 | IS: 118; C: 118 | <24 h: 1 | HC: 1 |
| loggini 2024 | Circ_0073239 | 0 | | | | | | 1 | U: 1 | Ce: 1 | IS: 118; C: 118 | <24 h: 1 | HC: 1 |
| zhang 2023 | Circ_0000097 | 1 | H: 1 | B: 1 | IS: 200; C: 100 | over: 1 | HC: 1 | 0 | | | | | |
| zhang 2023 | Circ_0000607 | 0 | | | | | | 1 | U: 1 | B: 1 | IS: 32; C: 32 | over: 1 | HC: 1 |
| zhang 2023 | Circ_0000698 | 0 | | | | | | 1 | H: 1 | B: 1 | IS: 32; C: 32 | over: 1 | HC: 1 |
| zhang 2023 | Circ_0001460 | 1 | H: 1 | Ce: 1 | IS: 168; C: 118 | <4.5 h: 1; <6 h: 1; <9 h: 1; <24 h: 1 | HC: 1 | 0 | | | | | |
| zhang 2023 | Circ_0001599 | 1 | H: 1 | Ce: 1 | IS: 168; C: 118 | <4.5 h: 1; <6 h: 1; <9 h: 1; <24 h: 1 | HC: 1 | 0 | | | | | |
| zhang 2023 | Circ_0002465 | 0 | | | | | | 1 | U: 1 | B: 1 | IS: 32; C: 32 | over: 1 | HC: 1 |
| zhang 2023 | Circ_0004338 | 1 | H: 1 | Ce: 1 | IS: 168; C: 118 | <4.5 h: 1; <6 h: 1; <9 h: 1; <24 h: 1 | HC: 1 | 0 | | | | | |
| zhang 2023 | Circ_0004494 | 1 | H: 1 | B: 1 | IS: 200; C: 100 | over: 1 | HC: 1 | 0 | | | | | |
| zhang 2023 | Circ_0005548 | 1 | U: 1 | B: 1 | IS: 32; C: 32 | over: 1 | HC: 1 | 0 | | | | | |
| zhang 2023 | Circ_0005585 | 0 | | | | | | 1 | H: 1 | B: 1 | IS: 32; C: 32 | over: 1 | HC: 1 |
| zhang 2023 | Circ_0006911 | 0 | | | | | | 1 | H: 1 | Ce: 1 | IS: 168; C: 118 | <4.5 h: 1; <6 h: 1; <9 h: 1; <24 h: 1 | HC: 1 |
| zhang 2023 | Circ_0007290 | 1 | H: 1 | B: 1 | IS: 200; C: 100 | over: 1 | HC: 1 | 0 | | | | | |
| zhang 2023 | Circ_0007637 | 0 | | | | | | 1 | H: 1 | Ce: 1 | IS: 168; C: 118 | <4.5 h: 1; <6 h: 1; <9 h: 1; <24 h: 1 | HC: 1 |
| zhang 2023 | Circ_0010155 | 0 | | | | | | 1 | H: 1 | B: 1 | IS: 32; C: 32 | over: 1 | HC: 1 |
| zhang 2023 | Circ_0039457 | 0 | | | | | | 1 | | B: 1 | IS: 8; C: 8 | <4.5 h: 1; <6 h: 1; <9 h: 1; <24 h: 1 | HC: 1 |
| zhang 2023 | Circ_0043837 | 0 | | | | | | 1 | H: 1 | B: 1 | IS: 32; C: 32 | over: 1 | HC: 1 |
| zhang 2023 | Circ_0072309 | 0 | | | | | | 1 | H: 1 | B: 1 | IS: 90; C: 75 | <24 h: 1 | HC: 1 |
| zhang 2023 | Circ_0090002 | 0 | | | | | | 1 | | B: 1 | IS: 8; C: 8 | <4.5 h: 1; <6 h: 1; <9 h: 1; <24 h: 1 | HC: 1 |
| zhang 2023 | Circ_0141720 | 1 | L: 1 | B: 1 | IS: 80; C: 30 | over: 1 | HC: 1 | 0 | | | | | |
| zhang 2023 | circRNA CCZ1 | 1 | H: 1 | B: 1 | IS: 145; C: 145 | over: 1 | HC: 1 | 0 | | | | | |
| zhang 2023 | circRNA CDC14A | 1 | H: 1 | B: 1 | IS: 145; C: 145 | over: 1 | HC: 1 | 0 | | | | | |
| zhang 2023 | circRNA CDC42BPA | 1 | H: 1 | B: 1 | IS: 145; C: 145 | over: 1 | HC: 1 | 0 | | | | | |
| zhang 2023 | circRNA CEP78 | 0 | | | | | | 1 | | Ce: 1 | IS: 5; C: 5 | over: 1 | HC: 1 |
| zhang 2023 | circRNA DAB1 | 0 | | | | | | 1 | | Ce: 1 | IS: 5; C: 5 | over: 1 | HC: 1 |

| | | | | | | | | | | | | | |
|---|---|---|---|---|---|---|---|---|---|---|---|---|---|
| zhang 2023 | circRNA DLGAP4 | 0 | | | | | | 1 | U: 1 | Ce: 1 | IS: 170; C: 170 | <24 h: 1 | HC: 1 |
| zhang 2023 | circRNA FBXW4 | 0 | | | | | | 1 | H: 1 | B: 1 | IS: 145; C: 145 | over: 1 | HC: 1 |
| zhang 2023 | circRNA GNB2L1 | 0 | | | | | | 1 | H: 1 | B: 1 | IS: 145; C: 145 | over: 1 | HC: 1 |
| zhang 2023 | circRNA HECTD1 | 2 | U: 1 | B: 1; Ce: 1 | IS: 197; C: 194 | <24 h: 2 | HC: 2 | 0 | | | | | |
| zhang 2023 | circRNA KLRK1 | 0 | | | | | | 1 | | Ce: 1 | IS: 5; C: 5 | over: 1 | HC: 1 |
| zhang 2023 | circRNA PDE4B | 0 | | | | | | 1 | H: 1 | B: 1 | IS: 145; C: 145 | over: 1 | HC: 1 |
| zhang 2023 | circRNA PIGB | 1 | | Ce: 1 | IS: 5; C: 5 | over: 1 | HC: 1 | 0 | | | | | |
| zhang 2023 | circRNA PLXNC1 | 1 | | Ce: 1 | IS: 5; C: 5 | over: 1 | HC: 1 | 0 | | | | | |
| zhang 2023 | circRNA RAP1B | 0 | | | | | | 1 | | Ce: 1 | IS: 5; C: 5 | over: 1 | HC: 1 |
| zhang 2023 | circRNA RBM33 | 1 | H: 1 | B: 1 | IS: 145; C: 145 | over: 1 | HC: 1 | 0 | | | | | |
| zhang 2023 | circRNA SCMH1 | 0 | | | | | | 1 | H: 1 | B: 1 | IS: 145; C: 145 | over: 1 | HC: 1 |
| zhang 2023 | circRNA TLK1 | 1 | U: 1 | B: 1 | IS: 71; C: 71 | <24 h: 1 | HC: 1 | 0 | | | | | |
| zhang 2023 | circRNA TMEM56 | 1 | | Ce: 1 | IS: 5; C: 5 | over: 1 | HC: 1 | 0 | | | | | |
| zhang 2023 | circRNA TREM1 | 1 | | Ce: 1 | IS: 5; C: 5 | over: 1 | HC: 1 | 0 | | | | | |
| zhang 2023 | circRNA ZCCHC11 | 1 | H: 1 | B: 1 | IS: 145; C: 145 | over: 1 | HC: 1 | 0 | | | | | |
| zhang 2025 | circOGDH | 1 | U: 1 | B: 1 | IS: 45; C: 32 | <24 h: 1 | HC: 1 | 0 | | | | | |

**Table S4. Characteristics of the included meta-analyses for lncRNA biomarkers**

| | | Higher in Stroke | | | | | | Lower in Stroke | | | | | |
|---|---|---|---|---|---|---|---|---|---|---|---|---|---|
| **Review** | **Biomarker** | **n** | **R** | **M** | **Subjects** | **Time to onset** | **Ctrl** | **n** | **R** | **M** | **Subjects** | **Time to onset** | **Ctrl** |
| pan 2024 | ADAMTS9-AS2 | 0 | | | | | | 1 | | B: 1 | IS: 43; C: 68 | over: 1 | HC: 1 |
| pan 2024 | NBAT1 | 1 | H: 1 | B: 1 | IS: 60; C: 60 | over: 1 | HC: 1 | 0 | | | | | |
| pan 2024 | SNHG15 | 1 | | B: 1 | IS: 30; C: 30 | over: 1 | HC: 1 | 0 | | | | | |
| pan 2024 | SNHG5 | 1 | | B: 1 | IS: 10; C: 10 | over: 1 | HC: 1 | 0 | | | | | |
| pan 2024 | TUG1 | 1 | H: 1 | B: 1 | IS: 60; C: 60 | over: 1 | HC: 1 | 0 | | | | | |
| pan 2024 | lncRNA AC136007.2 | 0 | | | | | | 1 | H: 1 | B: 1 | IS: 32; C: 32 | over: 1 | HC: 1 |
| pan 2024 | lncRNA AK139328 | 1 | | B: 1 | IS: 30; C: 30 | over: 1 | HC: 1 | 0 | | | | | |
| pan 2024 | lncRNA ANRIL | 3 | H: 2 | B: 3 | IS: 297; C: 278 | over: 3 | HC: 3 | 1 | U: 1 | B: 1 | IS: 126; C: 125 | over: 1 | HC: 1 |
| pan 2024 | lncRNA C14orf64 | 0 | | | | | | 1 | H: 1 | B: 1 | IS: 32; C: 32 | over: 1 | HC: 1 |
| pan 2024 | lncRNA CASC15 | 1 | H: 1 | B: 1 | IS: 98; C: 85 | over: 1 | HC: 1 | 0 | | | | | |
| pan 2024 | lncRNA DHFRL1-4 | 1 | | Ce: 1 | IS: 32; C: 32 | over: 1 | HC: 1 | 0 | | | | | |
| pan 2024 | lncRNA ENST00000530525 | 0 | | | | | | 1 | | B: 1 | IS: 5; C: 5 | over: 1 | HC: 1 |
| pan 2024 | lncRNA ENST00000568297 | 1 | H: 1 | B: 1 | IS: 50; C: 50 | over: 1 | HC: 1 | 0 | | | | | |
| pan 2024 | lncRNA FAM98A-3 | 1 | | B: 1 | IS: 30; C: 30 | over: 1 | HC: 1 | 0 | | | | | |
| pan 2024 | lncRNA GAS5 | 3 | L: 1; U: 1 | B: 2; Ce: 1 | IS: 268; C: 205 | over: 3 | HC: 3 | 0 | | | | | |
| pan 2024 | lncRNA H19 | 6 | U: 2; H: 1 | B: 6 | IS: 395; C: 314 | over: 6 | HC: 6 | 0 | | | | | |
| pan 2024 | lncRNA HULC | 1 | U: 1 | B: 1 | IS: 215; C: 215 | over: 1 | HC: 1 | 0 | | | | | |
| pan 2024 | lncRNA ITSN1-2 | 2 | | B: 1; Ce: 1 | IS: 422; C: 370 | over: 2 | HC: 2 | 0 | | | | | |
| pan 2024 | lncRNA KCNQ1OT1 | 1 | | B: 1 | IS: 42; C: 40 | over: 1 | HC: 1 | 0 | | | | | |
| pan 2024 | lncRNA MALAT1 | 0 | | | | | | 2 | H: 1 | B: 2 | IS: 220; C: 220 | over: 2 | HC: 2 |
| pan 2024 | lncRNA MEG3 | 1 | | Ce: 1 | IS: 170; C: 100 | over: 1 | HC: 1 | 0 | | | | | |
| pan 2024 | lncRNA MIAT | 1 | U: 1 | B: 1 | IS: 232; C: 232 | over: 1 | HC: 1 | 1 | U: 1 | B: 1 | IS: 80; C: 40 | over: 1 | HC: 1 |
| pan 2024 | lncRNA NEAT1 | 1 | U: 1 | B: 1 | IS: 210; C: 210 | over: 1 | HC: 1 | 0 | | | | | |
| pan 2024 | lncRNA NORAD | 1 | H: 1 | B: 1 | IS: 103; C: 95 | over: 1 | HC: 1 | 0 | | | | | |
| pan 2024 | lncRNA NR_046084 | 1 | H: 1 | B: 1 | IS: 50; C: 50 | over: 1 | HC: 1 | 0 | | | | | |
| pan 2024 | lncRNA OIP5-AS1 | 0 | | | | | | 1 | | B: 1 | IS: 15; C: 15 | over: 1 | HC: 1 |
| pan 2024 | lncRNA PVT1 | 2 | H: 1 | B: 2 | IS: 140; C: 140 | over: 2 | HC: 2 | 0 | | | | | |
| pan 2024 | lncRNA SERPINB9P1 | 0 | | | | | | 1 | | B: 1 | IS: 159; C: 153 | over: 1 | HC: 1 |
| pan 2024 | lncRNA SNHG16 | 0 | | | | | | 1 | | Ce: 1 | IS: 120; C: 60 | over: 1 | HC: 1 |

| | | | | | | | | | | | | | |
|---|---|---|---|---|---|---|---|---|---|---|---|---|---|
| pan 2024 | lncRNA TUG1 | 1 | | B: 1 | IS: 46; C: 46 | over: 1 | HC: 1 | 0 | | | | | |
| pan 2024 | lncRNA UCA1 | 1 | | Ce: 1 | IS: 160; C: 160 | over: 1 | HC: 1 | 0 | | | | | |
| pan 2024 | lncRNA WT1-as | 0 | | | | | | 1 | | B: 1 | IS: 30; C: 30 | over: 1 | HC: 1 |
| pan 2024 | lncRNA XIST | 0 | | | | | | 1 | | B: 1 | IS: 77; C: 60 | over: 1 | HC: 1 |
| pan 2024 | lncRNA ZFAS1 | 0 | | | | | | 1 | | Ce: 1 | IS: 241; C: 120 | over: 1 | HC: 1 |
| pan 2024 | ENSG00000196559 | 1 | | B: 1 | IS: 30; C: 30 | over: 1 | HC:1 | 0 | | | | | |
| pan 2024 | ENSG00000202198 | 1 | | B: 1 | IS: 30; C: 30 | over: 1 | HC: 1 | 0 | | | | | |
| pan 2024 | ENSG00000226482 | 1 | | B: 1 | IS: 30; C: 30 | over: 1 | HC: 1 | 0 | | | | | |
| pan 2024 | ENSG00000260539 | 1 | | B: 1 | IS: 30; C: 30 | over: 1 | HC: 1 | 0 | | | | | |
| pan 2024 | ENSG00000269900 | 1 | | B: 1 | IS: 30; C: 30 | over: 1 | HC: 1 | 0 | | | | | |
| pan 2024 | XLOC_013994_2 | 0 | | | | | | 1 | | B: 1 | IS: 30; C: 30 | over: 1 | HC: 1 |
| zhang 2025 | lncRNA H19 | 1 | L: 1 | B: 1 | IS: 36; C: 25 | <4.5 h: 1; <6 h: 1; <9 h: 1; <24 h: 1 | HC: 1 | 0 | | | | | |

**Table S5. Characteristics of the included meta-analyses for Cf DNA biomarkers**

| | | Higher in Stroke | | | | | |
|---|---|---|---|---|---|---|---|
| **Review** | **Biomarker** | **n** | **R** | **M** | **Subjects** | **Time to onset** | **Ctrl** |
| zhang 2025 | Cf DNA (LMW) | 6 | U: 6 | B: 6 | IS: 48; C: 30 | <4.5 h: 1; <6 h: 2; <9 h: 2; <24 h: 4; over: 2 | HC: 6 |
| zhang 2025 | Cf DNA (Nucleic Acid Stain) | 1 | U: 1 | B: 1 | IS: 243; C: 27 | over: 1 | HC: 1 |
| zhang 2025 | Cf DNA B-Globin | 2 | U: 2 | B: 2 | IS: 104; C: 65 | <4.5 h: 1; <6 h: 1; <9 h: 1; <24 h: 1; over: 1 | HC: 2 |
| zhang 2025 | Cf DNA MT-ND2 | 1 | U: 1 | B: 1 | IS: 50; C: 50 | over: 1 | HC: 1 |
| zhang 2025 | Cf DNA TERT | 1 | U: 1 | B: 1 | IS: 43; C: 20 | <24 h: 1 | SM: 1 |

**Table S6. Directions of biomarkers (pooled data from all primary studies with exclusion of duplicates): biochemistry**

| | Higher in Stroke | | | | | | No difference | | | | | | Lower in Stroke | | | | | |
|---|---|---|---|---|---|---|---|---|---|---|---|---|---|---|---|---|---|---|
| **Biomarker** | **n** | **R** | **M** | **Subjects** | **Time to onset** | **Ctrl** | **n** | **R** | **M** | **Subjects** | **Time to onset** | **Ctrl** | **n** | **R** | **M** | **Subjects** | **Time to onset** | **Ctrl** |
| ALP | 1 | L: 1 | B: 1 | IS: 119; C: 904 | over: 1 | OC: 1 | 0 | | | | | | 0 | | | | | |
| ALT | 0 | | | | | | 1 | L: 1 | B: 1 | IS: 119; C: 904 | over: 1 | OC: 1 | 0 | | | | | |
| ANP | 6 | L: 3; U: 3 | B: 6 | IS: 252; C: 199 | over: 6 | HC: 6 | 0 | | | | | | 1 | L: 1 | B: 1 | IS: 52; C: 100 | over: 1 | HC: 1 |
| AST | 0 | | | | | | 1 | L: 1 | B: 1 | IS: 119; C: 904 | over: 1 | OC: 1 | 0 | | | | | |
| Adiponectin | 0 | | | | | | 0 | | | | | | 1 | L: 1 | B: 1 | IS: 68; C: 185 | over: 1 | OC: 1 |
| Albumin | 0 | | | | | | 2 | L: 2 | B: 2 | IS: 88; C: 1235 | <6 h: 1; <9 h: 1; <24 h: 1; over: 1 | OC: 2 | 1 | L: 1 | B: 1 | IS: 119; C: 904 | over: 1 | OC: 1 |
| Ammonia | 0 | | | | | | 1 | L: 1 | B: 1 | IS: 119; C: 904 | over: 1 | OC: 1 | 0 | | | | | |
| BDNF | 10 | L: 5; U: 3; H: 2 | B: 10 | IS: 933; C: 1334 | over: 10 | HC: 10 | 7 | L: 5; U: 2 | B: 6; CSF: 1 | IS: 230; C: 426 | <24 h: 1; over: 6 | HC: 6; OC: 1 | 2 | L: 2 | B: 2 | IS: 156; C: 125 | over: 2 | HC: 2 |
| BNP | 8 | L: 1; U: 6; H: 1 | B: 8 | IS: 1245; C: 393 | <6 h: 1; <9 h: 1; <24 h: 8 | SM: 3; HC: 4; OC: 1 | 2 | U: 2 | B: 2 | IS: 1015; C: 139 | <6 h: 1; <9 h: 1; <24 h: 2 | SM: 2 | 0 | | | | | |
| Blood Urea Nitrogen | 2 | L: 2 | B: 2 | IS: 174; C: 2088 | over: 2 | OC: 2 | 3 | L: 3 | B: 3 | IS: 156; C: 1420 | <6 h: 1; <9 h: 1; <24 h: 1; over: 2 | OC: 3 | 0 | | | | | |
| CRP | 9 | L: 2; U: 1 | B: 3 | IS: 847; C: 2766 | <6 h: 1; <9 h: 1; <24 h: 1; over: 8 | HC: 6; OC: 3 | 1 | | | IS: 11; C: 9 | over: 1 | HC: 1 | 0 | | | | | |
| Calcium | 0 | | | | | | 2 | L: 2 | B: 2 | IS: 150; C: 955 | over: 2 | OC: 2 | 0 | | | | | |
| Calcium 2+ | 0 | | | | | | 1 | L: 1 | B: 1 | IS: 119; C: 904 | over: 1 | OC: 1 | 0 | | | | | |
| Chloride | 0 | | | | | | 3 | L: 3 | B: 3 | IS: 227; C: 3223 | over: 3 | OC: 3 | 0 | | | | | |
| Copeptin | 5 | L: 1; U: 4 | B: 1 | IS: 360; C: 921 | over: 5 | HC: 2; OC: 2 | 1 | U: 1 | | IS: 109; C: 63 | over: 1 | HC: 1 | 0 | | | | | |
| Creatinine | 4 | L: 4 | B: 4 | IS: 230; C: 2174 | <6 h: 1; <9 h: 1; <24 h: 1; over: 3 | OC: 4 | 2 | L: 2 | B: 2 | IS: 86; C: 1235 | over: 2 | OC: 2 | 0 | | | | | |
| Creatinine Kinase | 0 | | | | | | 1 | L: 1 | B: 1 | IS: 35; C: 100 | <6 h: 1; <9 h: 1; <24 h: 1 | OC: 1 | 0 | | | | | |
| Cystatin C | 8 | L: 8 | B: 8 | IS: 3690; C: 4756 | over: 8 | HC: 8 | 0 | | | | | | 1 | L: 1 | B: 1 | IS: 83; C: 71 | over: 1 | HC: 1 |
| Delta Neutrophil Index | 0 | | | | | | 1 | L: 1 | B: 1 | IS: 119; C: 904 | over: 1 | OC: 1 | 0 | | | | | |
| ESR | 0 | | | | | | 1 | U: 1 | B: 1 | IS: 56; C: 161 | over: 1 | OC: 1 | 0 | | | | | |
| Ferritin | 1 | L: 1 | CSF: 1 | IS: 33; C: 20 | over: 1 | HC: 1 | 0 | | | | | | 0 | | | | | |

| | | | | | | | | | | | | | | | | | | |
|---|---|---|---|---|---|---|---|---|---|---|---|---|---|---|---|---|---|---|
| Fibrinogen | 1 | L: 1 | B: 1 | IS: 126; C: 172 | over: 1 | OC: 1 | 0 | | | | | | 0 | | | | | |
| GFAP | 5 | L: 2; U: 3 | B: 5 | IS: 585; C: 378 | <4.5 h: 1; <6 h: 2; <9 h: 2; <24 h: 5 | SM: 1; HC: 4 | 8 | L: 4; U: 4 | B: 8 | IS: 718; C: 372 | <4.5 h: 3; <6 h: 5; <9 h: 6; <24 h: 8 | SM: 4; HC: 3; OC: 1 | 0 | | | | | |
| Glucose | 3 | L: 3 | B: 3 | IS: 131; C: 2354 | over: 3 | OC: 3 | 3 | L: 3 | B: 3 | IS: 185; C: 1055 | <6 h: 1; <9 h: 1; <24 h: 1; over: 2 | OC: 3 | 0 | | | | | |
| HDL | 0 | | | | | | 1 | L: 1 | B: 1 | IS: 68; C: 185 | over: 1 | OC: 1 | 0 | | | | | |
| HIF-1α | 0 | | | | | | 0 | | | | | | 1 | L: 1 | CSF: 1 | IS: 50; C: 30 | over: 1 | HC: 1 |
| HbA1c | 2 | L: 1; U: 1 | B: 2 | IS: 79; C: 196 | over: 2 | OC: 2 | 0 | | | | | | 0 | | | | | |
| Hemoglobin | 1 | L: 1 | B: 1 | IS: 23; C: 35 | over: 1 | OC: 1 | 4 | L: 4 | B: 4 | IS: 258; C: 3274 | over: 4 | OC: 4 | 0 | | | | | |
| ICAM-1 | 8 | L: 3; U: 5 | B: 8 | IS: 726; C: 555 | <24 h: 8 | HC: 5; OC: 2; NC: 1 | 2 | L: 1; U: 1 | B: 2 | IS: 132; C: 115 | <4.5 h: 1; <6 h: 1; <9 h: 1; <24 h: 2 | HC: 1; NC: 1 | 0 | | | | | |
| IL-6 | 14 | L: 12; U: 2 | | IS: 939; C: 685 | <4.5 h: 1; <6 h: 1; <9 h: 1; <24 h: 8; over: 6 | HC: 14 | 2 | L: 1; U: 1 | B: 1 | IS: 64; C: 85 | <24 h: 2 | HC: 1; OC: 1 | 0 | | | | | |
| INR | 0 | | | | | | 2 | L: 2 | B: 2 | IS: 154; C: 1004 | <6 h: 1; <9 h: 1; <24 h: 1; over: 1 | OC: 2 | 0 | | | | | |
| Inorganic Phosphate | 0 | | | | | | 0 | | | | | | 1 | L: 1 | B: 1 | IS: 119; C: 904 | over: 1 | OC: 1 |
| Ischemia-Modified Albumin | 16 | L: 16 | B: 16 | IS: 1045; C: 935 | <24 h: 16 | HC: 16 | 2 | L: 2 | B: 2 | IS: 91; C: 53 | <24 h: 2 | HC: 2 | 0 | | | | | |
| LDH | 0 | | | | | | 1 | L: 1 | B: 1 | IS: 55; C: 1184 | over: 1 | OC: 1 | 0 | | | | | |
| LDL | 1 | L: 1 | B: 1 | IS: 68; C: 185 | over: 1 | OC: 1 | 1 | U: 1 | B: 1 | IS: 56; C: 161 | over: 1 | OC: 1 | 0 | | | | | |
| Leukocytes | 4 | L: 3; U: 1 | B: 4 | IS: 167; C: 1431 | <6 h: 1; <9 h: 1; <24 h: 1; over: 3 | OC: 4 | 3 | L: 3 | B: 3 | IS: 205; C: 2139 | over: 3 | OC: 3 | 0 | | | | | |
| Lymphocytes | 0 | | | | | | 1 | U: 1 | B: 1 | IS: 62; C: 111 | over: 1 | OC: 1 | 1 | L: 1 | B: 1 | IS: 35; C: 100 | <6 h: 1; <9 h: 1; <24 h: 1 | OC: 1 |
| MMP-9 | 12 | L: 3; U: 8; H: 1 | B: 10 | IS: 1904; C: 697 | <6 h: 2; <9 h: 2; <24 h: 12 | SM: 4; HC: 8 | 14 | L: 3; U: 10; H: 1 | B: 14 | IS: 1858; C: 931 | <4.5 h: 1; <6 h: 4; <9 h: 5; <24 h: 14 | SM: 4; HC: 9; OC: 1 | 0 | | | | | |
| Magnesium 2+ | 1 | L: 1 | B: 1 | IS: 119; C: 904 | over: 1 | OC: 1 | 0 | | | | | | 0 | | | | | |
| Myoglobin | 0 | | | | | | 1 | L: 1 | B: 1 | IS: 35; C: 100 | <6 h: 1; <9 h: 1; <24 h: 1 | OC: 1 | 0 | | | | | |
| NGF | 0 | | | | | | 0 | | | | | | 1 | L: 1 | CSF: 1 | IS: 50; C: 30 | over: 1 | HC: 1 |
| NSE | 13 | L: 2; U: 11 | B: 13 | IS: 559; C: 589 | <4.5 h: 2; <6 h: 3; <9 h: 4; <24 h: 12; over: 1 | SM: 1; HC: 9; OC: 3 | 3 | L: 1; U: 2 | B: 3 | IS: 601; C: 225 | <6 h: 1; <9 h: 1; <24 h: 3 | SM: 2; OC: 1 | 0 | | | | | |
| NT-proBNP | 2 | L: 1; U: 1 | B: 2 | IS: 87; C: 87 | <24 h: 2 | HC: 2 | 1 | U: 1 | B: 1 | IS: 941; C: 193 | <6 h: 1; <9 h: 1; <24 h: 1 | SM: 1 | 0 | | | | | |
| Neutrophil Percentage | 1 | U: 1 | B: 1 | IS: 56; C: 161 | over: 1 | OC: 1 | 0 | | | | | | 0 | | | | | |

| | | | | | | | | | | | | | | | | | | |
|---|---|---|---|---|---|---|---|---|---|---|---|---|---|---|---|---|---|---|
| Neutrophil-to-Lymphocyte Ratio | 2 | L: 1; U: 1 | B: 2 | IS: 97; C: 211 | <6 h: 1; <9 h: 1; <24 h: 1; over: 1 | OC: 2 | 0 | | | | | | 0 | | | | | |
| Neutrophils | 2 | L: 1; U: 1 | B: 2 | IS: 97; C: 211 | <6 h: 1; <9 h: 1; <24 h: 1; over: 1 | OC: 2 | 0 | | | | | | 0 | | | | | |
| NfH SMI35 | 0 | | | | | | 1 | L: 1 | CSF: 1 | IS: 33; C: 20 | over: 1 | HC: 1 | 0 | | | | | |
| NfL | 2 | L: 2 | B: 2 | IS: 568; C: 648 | over: 2 | HC: 2 | 1 | L: 1 | B: 1 | IS: 31; C: 29 | <9 h: 1; <24 h: 1 | HC: 1 | 0 | | | | | |
| PT | 0 | | | | | | 1 | L: 1 | B: 1 | IS: 35; C: 100 | <6 h: 1; <9 h: 1; <24 h: 1 | OC: 1 | 0 | | | | | |
| Platelet-to-Lymphocyte Ratio | 0 | | | | | | 1 | U: 1 | B: 1 | IS: 62; C: 111 | over: 1 | OC: 1 | 0 | | | | | |
| Platelets | 1 | L: 1 | B: 1 | IS: 35; C: 100 | <6 h: 1; <9 h: 1; <24 h: 1 | OC: 1 | 5 | L: 4; U: 1 | B: 5 | IS: 288; C: 2236 | over: 5 | OC: 5 | 1 | L: 1 | B: 1 | IS: 55; C: 1184 | over: 1 | OC: 1 |
| Potassium | 0 | | | | | | 3 | L: 3 | B: 3 | IS: 203; C: 2090 | over: 3 | OC: 3 | 1 | L: 1 | B: 1 | IS: 55; C: 1184 | over: 1 | OC: 1 |
| Protein Z | 1 | U: 1 | B: 1 | IS: 173; C: 186 | over: 1 | HC: 1 | 1 | U: 1 | B: 1 | IS: 124; C: 125 | over: 1 | HC: 1 | 1 | U: 1 | B: 1 | IS: 154; C: 206 | over: 1 | HC: 1 |
| S100B | 26 | L: 15; U: 11 | B: 25; CSF: 1 | IS: 1883; C: 1691 | <4.5 h: 1; <6 h: 3; <9 h: 4; <24 h: 14; over: 12 | SM: 5; HC: 13; OC: 7; NC: 1 | 7 | L: 4; U: 3 | B: 7 | IS: 1683; C: 586 | <6 h: 1; <9 h: 1; <24 h: 5; over: 2 | SM: 3; HC: 4 | 0 | | | | | |
| Sodium | 0 | | | | | | 3 | L: 3 | B: 3 | IS: 203; C: 2090 | over: 3 | OC: 3 | 1 | L: 1 | B: 1 | IS: 55; C: 1184 | over: 1 | OC: 1 |
| TNF-α | 6 | L: 3; U: 3 | B: 6 | IS: 410; C: 396 | <24 h: 6 | HC: 2; NS: 1; OC: 1; NC: 2 | 4 | L: 2; U: 2 | B: 4 | IS: 332; C: 233 | <24 h: 4 | SM: 2; HC: 2 | 0 | | | | | |
| Thrombotic Microangiopathy Score | 1 | L: 1 | B: 1 | IS: 119; C: 904 | over: 1 | OC: 1 | 0 | | | | | | 0 | | | | | |
| Total Bilirubin | 0 | | | | | | 1 | L: 1 | B: 1 | IS: 119; C: 904 | over: 1 | OC: 1 | 0 | | | | | |
| Total Cholesterol | 0 | | | | | | 1 | L: 1 | B: 1 | IS: 68; C: 185 | over: 1 | OC: 1 | 0 | | | | | |
| Total Protein | 0 | | | | | | 0 | | | | | | 1 | L: 1 | B: 1 | IS: 119; C: 904 | over: 1 | OC: 1 |
| Triglycerides | 0 | | | | | | 1 | L: 1 | B: 1 | IS: 68; C: 185 | over: 1 | OC: 1 | 0 | | | | | |
| Troponin I | 0 | | | | | | 1 | L: 1 | B: 1 | IS: 35; C: 100 | <6 h: 1; <9 h: 1; <24 h: 1 | OC: 1 | 0 | | | | | |
| Urea | 0 | | | | | | 1 | L: 1 | B: 1 | IS: 31; C: 51 | over: 1 | OC: 1 | 0 | | | | | |
| Uric Acid | 0 | | | | | | 1 | L: 1 | B: 1 | IS: 119; C: 904 | over: 1 | OC: 1 | 0 | | | | | |
| VCAM-1 | 6 | L: 3; U: 3 | B: 6 | IS: 416; C: 434 | <4.5 h: 1; <6 h: 1; <9 h: 1; <24 h: 6 | HC: 4; OC: 1; NC: 1 | 4 | L: 1; U: 3 | B: 4 | IS: 434; C: 233 | <24 h: 4 | HC: 1; OC: 2; NC: 1 | 0 | | | | | |
| VEGF | 0 | | | | | | 10 | L: 1 | B: 9; CSF: 1 | IS: 683; C: 539 | <24 h: 6; over: 4 | HC: 10 | 0 | | | | | |
| aPTT | 1 | L: 1 | B: 1 | IS: 119; C: 904 | over: 1 | OC: 1 | 1 | L: 1 | B: 1 | IS: 35; C: 100 | <6 h: 1; <9 h: 1; <24 h: 1 | OC: 1 | 0 | | | | | |
| d-Dimer | 11 | L: 2; U: 8; H: 1 | B: 11 | IS: 1926; C: 926 | <6 h: 3; <9 h: 3; <24 h: 10; over: 1 | SM: 8; HC: 2; OC: 1 | 2 | U: 2 | B: 2 | IS: 252; C: 95 | <4.5 h: 1; <6 h: 1; <9 h: 1; <24 h: 2 | SM: 2 | 0 | | | | | |

| | | | | | | | | | | | | | |
|---|---|---|---|---|---|---|---|---|---|---|---|---|---|
| e-Selectin | 3 | L: 2; U: 1 | B: 3 | IS: 294; C: 301 | <24 h: 3 | HC: 2; OC: 1 | 2 | U: 2 | B: 2 | IS: 91; C: 67 | <24 h: 2 | HC: 1; OC: 1 | 0 |
| l-Selectin | 1 | U: 1 | B: 1 | IS: 67; C: 76 | <24 h: 1 | HC: 1 | 1 | U: 1 | B: 1 | IS: 22; C: 22 | <4.5 h: 1; <6 h: 1; <9 h: 1; <24 h: 1 | HC: 1 | 0 |
| p-Selectin | 3 | L: 2; U: 1 | B: 3 | IS: 252; C: 249 | <24 h: 3 | HC: 1; OC: 1; NC: 1 | 0 | | | | | | 0 |

**Table S7. Directions of biomarkers (pooled data from all primary studies with exclusion of duplicates): Cf DNA**

| | Higher in Stroke | | | | | |
|---|---|---|---|---|---|---|
| **Biomarker** | **n** | **R** | **M** | **Subjects** | **Time to onset** | **Ctrl** |
| Cf DNA (LMW) | 4 | U: 4 | B: 4 | IS: 32; C: 20 | <4.5 h: 1; <6 h: 2; <9 h: 2; <24 h: 3; over: 1 | HC: 4 |
| Cf DNA (Nucleic Acid Stain) | 1 | U: 1 | B: 1 | IS: 243; C: 27 | over: 1 | HC: 1 |
| Cf DNA B-Globin | 2 | U: 2 | B: 2 | IS: 104; C: 65 | <4.5 h: 1; <6 h: 1; <9 h: 1; <24 h: 1; over: 1 | HC: 2 |
| Cf DNA MT-ND2 | 1 | U: 1 | B: 1 | IS: 50; C: 50 | over: 1 | HC: 1 |
| Cf DNA TERT | 1 | U: 1 | B: 1 | IS: 43; C: 20 | <24 h: 1 | SM: 1 |

**Table S8. Directions of biomarkers (pooled data from all primary studies with exclusion of duplicates): CircRNA**

| | Higher in Stroke | | | | | | Lower in Stroke | | | | | |
|---|---|---|---|---|---|---|---|---|---|---|---|---|
| **Biomarker** | **n** | **R** | **M** | **Subjects** | **Time to onset** | **Ctrl** | **n** | **R** | **M** | **Subjects** | **Time to onset** | **Ctrl** |
| Circ_0000097 | 1 | H: 1 | B: 1 | IS: 200; C: 100 | over: 1 | HC: 1 | 0 | | | | | |
| Circ_0000607 | 0 | | | | | | 1 | U: 1 | B: 1 | IS: 32; C: 32 | over: 1 | HC: 1 |
| Circ_0000698 | 0 | | | | | | 1 | H: 1 | B: 1 | IS: 32; C: 32 | over: 1 | HC: 1 |
| Circ_0001460 | 2 | U: 1; H: 1 | Ce: 2 | IS: 286; C: 236 | <4.5 h: 1; <6 h: 1; <9 h: 1; <24 h: 2 | HC: 2 | 0 | | | | | |
| Circ_0001599 | 1 | H: 1 | Ce: 1 | IS: 168; C: 118 | <4.5 h: 1; <6 h: 1; <9 h: 1; <24 h: 1 | HC: 1 | 1 | U: 1 | Ce: 1 | IS: 118; C: 118 | <24 h: 1 | HC: 1 |
| Circ_0002465 | 0 | | | | | | 1 | U: 1 | B: 1 | IS: 32; C: 32 | over: 1 | HC: 1 |
| Circ_0004338 | 1 | H: 1 | Ce: 1 | IS: 168; C: 118 | <4.5 h: 1; <6 h: 1; <9 h: 1; <24 h: 1 | HC: 1 | 1 | U: 1 | Ce: 1 | IS: 118; C: 118 | <24 h: 1 | HC: 1 |
| Circ_0004494 | 1 | H: 1 | B: 1 | IS: 200; C: 100 | over: 1 | HC: 1 | 0 | | | | | |
| Circ_0005548 | 1 | U: 1 | B: 1 | IS: 32; C: 32 | over: 1 | HC: 1 | 0 | | | | | |
| Circ_0005585 | 0 | | | | | | 1 | H: 1 | B: 1 | IS: 32; C: 32 | over: 1 | HC: 1 |
| Circ_0006911 | 0 | | | | | | 1 | H: 1 | Ce: 1 | IS: 168; C: 118 | <4.5 h: 1; <6 h: 1; <9 h: 1; <24 h: 1 | HC: 1 |
| Circ_0007290 | 1 | H: 1 | B: 1 | IS: 200; C: 100 | over: 1 | HC: 1 | 0 | | | | | |
| Circ_0007637 | 0 | | | | | | 2 | U: 1; H: 1 | Ce: 2 | IS: 286; C: 236 | <4.5 h: 1; <6 h: 1; <9 h: 1; <24 h: 2 | HC: 2 |
| Circ_0010155 | 0 | | | | | | 1 | H: 1 | B: 1 | IS: 32; C: 32 | over: 1 | HC: 1 |
| Circ_0039457 | 0 | | | | | | 1 | | B: 1 | IS: 8; C: 8 | <4.5 h: 1; <6 h: 1; <9 h: 1; <24 h: 1 | HC: 1 |
| Circ_0043837 | 0 | | | | | | 1 | H: 1 | B: 1 | IS: 32; C: 32 | over: 1 | HC: 1 |
| Circ_0072309 | 0 | | | | | | 1 | H: 1 | B: 1 | IS: 90; C: 75 | <24 h: 1 | HC: 1 |
| Circ_0073239 | 0 | | | | | | 1 | U: 1 | Ce: 1 | IS: 118; C: 118 | <24 h: 1 | HC: 1 |
| Circ_0090002 | 0 | | | | | | 1 | | B: 1 | IS: 8; C: 8 | <4.5 h: 1; <6 h: 1; <9 h: 1; <24 h: 1 | HC: 1 |
| Circ_0141720 | 1 | L: 1 | B: 1 | IS: 80; C: 30 | over: 1 | HC: 1 | 0 | | | | | |
| circOGDH | 1 | U: 1 | B: 1 | IS: 45; C: 32 | <24 h: 1 | HC: 1 | 0 | | | | | |
| circRNA CCZ1 | 1 | H: 1 | B: 1 | IS: 145; C: 145 | over: 1 | HC: 1 | 0 | | | | | |
| circRNA CDC14A | 1 | H: 1 | B: 1 | IS: 145; C: 145 | over: 1 | HC: 1 | 0 | | | | | |
| circRNA CDC42BPA | 1 | H: 1 | B: 1 | IS: 145; C: 145 | over: 1 | HC: 1 | 0 | | | | | |
| circRNA CEP78 | 0 | | | | | | 1 | | Ce: 1 | IS: 5; C: 5 | over: 1 | HC: 1 |
| circRNA DAB1 | 0 | | | | | | 1 | | Ce: 1 | IS: 5; C: 5 | over: 1 | HC: 1 |
| circRNA DLGAP4 | 0 | | | | | | 1 | U: 1 | Ce: 1 | IS: 170; C: 170 | <24 h: 1 | HC: 1 |
| circRNA FBXW4 | 0 | | | | | | 1 | H: 1 | B: 1 | IS: 145; C: 145 | over: 1 | HC: 1 |
| circRNA GNB2L1 | 0 | | | | | | 1 | H: 1 | B: 1 | IS: 145; C: 145 | over: 1 | HC: 1 |

| | | | | | | | | | | | | |
|---|---|---|---|---|---|---|---|---|---|---|---|---|
| circRNA HECTD1 | 2 | U: 1 | B: 1; Ce: 1 | IS: 197; C: 194 | <24 h: 2 | HC: 2 | 0 | | | | | |
| circRNA KLRK1 | 0 | | | | | | 1 | | Ce: 1 | IS: 5; C: 5 | over: 1 | HC: 1 |
| circRNA PDE4B | 0 | | | | | | 1 | H: 1 | B: 1 | IS: 145; C: 145 | over: 1 | HC: 1 |
| circRNA PIGB | 1 | | Ce: 1 | IS: 5; C: 5 | over: 1 | HC: 1 | 0 | | | | | |
| circRNA PLXNC1 | 1 | | Ce: 1 | IS: 5; C: 5 | over: 1 | HC: 1 | 0 | | | | | |
| circRNA RAP1B | 0 | | | | | | 1 | | Ce: 1 | IS: 5; C: 5 | over: 1 | HC: 1 |
| circRNA RBM33 | 1 | H: 1 | B: 1 | IS: 145; C: 145 | over: 1 | HC: 1 | 0 | | | | | |
| circRNA SCMH1 | 0 | | | | | | 1 | H: 1 | B: 1 | IS: 145; C: 145 | over: 1 | HC: 1 |
| circRNA TLK1 | 1 | U: 1 | B: 1 | IS: 71; C: 71 | <24 h: 1 | HC: 1 | 0 | | | | | |
| circRNA TMEM56 | 1 | | Ce: 1 | IS: 5; C: 5 | over: 1 | HC: 1 | 0 | | | | | |
| circRNA TREM1 | 1 | | Ce: 1 | IS: 5; C: 5 | over: 1 | HC: 1 | 0 | | | | | |
| circRNA ZCCHC11 | 1 | H: 1 | B: 1 | IS: 145; C: 145 | over: 1 | HC: 1 | 0 | | | | | |

**Table S9. Directions of biomarkers (pooled data from all primary studies with exclusion of duplicates): gene/transcript**

| | Higher in Stroke | | | | | | Lower in Stroke | | | | | |
|---|---|---|---|---|---|---|---|---|---|---|---|---|
| **Biomarker** | **n** | **R** | **M** | **Subjects** | **Time to onset** | **Ctrl** | **n** | **R** | **M** | **Subjects** | **Time to onset** | **Ctrl** |
| EGR2 gene | 1 | U: 1 | Ce: 1 | IS: 8; C: 4 | <24 h: 1 | HC: 1 | 0 | | | | | |
| ENSG00000196559 | 1 | | B: 1 | IS: 30; C: 30 | over: 1 | HC: 1 | 0 | | | | | |
| ENSG00000202198 | 1 | | B: 1 | IS: 30; C: 30 | over: 1 | HC: 1 | 0 | | | | | |
| ENSG00000226482 | 1 | | B: 1 | IS: 30; C: 30 | over: 1 | HC: 1 | 0 | | | | | |
| ENSG00000260539 | 1 | | B: 1 | IS: 30; C: 30 | over: 1 | HC: 1 | 0 | | | | | |
| ENSG00000269900 | 1 | | B: 1 | IS: 30; C: 30 | over: 1 | HC: 1 | 0 | | | | | |
| XLOC_013994_2 | 0 | | | | | | 1 | | B: 1 | IS: 30; C: 30 | over: 1 | HC: 1 |

**Table S10. Directions of biomarkers (pooled data from all primary studies with exclusion of duplicates): lncRNA**

| | Higher in Stroke | | | | | | Lower in Stroke | | | | | |
|---|---|---|---|---|---|---|---|---|---|---|---|---|
| Biomarker | n | R | M | Subjects | Time to onset | Ctrl | n | R | M | Subjects | Time to onset | Ctrl |
| ADAMTS9-AS2 | 0 | | | | | | 1 | | B: 1 | IS: 43; C: 68 | over: 1 | HC: 1 |
| NBAT1 | 1 | H: 1 | B: 1 | IS: 60; C: 60 | over: 1 | HC: 1 | 0 | | | | | |
| SNHG15 | 1 | | B: 1 | IS: 30; C: 30 | over: 1 | HC: 1 | 0 | | | | | |
| SNHG5 | 1 | | B: 1 | IS: 10; C: 10 | over: 1 | HC: 1 | 0 | | | | | |
| TUG1 | 1 | H: 1 | B: 1 | IS: 60; C: 60 | over: 1 | HC: 1 | 0 | | | | | |
| lncRNA AC136007.2 | 0 | | | | | | 1 | H: 1 | B: 1 | IS: 32; C: 32 | over: 1 | HC: 1 |
| lncRNA AK139328 | 1 | | B: 1 | IS: 30; C: 30 | over: 1 | HC: 1 | 0 | | | | | |
| lncRNA ANRIL | 3 | H: 2 | B: 3 | IS: 297; C: 278 | over: 3 | HC: 3 | 1 | U: 1 | B: 1 | IS: 126; C: 125 | over: 1 | HC: 1 |
| lncRNA C14orf64 | 0 | | | | | | 1 | H: 1 | B: 1 | IS: 32; C: 32 | over: 1 | HC: 1 |
| lncRNA CASC15 | 1 | H: 1 | B: 1 | IS: 98; C: 85 | over: 1 | HC: 1 | 0 | | | | | |
| lncRNA DHFRL1-4 | 1 | | Ce: 1 | IS: 32; C: 32 | over: 1 | HC: 1 | 0 | | | | | |
| lncRNA ENST00000530525 | 0 | | | | | | 1 | | B: 1 | IS: 5; C: 5 | over: 1 | HC: 1 |
| lncRNA ENST00000568297 | 1 | H: 1 | B: 1 | IS: 50; C: 50 | over: 1 | HC: 1 | 0 | | | | | |
| lncRNA FAM98A-3 | 1 | | B: 1 | IS: 30; C: 30 | over: 1 | HC: 1 | 0 | | | | | |
| lncRNA GAS5 | 3 | L: 1; U: 1 | B: 2; Ce: 1 | IS: 268; C: 205 | over: 3 | HC: 3 | 0 | | | | | |
| lncRNA H19 | 7 | L: 1; U: 2; H: 1 | B: 7 | IS: 431; C: 339 | <4.5 h: 1; <6 h: 1; <9 h: 1; <24 h: 1; over: 6 | HC: 7 | 0 | | | | | |
| lncRNA HULC | 1 | U: 1 | B: 1 | IS: 215; C: 215 | over: 1 | HC: 1 | 0 | | | | | |
| lncRNA ITSN1-2 | 2 | | B: 1; Ce: 1 | IS: 422; C: 370 | over: 2 | HC: 2 | 0 | | | | | |
| lncRNA KCNQ1OT1 | 1 | | B: 1 | IS: 42; C: 40 | over: 1 | HC: 1 | 0 | | | | | |
| lncRNA MALAT1 | 0 | | | | | | 2 | H: 1 | B: 2 | IS: 220; C: 220 | over: 2 | HC: 2 |
| lncRNA MEG3 | 1 | | Ce: 1 | IS: 170; C: 100 | over: 1 | HC: 1 | 0 | | | | | |
| lncRNA MIAT | 1 | U: 1 | B: 1 | IS: 232; C: 232 | over: 1 | HC: 1 | 1 | U: 1 | B: 1 | IS: 80; C: 40 | over: 1 | HC: 1 |
| lncRNA NEAT1 | 1 | U: 1 | B: 1 | IS: 210; C: 210 | over: 1 | HC: 1 | 0 | | | | | |
| lncRNA NORAD | 1 | H: 1 | B: 1 | IS: 103; C: 95 | over: 1 | HC: 1 | 0 | | | | | |
| lncRNA NR_046084 | 1 | H: 1 | B: 1 | IS: 50; C: 50 | over: 1 | HC: 1 | 0 | | | | | |
| lncRNA OIP5-AS1 | 0 | | | | | | 1 | | B: 1 | IS: 15; C: 15 | over: 1 | HC: 1 |
| lncRNA PVT1 | 2 | H: 1 | B: 2 | IS: 140; C: 140 | over: 2 | HC: 2 | 0 | | | | | |
| lncRNA SERPINB9P1 | 0 | | | | | | 1 | | B: 1 | IS: 159; C: 153 | over: 1 | HC: 1 |
| lncRNA SNHG16 | 0 | | | | | | 1 | | Ce: 1 | IS: 120; C: 60 | over: 1 | HC: 1 |

| | | | | | | | | | | |
|---|---|---|---|---|---|---|---|---|---|---|
| lncRNA TUG1 | 1 | B: 1 | IS: 46; C: 46 | over: 1 | HC: 1 | 0 | | | | |
| lncRNA UCA1 | 1 | Ce: 1 | IS: 160; C: 160 | over: 1 | HC: 1 | 0 | | | | |
| lncRNA WT1-as | 0 | | | | | 1 | B: 1 | IS: 30; C: 30 | over: 1 | HC: 1 |
| lncRNA XIST | 0 | | | | | 1 | B: 1 | IS: 77; C: 60 | over: 1 | HC: 1 |
| lncRNA ZFAS1 | 0 | | | | | 1 | Ce: 1 | IS: 241; C: 120 | over: 1 | HC: 1 |

**Table S11. Directions of biomarkers (pooled data from all primary studies with exclusion of duplicates): snRNA**

| | Higher in Stroke | | | | | | Lower in Stroke | | | | | |
|---|---|---|---|---|---|---|---|---|---|---|---|---|
| **Biomarker** | **n** | **R** | **M** | **Subjects** | **Time to onset** | **Ctrl** | **n** | **R** | **M** | **Subjects** | **Time to onset** | **Ctrl** |
| RNU48-A2 | 1 | | B: 1 | IS: 55; C: 2360 | <24 h: 1 | NS: 1 | 0 | | | | | |
| RNU48-B1 | 1 | | B: 1 | IS: 55; C: 2360 | <24 h: 1 | NS: 1 | 0 | | | | | |
| RNU48-B2 | 1 | | B: 1 | IS: 55; C: 2360 | <24 h: 1 | NS: 1 | 0 | | | | | |
| U6-snRNA-A1 | 0 | | | | | | 1 | | B: 1 | IS: 55; C: 2360 | <24 h: 1 | NS: 1 |

### Table S12. Directions of biomarkers (pooled data from all primary studies with exclusion of duplicates): miRNA

| | Higher in Stroke | | | | | | Mixed or no difference | | | | | | Lower in Stroke | | | | | |
|---|---|---|---|---|---|---|---|---|---|---|---|---|---|---|---|---|---|---|
| Biomarker | n | R | M | Subjects | Time to onset | Ctrl | n | R | M | Subjects | Time to onset | Ctrl | n | R | M | Subjects | Time to onset | Ctrl |
| Let-7b | 1 | | B: 1 | IS: 87; C: 13 | <24 h: 1 | NS: 1 | 1 | H: 1 | B: 1 | IS: 38; C: 50 | <24 h: 1 | HC: 1 | 0 | | | | | |
| Let-7c | 2 | L: 1 | CSF: 2 | IS: 20; C: 20 | <24 h: 1; over: 1 | HC: 1; NS: 1 | 0 | | | | | | 0 | | | | | |
| Let-7d-3p | 0 | | | | | | 0 | | | | | | 2 | U: 1 | B: 2 | IS: 249; C: 2384 | <24 h: 2 | HC: 1; NS: 1 |
| Let-7e | 4 | L: 1; U: 2 | B: 3; CSF: 1 | IS: 198; C: 86 | <24 h: 3; over: 1 | HC: 3; NS: 1 | 0 | | | | | | 0 | | | | | |
| Let-7e-5p | 2 | H: 1 | B: 2 | IS: 356; C: 357 | <24 h: 2 | HC: 2 | 0 | | | | | | 0 | | | | | |
| Let-7f-5p | 0 | | | | | | 0 | | | | | | 1 | | B: 1 | IS: 28; C: 35 | <24 h: 1 | HC: 1 |
| Let-7i-5p | 0 | | | | | | 0 | | | | | | 2 | | B: 1; Ce: 1 | IS: 70; C: 63 | <24 h: 2 | HC: 1; NS: 1 |
| miR-101 | 0 | | | | | | 0 | | | | | | 2 | H: 1 | B: 2 | IS: 212; C: 220 | <24 h: 2 | HC: 1; NS: 1 |
| miR-106b-5p | 3 | U: 2 | B: 3 | IS: 286; C: 221 | <24 h: 3 | HC: 2; NS: 1 | 0 | | | | | | 0 | | | | | |
| miR-107 | 3 | | B: 2; CSF: 1 | IS: 168; C: 102 | <24 h: 3 | NS: 3 | 0 | | | | | | 0 | | | | | |
| miR-122-5p | 1 | | B: 1 | IS: 34; C: 11 | <24 h: 1 | HC: 1 | 0 | | | | | | 1 | | Ce: 1 | IS: 24; C: 24 | <24 h: 1 | HC: 1 |
| miR-1229-3p | 1 | | B: 1 | IS: 10; C: 11 | <24 h: 1 | HC: 1 | 0 | | | | | | 0 | | | | | |
| miR-1238-5p | 1 | | B: 1 | IS: 10; C: 11 | <24 h: 1 | HC: 1 | 0 | | | | | | 0 | | | | | |
| miR-124 | 0 | | | | | | 0 | | | | | | 3 | L: 1; U: 1 | B: 3 | IS: 193; C: 170 | <24 h: 3 | HC: 2; NS: 1 |
| miR-124-3p | 2 | | B: 1; CSF: 1 | IS: 33; C: 21 | <24 h: 2 | NS: 2 | 0 | | | | | | 4 | | B: 4 | IS: 96; C: 21 | <24 h: 4 | HC: 2; NS: 2 |
| miR-1246 | 3 | U: 1 | B: 2; CSF: 1 | IS: 255; C: 185 | <24 h: 3 | HC: 2; NS: 1 | 0 | | | | | | 0 | | | | | |
| miR-1255b-5p | 0 | | | | | | 0 | | | | | | 1 | U: 1 | B: 1 | IS: 22; C: 22 | <24 h: 1 | HC: 1 |
| miR-125a | 1 | | B: 1 | IS: 87; C: 13 | <24 h: 1 | NS: 1 | 0 | | | | | | 0 | | | | | |
| miR-125a-5p | 2 | L: 1; U: 1 | B: 2 | IS: 283; C: 195 | <24 h: 1; over: 1 | HC: 1; OC: 1 | 0 | | | | | | 0 | | | | | |
| miR-125b | 1 | | B: 1 | IS: 87; C: 13 | <24 h: 1 | NS: 1 | 0 | | | | | | 0 | | | | | |
| miR-125b-2 | 1 | H: 1 | B: 1 | IS: 45; C: 24 | <24 h: 1 | HC: 1 | 0 | | | | | | 0 | | | | | |
| miR-125b-2-3p | 1 | U: 1 | B: 1 | IS: 169; C: 24 | <24 h: 1 | HC: 1 | 0 | | | | | | 0 | | | | | |
| miR-125b-5p | 2 | L: 1; U: 1 | B: 2 | IS: 283; C: 195 | <24 h: 1; over: 1 | HC: 1; OC: 1 | 0 | | | | | | 1 | | B: 1 | IS: 55; C: 0 | <24 h: 1 | NS: 1 |
| miR-126 | 0 | | | | | | 0 | | | | | | 5 | H: 3 | B: 5 | IS: 546; C: 566 | <24 h: 5 | HC: 3; NS: 2 |
| miR-126-3p | 0 | | | | | | 0 | | | | | | 5 | | B: 5 | IS: 324; C: 330 | <24 h: 5 | HC: 5 |
| miR-1261 | 1 | U: 1 | B: 1 | IS: 169; C: 24 | <24 h: 1 | HC: 1 | 0 | | | | | | 0 | | | | | |
| miR-1264 | 1 | | B: 1 | | <24 h: 1 | NS: 1 | 0 | | | | | | 0 | | | | | |
| miR-1270 | 1 | | B: 1 | IS: 10; C: 11 | <24 h: 1 | HC: 1 | 0 | | | | | | 0 | | | | | |
| miR-1271-5p-B1 | 0 | | | | | | 0 | | | | | | 1 | | B: 1 | IS: 80; C: 2360 | <24 h: 1 | NS: 1 |

| | | | | | | | | | | | | | |
|---|---|---|---|---|---|---|---|---|---|---|---|---|---|
| miR-1273g-3p | 0 | | | | | | 0 | 1 | | B: 1 | IS: 17; C: 25 | <24 h: 1 | NS: 1 |
| miR-1275 | 1 | | B: 1 | IS: 279; C: 279 | <24 h: 1 | HC: 1 | 0 | 0 | | | | | |
| miR-1279 | 0 | | | | | | 0 | 1 | | B: 1 | | <24 h: 1 | NS: 1 |
| miR-128-3p | 3 | | B: 2; CSF: 1 | IS: 101; C: 81 | <24 h: 3 | HC: 1; NS: 2 | 0 | 0 | | | | | |
| miR-128b | 1 | | B: 1 | IS: 114; C: 58 | <24 h: 1 | NS: 1 | 0 | 0 | | | | | |
| miR-129-1-3p | 0 | | | | | | 0 | 1 | | B: 1 | IS: 80; C: 2360 | <24 h: 1 | NS: 1 |
| miR-129-2-3p | 0 | | | | | | 0 | 1 | | B: 1 | IS: 270; C: 270 | <24 h: 1 | NS: 1 |
| miR-1294 | 1 | | B: 1 | IS: 10; C: 11 | <24 h: 1 | HC: 1 | 0 | 0 | | | | | |
| miR-1299 | 1 | | B: 1 | IS: 117; C: 82 | <24 h: 1 | HC: 1 | 0 | 1 | U: 1 | B: 1 | IS: 169; C: 24 | <24 h: 1 | HC: 1 |
| miR-1301-3p | 1 | | B: 1 | IS: 10; C: 11 | <24 h: 1 | HC: 1 | 0 | 0 | | | | | |
| miR-130a | 0 | | | | | | 0 | 5 | U: 1; H: 2 | B: 5 | IS: 677; C: 540 | <24 h: 5 | HC: 3; NS: 2 |
| miR-130a-3p | 2 | | B: 1; Ce: 1 | IS: 52; C: 43 | <24 h: 2 | HC: 1; NS: 1 | 0 | 2 | | B: 2 | IS: 254; C: 258 | <24 h: 2 | HC: 2 |
| miR-130b-3p | 1 | | B: 1 | IS: 33; C: 23 | <24 h: 1 | NS: 1 | 0 | 0 | | | | | |
| miR-1321 | 1 | U: 1 | B: 1 | IS: 169; C: 24 | <24 h: 1 | HC: 1 | 0 | 0 | | | | | |
| miR-134 | 1 | H: 1 | B: 1 | IS: 50; C: 50 | <24 h: 1 | HC: 1 | 0 | 0 | | | | | |
| miR-140-3p | 1 | | B: 1 | IS: 33; C: 23 | <24 h: 1 | NS: 1 | 0 | 0 | | | | | |
| miR-140-5p | 1 | | B: 1 | IS: 10; C: 10 | <24 h: 1 | NS: 1 | 0 | 1 | | B: 1 | IS: 10; C: 11 | <24 h: 1 | HC: 1 |
| miR-142-3p | 0 | | | | | | 0 | 2 | | B: 2 | IS: 57; C: 107 | <24 h: 2 | HC: 2 |
| miR-143-3p | 2 | L: 1; U: 1 | B: 2 | IS: 283; C: 195 | <24 h: 1; over: 1 | HC: 1; OC: 1 | 0 | 0 | | | | | |
| miR-144-3p | 1 | | B: 1 | IS: 19; C: 5 | <24 h: 1 | HC: 1 | 0 | 1 | | B: 1 | IS: 10; C: 11 | <24 h: 1 | HC: 1 |
| miR-145 | 2 | H: 1 | B: 2 | IS: 176; C: 126 | <24 h: 2 | HC: 1; NS: 1 | 0 | 0 | | | | | |
| miR-145-5p | 1 | | B: 1 | IS: 146; C: 96 | <24 h: 1 | HC: 1 | 0 | 0 | | | | | |
| miR-146a-5p | 0 | | | | | | 0 | 3 | | B: 2; E: 1 | IS: 104; C: 52 | <24 h: 3 | HC: 2; NS: 1 |
| miR-146b | 1 | H: 1 | B: 1 | IS: 128; C: 102 | <24 h: 1 | HC: 1 | 0 | 0 | | | | | |
| miR-146b-5p | 1 | | B: 1 | IS: 128; C: 102 | <24 h: 1 | HC: 1 | 0 | 0 | | | | | |
| miR-148a-3p | 0 | | | | | | 0 | 1 | | Ce: 1 | IS: 24; C: 24 | <24 h: 1 | HC: 1 |
| miR-148b-3p | 0 | | | | | | 0 | 1 | | B: 1 | IS: 77; C: 42 | <24 h: 1 | NS: 1 |
| miR-149-5p | 0 | | | | | | 0 | 1 | | B: 1 | IS: 47; C: 96 | <24 h: 1 | HC: 1 |
| miR-151a-3p | 2 | | B: 2 | IS: 43; C: 33 | <24 h: 2 | NS: 2 | 0 | 0 | | | | | |
| miR-151b | 1 | | B: 1 | IS: 77; C: 42 | <24 h: 1 | NS: 1 | 0 | 0 | | | | | |
| miR-153 | 1 | | B: 1 | IS: 114; C: 58 | <24 h: 1 | NS: 1 | 0 | 0 | | | | | |
| miR-155 | 2 | U: 1 | B: 2 | IS: 46; C: 50 | <24 h: 2 | HC: 1; NS: 1 | 0 | 0 | | | | | |
| miR-15a | 1 | | B: 1 | IS: 106; C: 120 | <24 h: 1 | NS: 1 | 0 | 1 | | B: 1 | IS: 46; C: 39 | <24 h: 1 | NS: 1 |

| | | | | | | | | | | | | | |
|---|---|---|---|---|---|---|---|---|---|---|---|---|---|
| miR-15a-5p | 1 | | B: 1 | IS: 106; C: 120 | <24 h: 1 | HC: 1 | 0 | 0 | | | | | |
| miR-16 | 4 | U: 2 | B: 4 | IS: 292; C: 173 | <6 h: 1; <9 h: 1; <24 h: 4 | HC: 2; NS: 2 | 0 | 0 | | | | | |
| miR-16-1-3p | 1 | | B: 1 | IS: 80; C: 2360 | <24 h: 1 | NS: 1 | 0 | 0 | | | | | |
| miR-16-5p | 3 | | B: 3 | IS: 158; C: 148 | <24 h: 3 | HC: 2; NS: 1 | 0 | 0 | | | | | |
| miR-17-5p | 2 | | B: 2 | IS: 245; C: 154 | <24 h: 2 | HC: 1; NS: 1 | 0 | 0 | | | | | |
| miR-17-92 | 0 | | | | | | 0 | 1 | | E: 1 | | <24 h: 1 | NS: 1 |
| miR-181a-3p | 1 | | B: 1 | IS: 10; C: 10 | <24 h: 1 | NS: 1 | 0 | 0 | | | | | |
| miR-181a-5p | 1 | | B: 1 | IS: 227; C: 92 | <24 h: 1 | NS: 1 | 0 | 0 | | | | | |
| miR-181b-3p | 1 | | B: 1 | IS: 5; C: 5 | <24 h: 1 | NS: 1 | 0 | 0 | | | | | |
| miR-181d-5p | 1 | | B: 1 | IS: 5; C: 5 | <24 h: 1 | NS: 1 | 0 | 0 | | | | | |
| miR-185 | 4 | H: 2 | B: 4 | IS: 508; C: 516 | <24 h: 4 | HC: 2; NS: 2 | 0 | 0 | | | | | |
| miR-185-5p | 3 | | B: 3 | IS: 396; C: 308 | <24 h: 3 | HC: 3 | 0 | 1 | | B: 1 | IS: 60; C: 30 | <24 h: 1 | HC: 1 |
| miR-186-5p | 0 | | | | | | 0 | 1 | | B: 1 | IS: 10; C: 11 | <24 h: 1 | HC: 1 |
| miR-18b-5p | 0 | | | | | | 0 | 2 | | B: 2 | IS: 20; C: 21 | <24 h: 2 | HC: 1; NS: 1 |
| miR-1908 | 0 | | | | | | 0 | 1 | | B: 1 | IS: 87; C: 13 | <24 h: 1 | NS: 1 |
| miR-1913 | 0 | | | | | | 0 | 1 | | B: 1 | IS: 117; C: 82 | <24 h: 1 | HC: 1 |
| miR-192-5p | 0 | | | | | | 0 | 1 | | B: 1 | IS: 55; C: 0 | <24 h: 1 | NS: 1 |
| miR-193b-3p | 0 | | | | | | 0 | 1 | | B: 1 | IS: 80; C: 2360 | <24 h: 1 | NS: 1 |
| miR-195-5p | 1 | | B: 1 | IS: 18; C: 20 | <24 h: 1 | HC: 1 | 0 | 0 | | | | | |
| miR-19a | 0 | | | | | | 0 | 1 | | B: 1 | IS: 106; C: 110 | <24 h: 1 | NS: 1 |
| miR-19a-3p | 1 | | B: 1 | IS: 28; C: 35 | <24 h: 1 | HC: 1 | 0 | 2 | | B: 1; Ce: 1 | IS: 34; C: 35 | <24 h: 2 | HC: 2 |
| miR-206 | 1 | H: 1 | B: 1 | IS: 106; C: 110 | <24 h: 1 | HC: 1 | 0 | 0 | | | | | |
| miR-208a | 0 | | | | | | 0 | 1 | U: 1 | B: 1 | IS: 169; C: 24 | <24 h: 1 | HC: 1 |
| miR-20b-5p | 1 | | B: 1 | IS: 139; C: 34 | <24 h: 1 | HC: 1 | 0 | 0 | | | | | |
| miR-21 | 1 | | B: 1 | IS: 30; C: 30 | <24 h: 1 | NS: 1 | 0 | 1 | U: 1 | B: 1 | IS: 68; C: 21 | <24 h: 1 | HC: 1 |
| miR-21-5p | 2 | | B: 2 | IS: 246; C: 97 | <24 h: 2 | HC: 1; NS: 1 | 0 | 0 | | | | | |
| miR-210-3p | 0 | | | | | | 0 | 1 | | B: 1 | IS: 112; C: 60 | <24 h: 1 | HC: 1 |
| miR-211-5p | 1 | | B: 1 | IS: 34; C: 11 | <24 h: 1 | HC: 1 | 0 | 0 | | | | | |
| miR-218 | 4 | H: 2 | B: 4 | IS: 508; C: 516 | <24 h: 4 | HC: 2; NS: 2 | 0 | 0 | | | | | |
| miR-218-5p | 1 | | B: 1 | IS: 106; C: 110 | <24 h: 1 | HC: 1 | 0 | 1 | | B: 1 | IS: 10; C: 10 | <24 h: 1 | HC: 1 |
| miR-219 | 0 | | | | | | 0 | 1 | L: 1 | B: 1 | IS: 31; C: 11 | <24 h: 1 | HC: 1 |
| miR-219-5p | 1 | | B: 1 | IS: 148; C: 148 | <24 h: 1 | HC: 1 | 0 | 0 | | | | | |
| miR-22-3p | 1 | | B: 1 | IS: 10; C: 10 | <24 h: 1 | HC: 1 | 0 | 1 | | B: 1 | IS: 34; C: 11 | <24 h: 1 | HC: 1 |

| | | | | | | | | | | | | | |
|---|---|---|---|---|---|---|---|---|---|---|---|---|---|
| miR-22-5p | 0 | | | | | | 0 | 1 | U: 1 | B: 1 | IS: 169; C: 24 | <24 h: 1 | HC: 1 |
| miR-221 | 0 | | | | | | 0 | 2 | H: 1 | B: 2 | IS: 176; C: 126 | <24 h: 2 | HC: 1; NS: 1 |
| miR-221-3p | 2 | L: 1 | CSF: 2 | IS: 20; C: 20 | <24 h: 1; over: 1 | HC: 1; NS: 1 | 0 | 2 | U: 1 | B: 2 | IS: 224; C: 135 | <24 h: 2 | HC: 2 |
| miR-222 | 4 | H: 2 | B: 4 | IS: 508; C: 516 | <24 h: 4 | HC: 2; NS: 2 | 0 | 0 | | | | | |
| miR-222-3p | 2 | | B: 2 | IS: 254; C: 258 | <24 h: 2 | HC: 2 | 0 | 0 | | | | | |
| miR-223 | 1 | | E: 1 | IS: 50; C: 33 | <24 h: 1 | NS: 1 | 0 | 0 | | | | | |
| miR-223-3p | 2 | | B: 2 | IS: 52; C: 28 | <24 h: 2 | HC: 1; NS: 1 | 0 | 0 | | | | | |
| miR-224-3p | 0 | | | | | | 0 | 1 | | B: 1 | IS: 117; C: 82 | <24 h: 1 | HC: 1 |
| miR-23a | 0 | | | | | | 0 | 2 | H: 1 | B: 2 | IS: 176; C: 126 | <24 h: 2 | HC: 1; NS: 1 |
| miR-23a-3p | 1 | | B: 1 | IS: 10; C: 10 | <24 h: 1 | HC: 1 | 0 | 2 | | B: 2 | IS: 180; C: 107 | <24 h: 2 | HC: 2 |
| miR-23b-3p | 1 | | B: 1 | IS: 227; C: 92 | <24 h: 1 | NS: 1 | 0 | 0 | | | | | |
| miR-24 | 0 | | | | | | 0 | 1 | U: 1 | B: 1 | IS: 68; C: 21 | <24 h: 1 | HC: 1 |
| miR-25-3p | 1 | | B: 1 | IS: 33; C: 23 | <24 h: 1 | NS: 1 | 0 | 0 | | | | | |
| miR-26b-5p | 1 | | B: 1 | IS: 80; C: 2360 | <24 h: 1 | NS: 1 | 0 | 0 | | | | | |
| miR-27a | 1 | H: 1 | B: 1 | IS: 45; C: 24 | <24 h: 1 | HC: 1 | 0 | 0 | | | | | |
| miR-27a-5p | 1 | U: 1 | B: 1 | IS: 169; C: 24 | <24 h: 1 | HC: 1 | 0 | 0 | | | | | |
| miR-27b-3p | 2 | | B: 2 | IS: 216; C: 76 | <24 h: 2 | HC: 1; NS: 1 | 0 | 0 | | | | | |
| miR-28-5p | 1 | | B: 1 | IS: 55; C: 2360 | <24 h: 1 | NS: 1 | 0 | 0 | | | | | |
| miR-296 | 0 | | | | | | 0 | 1 | | B: 1 | IS: 106; C: 110 | <24 h: 1 | NS: 1 |
| miR-29b | 1 | | B: 1 | IS: 30; C: 30 | <24 h: 1 | NS: 1 | 0 | 0 | | | | | |
| miR-29b-2-5p | 1 | | B: 1 | IS: 80; C: 2360 | <24 h: 1 | NS: 1 | 0 | 0 | | | | | |
| miR-29b-3p | 1 | | B: 1 | IS: 227; C: 92 | <24 h: 1 | NS: 1 | 0 | 0 | | | | | |
| miR-301a-3p | 0 | | | | | | 0 | 1 | | B: 1 | IS: 10; C: 11 | <24 h: 1 | HC: 1 |
| miR-30a | 0 | | | | | | 0 | 1 | H: 1 | B: 1 | IS: 38; C: 50 | <24 h: 1 | HC: 1 |
| miR-30a-5p | 2 | L: 1 | B: 1; E: 1 | IS: 48; C: 48 | <6 h: 1; <9 h: 1; <24 h: 2 | HC: 1; NS: 1 | 0 | 2 | | B: 2 | IS: 48; C: 60 | <24 h: 2 | HC: 2 |
| miR-30c | 0 | | | | | | 0 | 1 | U: 1 | B: 1 | IS: 169; C: 24 | <24 h: 1 | HC: 1 |
| miR-30d-5p | 1 | | B: 1 | IS: 17; C: 25 | <24 h: 1 | NS: 1 | 0 | 1 | | B: 1 | IS: 34; C: 11 | <24 h: 1 | HC: 1 |
| miR-31-5p | 1 | | B: 1 | IS: 10; C: 10 | <24 h: 1 | NS: 1 | 0 | 0 | | | | | |
| miR-3149 | 1 | | B: 1 | IS: 117; C: 82 | <24 h: 1 | HC: 1 | 0 | 0 | | | | | |
| miR-32-3p | 2 | U: 1 | B: 2 | IS: 234; C: 164 | <24 h: 2 | HC: 2 | 0 | 0 | | | | | |
| miR-32-5p | 0 | | | | | | 0 | 1 | | B: 1 | IS: 10; C: 11 | <24 h: 1 | HC: 1 |
| miR-320a | 1 | | Ce: 1 | IS: 19; C: 20 | <24 h: 1 | HC: 1 | 0 | 0 | | | | | |
| miR-320a-3p | 1 | | B: 1 | IS: 19; C: 5 | <24 h: 1 | HC: 1 | 0 | 0 | | | | | |

| | | | | | | | | | | | | | | | | | | |
|---|---|---|---|---|---|---|---|---|---|---|---|---|---|---|---|---|---|---|
| miR-320b | 2 | | B: 2 | IS: 97; C: 2385 | <24 h: 2 | NS: 2 | 0 | | | | | | 1 | U: 1 | B: 1 | IS: 169; C: 24 | <24 h: 1 | HC: 1 |
| miR-320d | 1 | | B: 1 | IS: 17; C: 25 | <24 h: 1 | NS: 1 | 0 | | | | | | 3 | U: 2 | B: 2; Ce: 1 | IS: 329; C: 164 | <24 h: 3 | HC: 3 |
| miR-320e | 1 | | B: 1 | IS: 17; C: 25 | <24 h: 1 | NS: 1 | 0 | | | | | | 1 | U: 1 | B: 1 | IS: 136; C: 116 | <24 h: 1 | HC: 1 |
| miR-324-3p | 1 | | B: 1 | IS: 55; C: 2360 | <24 h: 1 | NS: 1 | 0 | | | | | | 0 | | | | | |
| miR-328-3p | 0 | | | | | | 0 | | | | | | 1 | | B: 1 | IS: 39; C: 20 | <24 h: 1 | NS: 1 |
| miR-330-3p | 0 | | | | | | 0 | | | | | | 1 | | B: 1 | IS: 10; C: 10 | <24 h: 1 | HC: 1 |
| miR-335-3p | 0 | | | | | | 0 | | | | | | 1 | | B: 1 | IS: 17; C: 25 | <24 h: 1 | NS: 1 |
| miR-335-5p | 0 | | | | | | 0 | | | | | | 1 | | B: 1 | IS: 10; C: 11 | <24 h: 1 | HC: 1 |
| miR-338 | 1 | L: 1 | CSF: 1 | IS: 9; C: 12 | over: 1 | HC: 1 | 0 | | | | | | 0 | | | | | |
| miR-33a-5p | 0 | | | | | | 0 | | | | | | 1 | | B: 1 | IS: 10; C: 10 | <24 h: 1 | HC: 1 |
| miR-340 | 0 | | | | | | 0 | | | | | | 1 | U: 1 | B: 1 | IS: 169; C: 24 | <24 h: 1 | HC: 1 |
| miR-340-5p | 0 | | | | | | 0 | | | | | | 1 | | B: 1 | IS: 10; C: 11 | <24 h: 1 | HC: 1 |
| miR-342-3p | 0 | | | | | | 1 | L: 1 | B: 1 | IS: 23; C: 35 | over: 1 | OC: 1 | 0 | | | | | |
| miR-34a-5p | 1 | | B: 1 | IS: 102; C: 97 | <24 h: 1 | HC: 1 | 0 | | | | | | 0 | | | | | |
| miR-362-3p | 0 | | | | | | 0 | | | | | | 1 | | B: 1 | IS: 10; C: 11 | <24 h: 1 | HC: 1 |
| miR-363-3p | 1 | | Ce: 1 | IS: 24; C: 24 | <24 h: 1 | HC: 1 | 0 | | | | | | 0 | | | | | |
| miR-376a-3p | 0 | | | | | | 1 | L: 1 | B: 1 | IS: 23; C: 35 | over: 1 | OC: 1 | 0 | | | | | |
| miR-376c-3p | 1 | | Ce: 1 | IS: 19; C: 20 | <24 h: 1 | HC: 1 | 0 | | | | | | 0 | | | | | |
| miR-377-5p | 0 | | | | | | 0 | | | | | | 1 | | B: 1 | IS: 117; C: 82 | <24 h: 1 | HC: 1 |
| miR-378 | 0 | | | | | | 0 | | | | | | 2 | H: 1 | B: 2 | IS: 212; C: 220 | <24 h: 2 | HC: 1; NS: 1 |
| miR-378a-5p | 0 | | | | | | 0 | | | | | | 1 | | B: 1 | IS: 106; C: 110 | <24 h: 1 | HC: 1 |
| miR-379-5p | 0 | | | | | | 0 | | | | | | 1 | | B: 1 | IS: 47; C: 96 | <24 h: 1 | HC: 1 |
| miR-382-5p | 0 | | | | | | 0 | | | | | | 1 | U: 1 | B: 1 | IS: 78; C: 39 | <24 h: 1 | HC: 1 |
| miR-411-5p | 0 | | | | | | 0 | | | | | | 1 | | B: 1 | IS: 47; C: 96 | <24 h: 1 | HC: 1 |
| miR-422a | 1 | U: 1 | B: 1 | IS: 169; C: 24 | <24 h: 1 | HC: 1 | 0 | | | | | | 0 | | | | | |
| miR-423-3p | 0 | | | | | | 0 | | | | | | 1 | U: 1 | B: 1 | IS: 169; C: 24 | <24 h: 1 | HC: 1 |
| miR-423-5p | 1 | | B: 1 | IS: 117; C: 82 | <24 h: 1 | HC: 1 | 0 | | | | | | 0 | | | | | |
| miR-424 | 2 | U: 1 | Ce: 2 | IS: 80; C: 54 | <6 h: 1; <9 h: 1; <24 h: 2 | HC: 1; NS: 1 | 0 | | | | | | 0 | | | | | |
| miR-424-5p | 1 | | B: 1 | IS: 142; C: 50 | <24 h: 1 | HC: 1 | 0 | | | | | | 0 | | | | | |
| miR-4306 | 1 | U: 1 | B: 1 | IS: 136; C: 116 | <24 h: 1 | HC: 1 | 0 | | | | | | 0 | | | | | |
| miR-432-5p | 1 | | Ce: 1 | IS: 19; C: 20 | <24 h: 1 | HC: 1 | 0 | | | | | | 0 | | | | | |
| miR-433-5p | 1 | L: 1 | B: 1 | IS: 23; C: 35 | over: 1 | OC: 1 | 0 | | | | | | 0 | | | | | |
| miR-4429 | 0 | | | | | | 0 | | | | | | 1 | | Ce: 1 | IS: 24; C: 24 | <24 h: 1 | HC: 1 |

| | | | | | | | | | | | | | |
|---|---|---|---|---|---|---|---|---|---|---|---|---|---|
| miR-4446-3p | 0 | | | | | | 0 | 1 | | B: 1 | IS: 47; C: 96 | <24 h: 1 | HC: 1 |
| miR-4454 | 0 | | | | | | 0 | 2 | | B: 2 | IS: 50; C: 48 | <24 h: 2 | NS: 2 |
| miR-451a | 3 | | B: 3 | IS: 182; C: 198 | <24 h: 3 | HC: 3 | 0 | 0 | | | | | |
| miR-4523 | 0 | | | | | | 0 | 1 | U: 1 | B: 1 | IS: 22; C: 22 | <24 h: 1 | HC: 1 |
| miR-4634 | 1 | | B: 1 | IS: 17; C: 25 | <24 h: 1 | NS: 1 | 0 | 0 | | | | | |
| miR-4656 | 1 | | Ce: 1 | IS: 19; C: 20 | <24 h: 1 | HC: 1 | 0 | 0 | | | | | |
| miR-4739 | 1 | | B: 1 | IS: 117; C: 82 | <24 h: 1 | HC: 1 | 0 | 0 | | | | | |
| miR-483-3p | 0 | | | | | | 0 | 1 | | B: 1 | IS: 55; C: 2360 | <24 h: 1 | NS: 1 |
| miR-484 | 1 | | B: 1 | IS: 33; C: 23 | <24 h: 1 | NS: 1 | 0 | 1 | | B: 1 | IS: 80; C: 2360 | <24 h: 1 | NS: 1 |
| miR-485-3p | 0 | | | | | | 0 | 1 | | B: 1 | IS: 47; C: 96 | <24 h: 1 | HC: 1 |
| miR-487 | 1 | | Ce: 1 | IS: 19; C: 20 | <24 h: 1 | HC: 1 | 0 | 0 | | | | | |
| miR-487b-3p | 1 | | Ce: 1 | IS: 24; C: 24 | <24 h: 1 | HC: 1 | 0 | 0 | | | | | |
| miR-488 | 1 | U: 1 | B: 1 | IS: 169; C: 24 | <24 h: 1 | HC: 1 | 0 | 0 | | | | | |
| miR-502-5p | 0 | | | | | | 0 | 1 | U: 1 | B: 1 | IS: 169; C: 24 | <24 h: 1 | HC: 1 |
| miR-503-5p | 1 | | Ce: 1 | IS: 19; C: 20 | <24 h: 1 | HC: 1 | 0 | 0 | | | | | |
| miR-505-5p | 1 | | B: 1 | IS: 10; C: 11 | <24 h: 1 | HC: 1 | 0 | 0 | | | | | |
| miR-505â€"5p | 1 | U: 1 | B: 1 | IS: 22; C: 22 | <24 h: 1 | HC: 1 | 0 | 0 | | | | | |
| miR-5100 | 0 | | | | | | 0 | 1 | | B: 1 | IS: 17; C: 25 | <24 h: 1 | NS: 1 |
| miR-517b-3p | 0 | | | | | | 0 | 1 | | B: 1 | IS: 10; C: 11 | <24 h: 1 | HC: 1 |
| miR-518b | 0 | | | | | | 0 | 1 | | B: 1 | IS: 117; C: 82 | <24 h: 1 | HC: 1 |
| miR-523-3p | 2 | L: 1 | CSF: 2 | IS: 20; C: 20 | <24 h: 1; over: 1 | HC: 1; NS: 1 | 0 | 0 | | | | | |
| miR-532-5p | 0 | | | | | | 0 | 2 | U: 1 | B: 2 | IS: 234; C: 164 | <24 h: 2 | HC: 2 |
| miR-544a | 1 | | B: 1 | IS: 10; C: 11 | <24 h: 1 | HC: 1 | 0 | 0 | | | | | |
| miR-549 | 1 | U: 1 | B: 1 | IS: 169; C: 24 | <24 h: 1 | HC: 1 | 0 | 0 | | | | | |
| miR-550b-2-5p | 0 | | | | | | 0 | 1 | U: 1 | B: 1 | IS: 22; C: 22 | <24 h: 1 | HC: 1 |
| miR-570 | 0 | | | | | | 0 | 1 | | B: 1 | | <24 h: 1 | NS: 1 |
| miR-574-3p | 1 | | B: 1 | IS: 55; C: 2360 | <24 h: 1 | NS: 1 | 0 | 1 | U: 1 | B: 1 | IS: 169; C: 24 | <24 h: 1 | HC: 1 |
| miR-574-5p | 0 | | | | | | 0 | 2 | U: 1 | B: 2 | IS: 216; C: 120 | <24 h: 2 | HC: 2 |
| miR-579-3p | 0 | | | | | | 0 | 1 | | B: 1 | IS: 10; C: 11 | <24 h: 1 | HC: 1 |
| miR-593 | 0 | | | | | | 0 | 1 | | B: 1 | | <24 h: 1 | NS: 1 |
| miR-605-3p | 1 | | B: 1 | IS: 5; C: 5 | <24 h: 1 | NS: 1 | 0 | 0 | | | | | |
| miR-605-5p | 1 | | B: 1 | IS: 5; C: 5 | <24 h: 1 | NS: 1 | 0 | 0 | | | | | |
| miR-617 | 1 | U: 1 | B: 1 | IS: 169; C: 24 | <24 h: 1 | HC: 1 | 0 | 0 | | | | | |

| | | | | | | | | | | | | | |
|---|---|---|---|---|---|---|---|---|---|---|---|---|---|
| miR-625-3p | 0 | | | | | | 0 | 1 | | B: 1 | IS: 55; C: 2360 | <24 h: 1 | NS: 1 |
| miR-627 | 1 | U: 1 | B: 1 | IS: 169; C: 24 | <24 h: 1 | HC: 1 | 0 | 0 | | | | | |
| miR-628-5p | 1 | | B: 1 | IS: 10; C: 11 | <24 h: 1 | HC: 1 | 0 | 0 | | | | | |
| miR-629-3p | 0 | | | | | | 0 | 1 | | B: 1 | IS: 80; C: 2360 | <24 h: 1 | NS: 1 |
| miR-630 | 0 | | | | | | 0 | 1 | | B: 1 | | <24 h: 1 | NS: 1 |
| miR-638 | 0 | | | | | | 0 | 1 | | B: 1 | IS: 22; C: 36 | <24 h: 1 | NS: 1 |
| miR-653 | 0 | | | | | | 0 | 1 | | B: 1 | | <24 h: 1 | NS: 1 |
| miR-660-5p | 0 | | | | | | 0 | 1 | | B: 1 | IS: 10; C: 11 | <24 h: 1 | HC: 1 |
| miR-664a-5p | 1 | | B: 1 | IS: 10; C: 11 | <24 h: 1 | HC: 1 | 0 | 0 | | | | | |
| miR-6721-5p | 0 | | | | | | 0 | 1 | | B: 1 | IS: 47; C: 96 | <24 h: 1 | HC: 1 |
| miR-676-3p | 0 | | | | | | 0 | 1 | | B: 1 | IS: 47; C: 96 | <24 h: 1 | HC: 1 |
| miR-6795-3p | 0 | | | | | | 0 | 1 | U: 1 | B: 1 | IS: 22; C: 22 | <24 h: 1 | HC: 1 |
| miR-7-2-3p | 1 | | B: 1 | IS: 87; C: 13 | <24 h: 1 | NS: 1 | 0 | 0 | | | | | |
| miR-874-3p | 0 | | | | | | 0 | 1 | | Ce: 1 | IS: 19; C: 20 | <24 h: 1 | HC: 1 |
| miR-877-5p | 1 | | B: 1 | IS: 10; C: 11 | <24 h: 1 | HC: 1 | 0 | 0 | | | | | |
| miR-886-5p | 0 | | | | | | 0 | 1 | U: 1 | B: 1 | IS: 169; C: 24 | <24 h: 1 | HC: 1 |
| miR-9 | 0 | | | | | | 0 | 1 | L: 1 | B: 1 | IS: 31; C: 11 | <24 h: 1 | HC: 1 |
| miR-9-3p | 1 | | CSF: 1 | IS: 21; C: 21 | <24 h: 1 | NS: 1 | 0 | 0 | | | | | |
| miR-9-5p | 1 | | B: 1 | | <24 h: 1 | NS: 1 | 0 | 2 | | B: 2 | IS: 41; C: 21 | <24 h: 2 | HC: 2 |
| miR-92a | 0 | | | | | | 0 | 1 | U: 1 | B: 1 | IS: 169; C: 24 | <24 h: 1 | HC: 1 |
| miR-93 | 0 | | | | | | 0 | 2 | U: 2 | B: 2 | IS: 202; C: 44 | <6 h: 1; <9 h: 1; <24 h: 2 | HC: 2 |
| miR-93-5p | 2 | | B: 2 | IS: 172; C: 57 | <24 h: 2 | HC: 1; NS: 1 | 0 | 0 | | | | | |
| miR-Let-7i | 0 | | | | | | 0 | 1 | H: 1 | B: 1 | IS: 106; C: 106 | <24 h: 1 | HC: 1 |
| miRNA-221-3p | 0 | | | | | | 0 | 1 | L: 1 | B: 1 | IS: 78; C: 39 | <6 h: 1; <9 h: 1; <24 h: 1 | HC: 1 |
| miRNA-335 | 0 | | | | | | 0 | 1 | L: 1 | B: 1 | IS: 168; C: 104 | <24 h: 1 | HC: 1 |
| miRNA-382-5p | 0 | | | | | | 0 | 1 | L: 1 | B: 1 | IS: 78; C: 39 | <6 h: 1; <9 h: 1; <24 h: 1 | HC: 1 |
