## Supplementary Material_Strings for "Early-Phase Fluid Diagnostic Biomarkers in Acute Ischemic Stroke: An Umbrella Review"

#### ***Research question***

*Which fluid biomarkers[I] can be useful for the diagnosis[O] of ischemic stroke[P] vs stroke mimics[C]?*

### Strings

MEDLINE

*String [Results: 668]*

*("Stroke"[Mesh] OR "Brain Ischemia"[Mesh] OR "Brain Infarction"[Mesh] OR stroke[tiab] OR ischemic stroke[tiab] OR cerebral ischemia[tiab])*
*AND*
*("Biomarkers"[Mesh] OR "Cerebrospinal Fluid"[Mesh] OR "Blood"[Mesh] OR biomarker*[tiab] OR "Activins"[Mesh] OR "Activated-leukocyte Cell Adhesion Molecule"[Mesh] OR "Adiponectin"[Mesh] OR "Alarmins"[Mesh] OR "alpha 1-antichymotrypsin"[Mesh] OR "alpha-Macroglobulins"[Mesh] OR "Anaphylatoxins"[Mesh] OR "Angiotensins"[Mesh] OR "Antifibrinolytic Agents"[Mesh] OR "Antithrombin III"[Mesh] OR "Apolipoproteins"[Mesh] OR "Aquaporin 4"[Mesh] OR "Aryldialkylphosphatase"[Mesh] OR "Beta 2-Glycoprotein I"[Mesh] OR “Bilirubin”[Mesh] OR "Brain-Derived Neurotrophic Factor"[Mesh] OR "C-Reactive Protein"[Mesh] OR "Cadherins"[Mesh] OR "Carcinoembryonic Antigen"[Mesh] OR "Caspases"[Mesh] OR "Cathepsin B"[Mesh] OR "CD146 Antigen"[Mesh] OR "CD24 Antigen"[Mesh] OR "Platelet Endothelial Cell Adhesion Molecule-1"[Mesh] OR "CD4 Immunoadhesins"[Mesh] OR "CD40 Antigens"[Mesh] OR "CD40 Ligand"[Mesh] OR "Cell Adhesion Molecules"[Mesh] OR "Cell Adhesion Molecules, Neuronal"[Mesh] OR "Ceruloplasmin"[Mesh] OR "Chimerin 1"[Mesh] OR "Chimerin Proteins"[Mesh] OR "Chitinases"[Mesh] OR "Cholesterol Ester Transfer Proteins"[Mesh] OR "Chromogranin A"[Mesh] OR "Clusterin"[Mesh] OR "Complement C1"[Mesh] OR "Complement C2"[Mesh] OR "Complement C3"[Mesh] OR "Complement C4"[Mesh] OR "Complement C5"[Mesh] OR "Complement C6"[Mesh] OR "Complement C7"[Mesh] OR "Complement C8"[Mesh] OR "Complement C9"[Mesh] OR "Complement Activating Enzymes"[Mesh] OR "Complement FactOR B"[Mesh] OR "Complement InactivatOR Proteins"[Mesh] OR "Complement Membrane Attack Complex"[Mesh] OR "Complement System Proteins"[Mesh] OR "E-Selectin"[Mesh] OR "Endothelial ProgenitOR Cells"[Mesh] OR "Endothelins"[Mesh] OR "Erythropoietin"[Mesh] OR "Ferritins"[Mesh] OR "Fibronectins"[Mesh] OR "Fibrinogen"[Mesh] OR "Fructose-Bisphosphate Aldolase"[Mesh] OR "Interleukins"[Mesh] OR "FactOR V"[Mesh] OR "FactOR VII"[Mesh] OR "FactOR VIII"[Mesh] OR "FactOR IX"[Mesh] OR "FactOR X"[Mesh] OR "FactOR XI"[Mesh] OR "FactOR XII"[Mesh] OR "FactOR XIIa"[Mesh] OR "Fatty Acid-Binding Proteins"[Mesh] OR "Fatty Acids, Nonesterified"[Mesh] OR "Fibrin Fibrinogen Degradation Products"[Mesh] OR "Fibroblast Growth FactOR 2"[Mesh] OR "Fibrinopeptide A"[Mesh] OR "Fibrinopeptide B"[Mesh] OR "Follistatin"[Mesh] OR "Follistatin-Related Proteins"[Mesh] OR "gamma-Aminobutyric Acid"[Mesh] OR "Glial Fibrillary Acidic Protein"[Mesh] OR "Growth Differentiation FactOR 15"[Mesh] OR "Glutathione Transferase"[Mesh] OR "Granulocyte-Macrophage Colony-Stimulating Factor"[Mesh] OR "Growth Hormone"[Mesh] OR "Haptoglobins"[Mesh] OR*

*"Hemopexin"[Mesh] OR "Heparin CofactOR II"[Mesh] OR "Human Growth Hormone"[Mesh] OR "Immunoglobulin G"[Mesh] OR "HMGB1 Protein"[Mesh] OR "Hydrocortisone"[Mesh] OR "Inhibin-beta Subunits"[Mesh] OR "Intercellular Adhesion Molecule-1"[Mesh] OR "Interleukin-33"[Mesh] OR "Interleukin-6"[Mesh] OR "Matrix Metalloproteinase 9"[Mesh] OR "MicroRNAs"[Mesh] OR "Natriuretic Peptide, Brain"[Mesh] OR "Orosomucoid"[Mesh] OR "Peptidyl-Dipeptidase A"[Mesh] OR "Phosphopyruvate Hydratase"[Mesh] OR "Procalcitonin"[Mesh] OR "Proteomics"[Mesh] OR "ReceptOR fOR Advanced Glycation End Products"[Mesh] OR "Receptors, Lymphocyte Homing"[Mesh] OR "Retinol-Binding Proteins, Plasma"[Mesh] OR "S100 Calcium Binding Protein beta Subunit"[Mesh] OR "Selectins"[Mesh] OR "Tau Proteins"[Mesh] OR "Triggering ReceptOR Expressed on Myeloid Cells-1"[Mesh] OR "TumOR Necrosis Factor-alpha"[Mesh] OR "Vascular Cell Adhesion Molecule-1"[Mesh] OR "von Willebrand Factor"[Mesh] OR activins[tiab] OR adiponectin[tiab] OR aldolase[tiab] OR "alpha-Macroglobulins"[tiab] OR "alpha 1-antichymotrypsin"[tiab] OR angiotensin*[tiab] OR antiplasmin[tiab] OR antithrombin[tiab] OR apolipoprotein*[tiab] OR AQP4[tiab] OR "beta 2-Glycoprotein I"[tiab] OR blood glucose[tiab] OR caspase*[tiab] OR "cathepsin B"[tiab] OR cd34[tiab] OR cd40[tiab] OR ceruloplasmin[tiab] OR chitinase[tiab] OR "cholesterol ester transfer proteins"[tiab] OR chromogranin[tiab] OR clusterin[tiab] OR copeptin[tiab] OR D-dimer[tiab] OR endothelial progenitOR cells[tiab] OR "Fibroblast Growth FactOR 2"[tiab] OR ferritin[tiab] OR fibronectin*[tiab] OR GABA[tiab] OR GDF15[tiab] OR GFAP[tiab] OR HMGB1[tiab] OR ICAM-1[tiab] OR inhibins[tiab] OR MMP9[tiab] OR MR-proANP[tiab] OR neurofilament[tiab] OR neutrophil-to-lymphocyte[tiab] OR NfL[tiab] OR NT-proBNP[tiab] OR orosomucoid[tiab] OR panel[tiab] OR "Peptidyl-Dipeptidase A"[tiab] OR procalcitonin[tiab] OR RAGE[tiab] OR RBP4[tiab] OR remnant cholesterol[tiab] OR S100B[tiab] OR TREM1[tiab] OR TREM2[tiab] OR TNF-alpha[tiab] OR tau[tiab] OR vWF[tiab] OR molecule[tiab] OR bilirubin[tiab])*
*AND*
*("Diagnosis"[Mesh] OR diagnos*[tiab] OR "differential diagnosis"[tiab] OR detect*[tiab])*

Time limits: inception to May 10rd, 2025
Filters: Systematic review, Meta-analysis, Human

<u>Epistemonikos</u>

*String [Results: 214]*

*(title:(("stroke" OR "ischemic stroke" OR "brain ischemia" OR "brain infarction" OR "cerebral ischemia") AND ("biomarker*" OR "cerebrospinal fluid" OR "blood" OR "plasma" OR "serum" OR "C-reactive protein" OR "S100B" OR "interleukin-6" OR "interleukin-33" OR "NT-proBNP" OR "MR-proANP" OR "GFAP" OR "NfL" OR "neurofilament" OR "neuron-specific enolase" OR "MMP9" OR "copeptin" OR "cystatin C" OR "BDNF" OR "TREM2" OR "TREM1" OR "GDF15" OR "neutrophil-to-lymphocyte ratio" OR "fibrinogen" OR "ferritin" OR "D-dimer" OR "HMGB1" OR "AQP4" OR "procalcitonin" OR "mannose-binding lectin" OR "adipocyte fatty acid-binding protein" OR "cortisol" OR "receptOR fOR advanced glycation end-*

*products" OR "intercellular adhesion molecule-1" OR "von Willebrand factor" OR "matrix metalloproteinase-9" OR "interleukin-6" OR "tumOR necrosis factor-alpha" OR "activated protein C" OR "tau protein" OR "gamma aminobutyric acid" OR "blood glucose" OR "endothelial progenitOR cells" OR "CD34-positive cells" OR "miRNA" OR "panel" OR "RBP-4" OR "remnant cholesterol")*
*AND ("diagnosis" OR "early diagnosis" OR "diagnostic techniques OR "procedures" OR "differential diagnosis" OR "detection")) OR abstract:(("stroke" OR "ischemic stroke" OR "brain ischemia" OR "brain infarction" OR "cerebral ischemia") AND ("biomarker*" OR "cerebrospinal fluid" OR "blood" OR "plasma" OR "serum" OR "C-reactive protein" OR "S100B" OR "interleukin-6" OR "interleukin-33" OR "NT-proBNP" OR "MR-proANP" OR "GFAP" OR "NfL" OR "neurofilament" OR "neuron-specific enolase" OR "MMP9" OR "copeptin" OR "cystatin C" OR "BDNF" OR "TREM2" OR "TREM1" OR "GDF15" OR "neutrophil-to-lymphocyte ratio" OR "fibrinogen" OR "ferritin" OR "D-dimer" OR "HMGB1" OR "AQP4" OR "procalcitonin" OR "mannose-binding lectin" OR "adipocyte fatty acid-binding protein" OR "cortisol" OR "receptOR fOR advanced glycation end-products" OR "intercellular adhesion molecule-1" OR "von Willebrand factor" OR "matrix metalloproteinase-9" OR "interleukin-6" OR "tumOR necrosis factor-alpha" OR "activated protein C" OR "tau protein" OR "gamma aminobutyric acid" OR "blood glucose" OR "endothelial progenitOR cells" OR "CD34-positive cells" OR "miRNA" OR "panel" OR "RBP-4" OR "remnant cholesterol")*
*AND ("diagnosis" OR "early diagnosis" OR "diagnostic techniques" OR "diagnostic procedures" OR "differential diagnosis" OR "detection")))*

Time limits: Inception to May 10rd, 2025
Filters: Systematic Reviews

### Cochrane Database of Systematic Reviews

*String [Results: 28]*

*([mh "Stroke"] OR [mh "Brain Ischemia"] OR [mh "Brain Infarction"] OR stroke:ti,ab,kw OR ischemic stroke:ti,ab,kw OR cerebral ischemia:ti,ab,kw OR "brain infarction":ti,ab,kw)*
*AND*
*([mh "Biomarkers"] OR [mh "Cerebrospinal Fluid"] OR [mh "Blood"] OR [mh "C-Reactive Protein"] OR [mh "S100 Calcium Binding Protein beta Subunit"] OR [mh "Interleukin-6"] OR [mh "Interleukin-33"] OR [mh "Interleukins"] OR [mh "Glial Fibrillary Acidic Protein"] OR [mh "Phosphopyruvate Hydratase"] OR [mh "Matrix Metalloproteinase 9"] OR [mh "Glycopeptides"] OR [mh "Cystatins"] OR [mh "Brain-Derived Neurotrophic Factor"] OR [mh "Growth Differentiation FactOR 15"] OR [mh "Fibrinogen"] OR [mh "Ferritins"] OR [mh "Fibrin Fibrinogen Degradation Products"] OR [mh "HMGB1 Protein"] OR [mh "Aquaporin 4"] OR [mh "Procalcitonin"] OR [mh "Proteomics"] OR [mh "Fatty Acid-Binding Proteins"] OR [mh "Hydrocortisone"] OR [mh "ReceptOR fOR Advanced Glycation End Products"] OR [mh "Intercellular Adhesion Molecule-1"] OR [mh "von Willebrand Factor"] OR [mh "TumOR Necrosis Factor-alpha"] OR [mh "tau Proteins"] OR [mh "gamma-Aminobutyric Acid"] OR [mh "Endothelial ProgenitOR Cells"] OR [mh "MicroRNAs"] OR [mh "Retinol-Binding Proteins, Plasma"] OR "NT-proBNP":ti,ab,kw OR "copeptin":ti,ab,kw OR "TREM2":ti,ab,kw*

*OR "TREM1":ti,ab,kw OR "neurofilament protein L":ti,ab,kw OR "midregional pro-atrial natriuretic peptide":ti,ab,kw OR "activated protein C":ti,ab,kw OR "blood glucose":ti,ab,kw OR "CD34-positive cells":ti,ab,kw OR "panel":ti,ab,kw OR "remnant-like particle cholesterol":ti,ab,kw OR "mannose-binding lectin":ti,ab,kw OR "neutrophil-to-lymphocyte":ti,ab,kw)*

*AND*

*([mh "Diagnosis"] OR diagnos*:ti,ab,kw OR "differential diagnosis":ti,ab,kw OR detect*:ti,ab,kw)*

Time limits: inception to May 10rd, 2025
Filters: Cochrane Reviews
